# Computational Mechanisms of Approach-Avoidance Conflict Predictively Differentiate Between Affective and Substance Use Disorders

**DOI:** 10.1101/2025.02.10.25321894

**Authors:** Marishka M. Mehta, Navid Hakimi, Orestes Pena, Taylor Torres, Carter M. Goldman, Claire A. Lavalley, Jennifer L. Stewart, Hannah Berg, Maria Ironside, Martin P. Paulus, Robin Aupperle, Ryan Smith

## Abstract

Psychiatric disorders are highly heterogeneous and often co-morbid, posing specific challenges for effective treatment. Recently, computational modeling has emerged as a promising approach for characterizing sources of this heterogeneity, which could potentially aid in clinical differentiation. In this study, we tested whether computational mechanisms of decision-making under approach-avoidance conflict (AAC) – where behavior is expected to have both positive and negative outcomes – may have utility in this regard. We first carried out a set of pre-registered modeling analyses in a sample of 480 individuals who completed an established AAC task. These analyses aimed to replicate cross-sectional and longitudinal results from a prior dataset (N=478) – suggesting that mechanisms of decision uncertainty (*DU*) and emotion conflict (*EC*) differentiate individuals with depression, anxiety, substance use disorders, and healthy comparisons. We then combined the prior and current datasets and employed a stacked machine learning approach to assess whether these computational measures could successfully perform out-of-sample classification between diagnostic groups. This revealed above-chance differentiation between affective and substance use disorders (balanced accuracy > 0.688), both in the presence and absence of co-morbidities. These results demonstrate the predictive utility of computational measures in characterizing distinct mechanisms of psychopathology and may point to novel treatment targets.

---

The presence of comorbid psychiatric disorders is associated with greater severity, chronicity, poorer treatment response, and overall functional impairment (Kessler et al., 2015; Klein Hofmeijer-Sevink et al., 2012; Plana-Ripoll et al., 2020; Plana-Ripoll et al., 2019). Comorbid psychiatric disorders show considerable heterogeneity, which can pose further difficulties for prognosis and treatment. Unfortunately, the mechanisms leading to this pattern of comorbidity and heterogeneity remain poorly understood, limiting development of more targeted treatments. It is expected that some of the mechanisms underlying psychiatric disorders are transdiagnostic and account for comorbidities (Dalgleish et al., 2020), while other mechanisms may differentiate clinical profiles that have distinct underlying causes. If so, developing more targeted and individualized treatments will likely require identification of both transdiagnostic and diagnosis-specific mechanisms.

One prominent set of cognitive and neurocomputational mechanisms of transdiagnostic relevance pertains to approach-avoidance conflict (AAC), where available choices are expected to lead to both positive and negative outcomes (Aupperle & Paulus, 2010). In depression and anxiety disorders (DEP/ANX), for example, maladaptive resolution of this conflict can manifest as avoidance of rewarding activities in anticipation of feared negative outcomes (Barlow et al., 2016). Similarly, individuals with substance use disorders (SUDs) often experience conflict between cravings, withdrawal, and the negative consequences of dependence, possibly leading to continued substance use despite loss of career, social support, and physical health. Given its broad relevance, a growing body of clinical research has emerged in recent years examining maladaptive decision processes under AAC in different clinical populations (for a detailed review, refer Letkiewicz et al. (2023)), with the goal of identifying novel and individualized treatment targets (Paulus, 2017). Within this body of work, several paradigms have been used to study AAC (reviewed by Kirlic et al. (2017)). More recently, computational modeling approaches have highlighted cognitive processes, such as suboptimal reward valuation, uncertainty, or inference, which may contribute to maladaptive AAC behavior c. However, the clinical utility of these findings remains limited by sparse evidence of replicability and longitudinal stability of computational markers. Such investigations will be crucial for progress in computational psychiatry and its continued effort to identify underlying mechanisms and predictors of treatment outcomes (Huys et al., 2021).

In previous work, we have investigated cross-sectional differences in, and longitudinal stability of, AAC behavior in transdiagnostic samples including healthy comparisons (HCs), individuals with DEP/ANX, and individuals with SUDs (Smith et al., 2021b; Smith et al., 2021c). This work made use of a novel computational model capable of disentangling two dimensions of AAC: the subjective value of negative stimuli, referred to as emotion conflict (*EC*), and the inconsistency in choice under identical conflict conditions, referred to as decision uncertainty (*DU*). We found that individuals in both clinical groups showed higher *DU* and lower *EC* than HCs, which was consistent at a 1-year follow-up visit. The cross-sectional results at baseline were also recently replicated in an independent sample (Smith et al., 2023), suggesting generalizability. However, the longitudinal findings remain to be replicated. It also remains unclear whether these model-based measures support out-of-sample prediction and whether they are fully transdiagnostic or might also differentiate clinical sub-populations.

Our previous work also found that observed group differences in *EC* were driven largely by females. This added to a growing literature on sex differences in diagnostic frequency, symptom experience, and coping mechanisms (Gobinath et al., 2017; Green et al., 2019; Kelly et al., 2008; Matud, 2004; Riecher-Rössler, 2017; Rutter et al., 2003). For example, females appear more prone to affective disorders (Green et al., 2019; Rutter et al., 2003), and specific difficulties in emotion regulation have also been associated with symptom severity in females but not males (Kelly et al., 2008). It is therefore possible that the sex differences we have observed in AAC behavior could contribute to these specific difficulties.

In the current study, we first carry out a pre-registered replication (https://osf.io/7hsx9) of our earlier longitudinal results and observed sex differences in a new clinical sample to support their generalizability. We then combine our prior and current datasets, affording greater statistical power to test whether narrower diagnostic groups (i.e., different affective or substance use disorders) may be differentiated by longitudinal dynamics in *DU* or *EC*. Finally, we use state-of-the-art machine learning approaches to test the out-of-sample predictive accuracy of these computational measures in clinical categorization. This provides a direct test of whether *DU* and *EC* levels are jointly capable of differentiating if an individual has an affective disorder, at least one SUD, or both, and thus whether they might underlie transdiagnostic or diagnosis-specific aspects of these disorders. Together, these results demonstrate how a computational characterization of behavior under AAC may provide both mechanistic and pragmatic information that could be useful in a clinical setting.

## Method

### Participants

This study leveraged a community-based sample (N=1050) from the Tulsa 1000 (T1000) study (Victor et al., 2018), which was recruited through radio, electronic media, treatment center referrals, and word of mouth. All participants were between the ages of 18–55 years. Transdiagnostic population screening was done using dimensional measures: Patient Health Questionnaire (PHQ; (Kroenke et al., 2001)) ≥ 10, Overall Anxiety Severity and Impairment Scale (OASIS; (Norman et al., 2006)) ≥ 8, and/or Drug Abuse Screening Test (DAST-10; (Bohn et al., 1991)) score ≥ 3. Healthy individuals did not show elevated symptoms or meet criteria for any psychiatric diagnosis. Exclusion criteria included a positive test for drugs of abuse; diagnosis of psychotic, bipolar, or obsessive-compulsive disorders; or reported history of moderate to severe traumatic brain injury, neurologic disorders, or severe or unstable medical conditions, active suicidal intent, or plan, or change in medication dose within 6 weeks. For detailed inclusion and exclusion criteria, see Victor et al. (2018).

As with prior work in the *exploratory* sample used in our prior reports (i.e., taken from the first 500 participants recruited to the T1000 study; (Smith et al., 2021b; Smith et al., 2021c)), the *confirmatory* sample (i.e., taken from the subsequent 550 participants recruited to the T1000 study; (Smith et al., 2023)) was divided into three groups: (i) HCs (baseline: N=97, 1-year follow-up: N=69); (ii) DEP/ANX (baseline: N=208, 1-year follow-up: N=135; including individuals with major depressive disorder with or without social anxiety disorder, generalized anxiety disorder, panic disorder, and/or posttraumatic stress disorder); (iii) SUDs (baseline: N=175, 1-year follow-up: N=83; with or without comorbid anxiety/depression). A full breakdown of co-morbidities is provided in **Supplementary Table 1**. Diagnostic grouping was performed in accordance with the DSM-IV or DSM-5 criteria using the Mini International Neuropsychiatric Inventory (MINI; (Sheehan et al., 1998)), which was administered at the baseline visit by a trained professional. Participant grouping was retained at 1-year follow-up, irrespective of change in diagnostic status (while analyses assessing change in symptoms over time tested for relevant potential effects; see **Supplementary Figure 1**).

### Data collection procedure

The T1000 protocol included intensive assessment of demographic, clinical, and psychiatric features, with a focus on negative and positive affect, arousal, and cognitive functioning. The complete list of assessments, along with their validity and reliability, are reported elsewhere (Victor et al., 2018). In this study, we examine the following measures, which were collected at baseline and 1-year follow-up: Patient-Reported Outcomes Measurement Information System (PROMIS) depression and anxiety scales (Cella et al., 2010), the Behavioral Activation/Inhibition scales (BIS/BAS; (Carver & White, 1994)), the Positive and Negative Affect Schedule (PANAS; (Watson et al., 1988)), the Anxiety Sensitivity Index (ASI; (Sandin et al., 2001)), the Temporal Experience of Pleasure Scale (TEPS; (Gard et al., 2006)), and the State-Trait Anxiety Inventory (STAI; (Spielberger et al., 1970)). All data collection procedures were approved by the Western Institutional Review Board. All participants provided written informed consent before completion of the study protocol, in accordance with the Declaration of Helsinki, and were compensated for participation (ClinicalTrials.gov identifier: #NCT02450240).

### Approach-avoidance conflict (AAC) task

The AAC task (**Figure 1**) has been described in detail in our previous work (Aupperle et al., 2015; Aupperle et al., 2011; Smith et al., 2021b). Briefly, a picture of an avatar standing above a runway, at its starting position, was presented on each trial. Participants were asked to move the avatar to one of the nine positions on the runway, toward the cues presented on either side. The cue consisted of an image – sun or cloud – representing a positive or negative affective image-sound pair, respectively, that would be shown with higher probability as the avatar moved closer to the associated side (detailed below). There was also a rectangular bar on each side, where the height of the red fill represented reward points associated with the linked image-sound pair. There were five trial types across 60 trials (12 each), defined by the cues presented: avoid-threat (AV), approach-reward (APP), and three levels of conflict trials (CONF2, CONF4, CONF6). In AV trials, the cloud and sun images were presented on opposing sides, with 0 reward points associated with either. APP trials provided positive affective stimuli on both sides, where one side was associated with 0 points and the other with 2 points. Lastly, in conflict trials, cues presented always included the sun with 0 reward points and the cloud with levels of 2 (CONF2), 4 (CONF4), and 6 (CONF6) reward points. These reward points did not lead to additional monetary compensation. The affective images and sounds were sampled from the International Affective Picture System (IAPS; (Lang et al., 2008)), International Affective Digitized Sounds (IADS; (Lang & Bradley, 1999)), and other freely available audio files (refer to Aupperle et al. (2015) and Chrysikou et al. (2017)).

**Figure 1.**
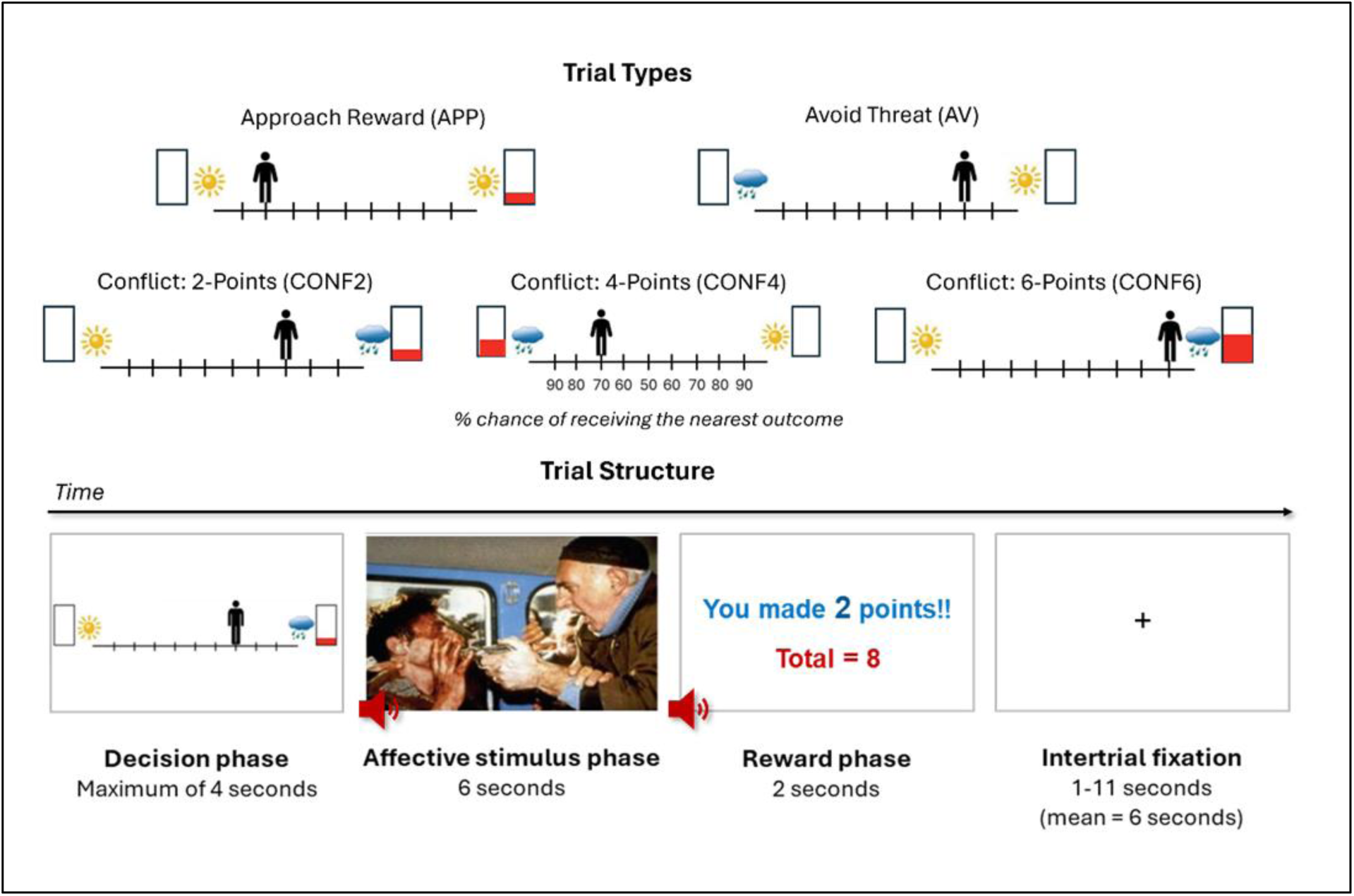
The approach-avoidance (AAC) task. ***Bottom***: Each trial is divided into a decision phase, an affective stimulus phase, and a reward phase. Trials are separated by a variable intertrial fixation time. ***Top***: During the decision phase, participants choose to move an avatar to one of nine positions on a runway. Pictures are presented on each side of the runway, indicating the types of stimuli that could be presented during the affective stimulus and reward phases. The sun and cloud images represented potential positive and negative affective stimuli, respectively (each being an image–sound combination). The height of the red fill in a rectangle signified the number of points that would be received in the reward phase (ranging from 0 to 6 points). Participants were instructed that the final position of the avatar determined the probability of each of these outcomes occurring (in increments of 10%, from 90% to 10% with each step away from the associated stimulus indicator images). The five trial types and associated probabilities of each outcome at each runway position. The task consisted of 60 trials, with 12 of each of the five trial types.

Model-free task measures included: 1) average chosen runway position; 2) choice variability, measured as within-subject standard deviation in chosen runway position (i.e., measuring response consistency); and 3) response times (RTs, i.e., time to initial button press) across trials.

Participants also completed a short post-task Likert scale survey (specific questions discussed below).

### Computational modeling

As in our previous studies (Smith et al., 2021b; Smith et al., 2021c; Smith et al., 2023), a two-parameter partially observable Markov decision process (POMDP) framework (**Table 1**) was used to model AAC task behavior. The model estimated parameters reflecting decision uncertainty (*DU*) and the subjective aversion to negative stimuli (i.e., relative to subjective value assigned to each point that could be won; “emotion conflict” [*EC*]). Higher *EC* values reflect greater avoidance of negative image-sound pairs, depending on the reward point value on offer. Higher *DU* values suggest greater variability in choice under identical task conditions and the tendency to adopt choices nearer to the middle of the runway. Thus, these parameters capture two dimensions of AAC.

**Table 1.** Markov decision process model of the AAC task.

| Model variable | General definition | Model-specific specification |
| --- | --- | --- |
| $o_t$ | Observable outcomes at time $t$. | 1. Observed position on the runway (10 possible observations. This included a “starting” position and the nine final positions on the runway that could be chosen).<br>2. Cues indicating trial type (five possible observations, corresponding to the five trial types).<br>3. Stimuli observed at the end of each trial. This formally included seven possible observations: a “starting” observation, the positive stimulus with 0 or 2 points, and the negative affective stimulus with 0, 2, 4, or 6 points. |
| $s_t$ | Beliefs about hidden states at time $t$. | 1. Beliefs about position on the runway: 10 possible belief states with an identity mapping to the observations in $o_t$ (runway position).<br>2. Beliefs about trial type: 5 possible belief states (corresponding to the five trial types) with an identity mapping to the observations in $o_t$ (trial types). |
| $\pi$ | A distribution over action options encoding the probability of choosing each action. | Encodes all the allowable actions. Here, actions are decisions to transition from the starting state to each of the nine possible runway positions. |
| $\beta$ ($DU$) | A ‘temperature’ parameter within a softmax choice function. Higher values for this parameter reduce the likelihood that actions expected to generate the most preferred outcomes will be chosen. | Higher $DU$ values reflect lower confidence about the best decision and lead to less deterministic action selection. This means that a participant is more likely to choose locations in the middle of the runway and more likely to make inconsistent choices across trials. Therefore, $DU$ encodes participants’ decision uncertainty during the task. |
| **A matrix**<br>$P(o_t\vert s_t)$ | A matrix encoding beliefs about the relationship between hidden states and observable outcomes (i.e., the likelihood that specific outcomes will be observed given specific hidden states). | Encodes beliefs about the relationship between position on the runway and the probability of observing each outcome, given beliefs about trial type. |
| **B matrix**<br>$P(s_{t+1}\vert s_t, \pi)$ | A matrix encoding transition probabilities, or beliefs about how hidden states will evolve over time depending on choice of action. | Encodes beliefs about all the possible ways participants could choose to move the avatar, along with the belief that the task condition (trial type) will not change within a trial. |
| **C matrix**<br>**$\ln P(o_t\vert C)$** | A matrix encoding the degree to which some observed outcomes are preferred over others at each time point $t$ (technically modeled as prior expectations conditioned on subjective stimulus value). The values for each column (representing $t$) in this matrix are passed through a softmax function to generate a probability distribution, which is then log-transformed. | Encodes stronger positive preferences for receiving higher amounts of points, and negative preferences for the aversive stimuli. These values are relative to an anchor value of 0 (for the “safe” positive stimulus). The **$EC$** parameter encodes the value of participants’ preferences against observing the aversive stimuli (i.e., prior to being passed through the softmax function and log-transformed). This value therefore competes against the value of points that could be won (e.g., if $EC = 4$, this means at least four reward points are needed to outweigh the unpleasantness of the aversive stimuli). |
| **D matrix**<br>**$P(s_1)$** | A matrix encoding beliefs about (a probability distribution over) initial hidden states. | The simulated participant always begins in an initial starting state and believes that each trial type is equally likely (before observing the trial type cues). |

**Note:** $DU$ = decision uncertainty; $EC$ = emotion conflict. Trial types: Approach (APP), Avoid (AV), Conflict (2: CONF2, 4:CONF4, 6: CONF6)

Parameter estimates were optimized by maximizing the likelihood of participants’ choice behavior using a standard Bayesian approach called Variational Laplace (Friston et al., 2007). Model simulations were done using the spm_MDP_VB_X.m MATLAB script, within the freely available SPM academic software package (http://www.fil.ion.ucl.ac.uk/spm/). MATLAB scripts used to model the task are provided in Appendix 3 of Smith et al. (2021b).

We also examined if the choice-based model could capture aspects of decision deliberation reflected by RTs. Here, a trial-by-trial measure of *choice uncertainty* was calculated as the entropy of the probability distribution over actions (*π*):

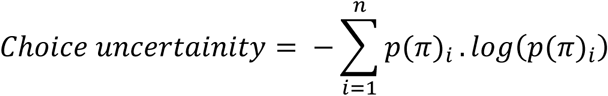

Pearson correlation coefficients were computed for each participant between trial-by-trial RTs and *choice uncertainty*. A linear mixed effects model (LME) was then used to test whether this association was significantly greater than 0 across participants, while controlling for the effects of group, time, their interaction, and individual-level random effects. Secondary analyses were performed to evaluate the relationship between RTs and chosen runway position, and RTs and model parameters. Here, LMEs were used to predict chosen runway position and model parameters based on group, time, and RTs, and possible *group x time* and *group x RT* interactions.

### Sample differences

Differences in participant characteristics were measured at the baseline and 1-year follow-up visit, and between the exploratory and confirmatory samples using frequentist and Bayesian approaches. For each group, possible differences in model parameters at baseline, follow-up, and their pre-post change were separately assessed using t-tests and regression. Equivalent Bayesian t-tests and regressions were performed to compute Bayes Factors (BFs; using the Bayesfactor package in R [https://richarddmorey.github.io/BayesFactor/]), which reflect the ratio of the probability of observed data under models with vs. without an effect of interest. Thus, BFs < 1 provide evidence in favor of a null model. For parameter change scores, these Bayesian regression models included sample type and baseline scores as predictors of change scores. The null model retained baseline scores as a predictor to isolate the effect of sample type (exploratory vs. confirmatory) from any potential confounding effects of baseline differences.

### Statistical analysis

All statistical analyses were performed using the R statistical package (2023; www.R-project.org/). Each analysis below follows the pre-registered analysis plan (https://osf.io/7hsx9) for replicating our prior results (Smith et al., 2021c). Additional analyses were conducted in the combined exploratory and confirmatory datasets to address more granular clinical group differences and out-of-sample classification of those with each disorder (see below for details).

#### Model validity

Model performance was evaluated using the average action probability and accuracy scores under the best-fit model for each participant, reflecting the average percentage of trials for which the action with the highest probability in the model matched the action chosen. The relationship between model parameters (*EC* and *DU*) and RTs was also examined, with the expectation that higher *EC* and *DU* would correlate with slower RTs. The relationship between parameter estimates and post-task Likert scales was also assessed, with the expectation that higher *EC* values would be associated with greater self-reported avoidance motivation and anxiety during the task, while higher *DU* would be associated with greater self-reported difficulty in making decisions on the task.

#### Within-subject stability

Consistency in scores over time (baseline to 1-year follow-up) was evaluated using single-measure consistency intra-class correlations [ICC(3, 1)]. These analyses were carried out for individual parameter estimates (*EC* and *DU*), model-free task measures (RTs, average chosen runway position, choice variability), clinical measures (PHQ, OASIS, and DAST), dimensional affective measures (ASI, BIS/BAS, PROMIS, PANAS, STAI, and TEPS), and post-task survey items. This was done across all participants and then further analyzed for each group separately. In line with prior suggestions, ICCs below 0.4 were considered ‘poor’, greater than 0.40 were considered ‘fair’, and greater than 0.60 were considered ‘good’ (Cicchetti, 1994).

#### Clinical differences

LMEs were used to investigate the impact of the group (sum coded; HC=-1) on model parameter estimates, model-free descriptive measures, and post-task scale items. These models also assessed the effect of time (sum coded: baseline = −1; follow-up = 1) and its interaction with group. Further LMEs also confirmed that the observed effects were not accounted for by age (centered) or biological sex (sum coded: female = −1; male = 1), along with their interaction with group. Similar to our previous report (Smith et al., 2023), an additional LME was performed controlling for the effect of WRAT Reading scores (a measure of premorbid cognitive ability (Johnstone et al., 1996)) and its interaction with group in a subset of participants with available data (N_Baseline_=391; N_Followup_=238).

Partial correlations were performed assessing whether baseline task measures (*EC*, *DU*, RTs, and average chosen runway position) predicted change in dimensional measures from baseline to follow-up (ASI, BIS/BAS, PROMIS, PANAS, STAI, and TEPS) while accounting for baseline symptom values. Similar analyses were also carried out in relation to symptom measures within the clinical groups. Further analyses also tested potential associations between changes in task measures and changes in symptoms over time (accounting for baseline values for each). Bayes factors were also calculated for these correlations to more strongly support null results in cases where frequentist analyses did not replicate previous findings. When computing these Bayes factors, the null hypothesis model included possible effects of baseline symptom levels.

#### Combined Sample Analyses

After completing pre-registered analyses, the exploratory (N_Baseline_=478; N_Follow-up_=324) and confirmatory (N_Baseline_=480; N_Follow-up_=287) samples were combined to ask novel questions benefiting from additional statistical power.

##### Disorder Specific analyses

The consistency of clinical differences in model parameters between HCs and more specific diagnostic categories (e.g., generalized anxiety disorder, social anxiety disorder, methamphetamine use disorder, opioid use disorder) within each group was examined using LMEs with the same structure described above: *parameter ∼ group*time + (1|participant)* and *parameter ∼ group + time + group*age + group\**sex *+ (1|participant)*. Each analysis was performed on a subsample including only the data from HCs and individuals with the specified disorder (irrespective of comorbidities; thus, some participants were included in more than one clinical group). Due to the small sample of individuals with hallucinogen use disorder (N_Baseline_=15; N_Follow-up_=4), this subgroup was not analyzed separately.

#### Predictive clinical categorization

To further assess the clinical utility of our computational model parameters, we utilized standard binary classification machine learning techniques, applying 5-fold cross-validation to predict group membership within our clinical sample (i.e., removing HCs from all analyses) using computational task metrics. In particular, we tested out-of-sample accuracy in classifying: 1) the exclusive presence of affective disorders or SUDs (i.e., with no comorbidity between the two; N=320 and 51, respectively); 2) the presence versus. absence of SUDs (with or without comorbid affective disorders; N=167 and 327, respectively); 3) the presence vs. absence of affective disorders (with or without comorbid SUDs; N=436 and 58, respectively); and 4) the presence vs. absence of comorbidity generally (i.e. those with only SUDs or only DEP/ANX vs. those with both types of disorders; N=123 and 371, respectively).

To do so, an established stacked ensemble approach was used (detailed within Ekhtiari et al. (2019)), in which three different classification approaches were selected. Their performance was compared, and an optimized combination of their predictions was then evaluated. The chosen algorithms were selected to represent different classes of classification approaches: elastic net (ENET), k-nearest neighbors (KNN), and Bagged AdaBoost (ADABAG). Briefly, ENET is a regularized logistic regression approach that combines lasso (L1) and ridge (L2) penalty terms. It is particularly useful in overcoming multicollinearity and avoiding over-fitting. A parameter *α* balances the proportion of the L1 and L2 penalties. A second parameter λ sets the weight on the loss function (parameter optimization is described further below). KNN is a non-parametric supervised learning algorithm. The parameter *k* refers to the number of nearest observations considered to predict the classification label. Finally, ADABAG combines bootstrap aggregation (Bagging) and adaptive boosting (AdaBoost) approaches to improve model accuracy and stability. AdaBoost iteratively classifies weak classifiers (decision trees), where each classifier uses previously misclassified observations. It adaptively increases the weight placed on currently misclassified observations while reducing the weight of data already classified correctly. This aids in reducing bias, variance, and overfitting. However, it can be susceptible to outliers and noisy data. To address this, bagging was used to generate datasets consisting of random samples (with replacement) from the original dataset. AdaBoost models were then trained on these generated datasets. This method improves consistency in prediction by combining the outputs from several weak classifiers, each trained on a generated dataset. AdaBoost has two parameters, *mfinal* and *maxdepth*, which control the number of boosting iterations and maximum tree depth, respectively.

The 5-fold cross-validation consisted of first dividing the dataset into five equal subsets. For each of the five rounds, four subsets were combined and used as a training dataset and the fifth was used as a testing dataset. In each round, the training dataset was also further divided into training (80%) and validation (20%) subsets. The validation subset was used for finding optimal hyperparameters. Here, seven different starting values were randomly selected for each hyper-parameter and a grid-search approach was used to find the optimal values. After parameter tuning was complete, the stacked ensemble approach then used linear aggregation to combine the predictions of each algorithm. Specifically, the weight on each algorithm was determined by respective model fit (area under the curve [AUC]), such that algorithms with better fits to the data were afforded higher weight in generating the stacked prediction. This process was repeated five times to get a stable comparison of fitted stacked models (STACK), and we report the average performance across these 5 folds. In addition, a variable importance (VI) metric was computed to measure the average scaled contribution (between 0 to 100) of each predictor. Note that, due to the unbalanced sample size, an up-sampling approach was also used during the cross-validation process.

Our approach focused on sequentially assessing the degree to which incorporating information derived from our computational model could improve prediction beyond basic demographic variables (age and sex). As such, we tested and compared models with five sets of predictors: 1) age and sex alone; 2) model parameters (*EC* and *DU* at baseline and follow-up) alone; 3) model parameters with age and sex; 4) model parameters and *r* values representing the correlation between trial-by-trial *choice uncertainty* and RTs; and 5) model parameters, *r* values, and age and sex. Because prior and current results show consistent *group x age* and *group x sex* interactions in predicting *DU* and *EC*, inclusion of demographic variables allowed us to test if they were sufficient to predict clinical grouping on their own (i.e., the observed differences are a function of sample characteristics) or whether model-based information led to substantial improvement in classification. All analyses were conducted using the caretStack library (https://github.com/kforthman/caretStack) in R. In addition, glmnet, Matrix, adabag and plyr libraries were used for base models. A full list of available base models can be found at https://topepo.github.io/caret/available-models.html.

## Results

### Participant Characteristics

Participant demographic and clinical measures are shown in **Table 2**. The descriptive statistics for model-based measures (*DU* and *EC*) are shown in **Table 3**. These measures were weakly correlated (*r*=0.14, *p=*0.020). As in our previous reports, parameters were log-transformed to minimize skew for subsequent analyses using the *optLog* R package (https://github.com/kforthman/optLog). The resulting log-transformed values were: HCs (*DU* = 0.73±0.96; *EC* = 1.06±0.87), DEP/ANX (*DU* = 0.70±1.03; *EC* = 0.83±0.78), SUDs (*DU* = 0.79±0.99; *EC* = 0.43±0.71).

**Table 2.** Summary statistics and group differences for demographic and clinical measures.

|  | **HCs** | **DEP/ANX** | **SUDs** | ***p*\*** |
| --- | --- | --- | --- | --- |
| Baseline | N = 97 | N = 208 | N = 175 |  |
| Follow-up | N = 69 | N = 135 | N = 83 |  |
| **Age** |  |  |  |  |
| Baseline | 32.09 (11.10) | 33.74 (10.17) | 33.76 (8.42) | 0.33 |
| Follow-up | 34.50 (11.30) | 34.69(10.17) | 34.41 (8.56) | 0.978 |
| **Sex (Male)<sup>+</sup>** |  |  |  |  |
| Baseline | 38 (39.2) | 51 (24.5) | 68 (38.9) | 0.004** |
| Follow-up | 23 (33.30) | 29 (21.5) | 22.00 (26.5) | 0.184 |
| **PHQ** |  |  |  |  |
| Baseline | 1.23 (1.82) | 12.53 (5.00) | 6.59 (5.76) | <0.001*** |
| Follow-up | 0.91 (1.73) | 8.47 (5.62) | 3.30 (4.44) | <0.001*** |
| **OASIS** |  |  |  |  |
| **Baseline** | 1.10 (1.62) | 9.65 (3.58) | 5.77 (4.46) | <0.001*** |
| Follow-up | 0.86 (1.75) | 7.11 (4.36) | 3.26 (3.73) | <0.001*** |
| **DAST-10** |  |  |  |  |
| Baseline | 0.19 (0.49) | 0.41 (0.95) | 7.46 (2.23) | <0.001*** |
| Follow-up | 0.30 (0.55) | 0.37 (0.68) | 2.27 (2.53) | <0.001*** |
| **WRAT reading score** |  |  |  |  |
| Baseline | 62.81 (5.31) | 62.75 (5.06) | 7.46 (2.23) | <0.001*** |
| Follow-up | 63.25 (5.63) | 63.02 (5.03) | 57.12 (7.12) | <0.001*** |

**Note:** PHQ = Patient Health Questionnaire; OASIS = Overall Anxiety Severity and Impairment Scale; DAST-10 = Drug Abuse Screening Test; WRAT = Wide Range Achievement Test; \**p*-values are based on ANOVAs testing for significant differences between the three groups (\*\*\**p* < 0.001; \*\**p* < 0.01; \**p* < 0.05); <sup>+</sup>Sex is reported in terms of counts and percentages. Reported *p*-values are based on Chi-Squared tests.

**Table 3.** Summary statistics and group differences for model parameters and performance.

|  | **HCS** | **DEP/ANX** | **SUDs** | ***p*\*** |
| --- | --- | --- | --- | --- |
| Baseline | N = 97 | N = 208 | N = 175 |  |
| Follow-up | N = 69 | N = 135 | N = 84 |  |
| **EC** |  |  |  |  |
| Baseline | 3.53 (3.57) | 2.97 (3.06) | 1.69 (1.94) | <0.001*** |
| Follow-up | 3.75 (3.22) | 2.90 (3.05) | 1.75 (2.10) | <0.001*** |
| **DU** |  |  |  |  |
| Baseline | 3.96 (4.65) | 4.14 (4.57) | 5.26 (5.72) | 0.048* |
| Follow-up | 3.22 (3.88) | 3.51 (4.79) | 3.63 (4.67) | 0.855 |
| **Model Accuracy** |  |  |  |  |
| Baseline | 0.77 (0.24) | 0.76 (0.24) | 0.72 (0.24) | 0.185 |
| Follow-up | 0.83 (0.22) | 0.82 (0.24) | 0.80 (0.23) | 0.697 |
| **Action Probability** |  |  |  |  |
| Baseline | 0.64 (0.29) | 0.62 (0.29) | 0.56 (0.27) | 0.019* |
| Follow-up | 0.70 (0.28) | 0.70 (0.29) | 0.67 (0.27) | 0.696 |

**Note:** *EC* = Emotion Conflict; *DU* = Decision Uncertainty; \**p*-values are based on ANOVAs testing for significant differences between the three groups. \**p* < 0.05, \*\**p* < 0.01, \*\*\**p* < 0.001.

Participant characteristics at baseline for those who did versus did not return for the follow-up visit are provided in **Supplementary Tables 2-3**. On average, participants who did not return for the follow-up visit had higher scores on the Drug Abuse Screening Test (DAST: *t*(368.68)=4.15, *p<*0.001, *d*=0.398) and lower scores on the Wide-Range Achievement Test Reading Sub-Test (WRAT), which was used to approximate pre-morbid cognitive capacity (*t*(325.23)=-2.11, *p=*0.036, *d*=-0.216; see measure descriptions below for more details). There was also a sex difference, with higher attrition in males, both overall (χ^2^(1, N=480)=14.77, *p*<0.001) and when restricting to those with SUDs (χ^2^(1, N=175)=9.17, *p*=0.002). However, males who did vs. did not return for the follow-up visit did not differ in computational measures or in anxiety, depression, or substance use severity (|*ts*|≤1.68, *ps≥*0.108). No other demographic or symptom severity differences were found between males and females or between those who did or did not return for follow-up in each clinical group separately (|*ts*|≤1.59, *ps≥*0.118).

Note that, given this differential attrition, the linear mixed effects analyses (LMEs) detailed below were performed using all participant data, including baseline data from those who did not return for follow-up. This was based on the intention-to-treat principle, with the goal of minimizing potential biasing effects of drop out on longitudinal findings (i.e., as LMEs can incorporate baseline data in model estimation despite partial missing data at follow-up). To examine trajectories of individual change, supplementary analyses were also performed when only including those who returned for the follow-up visit.

### Comparison between exploratory and confirmatory samples

Participant characteristics at follow-up in the exploratory and confirmatory samples are compared in **Table 4**. Individuals in the confirmatory sample tended to be younger (*t*(608.34)=2.05, *p*=0.041, *d*=0.165) and had lower scores on the Overall Anxiety Severity and Impairment Scale (OASIS; *t*(506.89)=2.20, *p<*0.028, *d*=0.192). When restricting analyses to the individual clinical groups, it was specifically individuals in the DEP/ANX group in the confirmatory dataset who were younger than those in the exploratory dataset (*t*(307.16) = 2.05, *p=*0.041, *d*=0.225). No other demographic differences were found (**Supplementary Table 4**).

**Table 4.** Differences in participant characteristics in the exploratory and confirmatory sample at 1-year follow-up.

| Returned for follow-up | Exploratory (N = 324) | Confirmatory (N = 287) | *p* * |
| --- | --- | --- | --- |
| Age | 36.30 (10.92) | 34.57 (9.99) | 0.041* |
| Sex (Male) | 108 (33.3) | 74 (25.8) | 0.051 |
| PHQ | 5.55 (5.96) | 5.12 (5.62) | 0.396 |
| OASIS | 5.36 (4.88) | 4.46 (4.52) | 0.028* |
| DAST-10 | 1.16 (2.03) | 0.91 (1.70) | 0.118 |
| WRAT reading score | 61.96 (5.43) | 61.41 (6.40) | 0.279 |

**Note:** PHQ = Patient Health Questionnaire; OASIS = Overall Anxiety Severity and Impairment Scale; DAST = Drug Abuse Screening Test; \**p*-values are based on a t-test for significant differences between the three groups (\**p* < 0.05, \*\**p* < 0.01, \*\*\**p* < 0.001). Sex is reported in terms of counts and percentages. Reported *p*-values are based on Chi-Squared tests.

Figure 2 illustrates the difference in model parameter values between exploratory and confirmatory samples. Here we evaluate sample differences for each group at baseline (all participant data), follow-up, and the change from baseline to follow-up (i.e., only in participants that returned for follow-up). At baseline, lower *DU* values were observed in the confirmatory sample than in the exploratory sample for both DEP/ANX (*t*(417.3)=2.86, *p*=0.004, *d*=0.27, Bayes’ Factor [BF]=5.89) and SUDs (*t*(331.86)=2.6, *p*=0.010, *d*=0.283, BF=2.92). At follow-up, HCs also had higher *DU* in the confirmatory sample when compared to the exploratory sample (*t*(99.97)=-2.05, *p*=0.043*, d*=-0.386, BF=1.30). No other sample differences were observed for *DU* or *EC* at baseline or follow-up (|*ts*|<1.13, *ps*>0.26, BFs<0.24). However, some further sample differences were identified in how parameter values changed between baseline and follow-up (see Figure 2). First, while DU values decreased from baseline to follow-up in all groups and in both samples, these decreases were stronger in the exploratory sample than in the confirmatory sample for HCs (*F*(1, 114)=6.83, *p*=0.010) and DEP/ANX (*F*(1, 324)=3.97, *p*=0.030). In contrast, SUDs showed similar decreases in DU over time in both samples.

**Figure 2.**
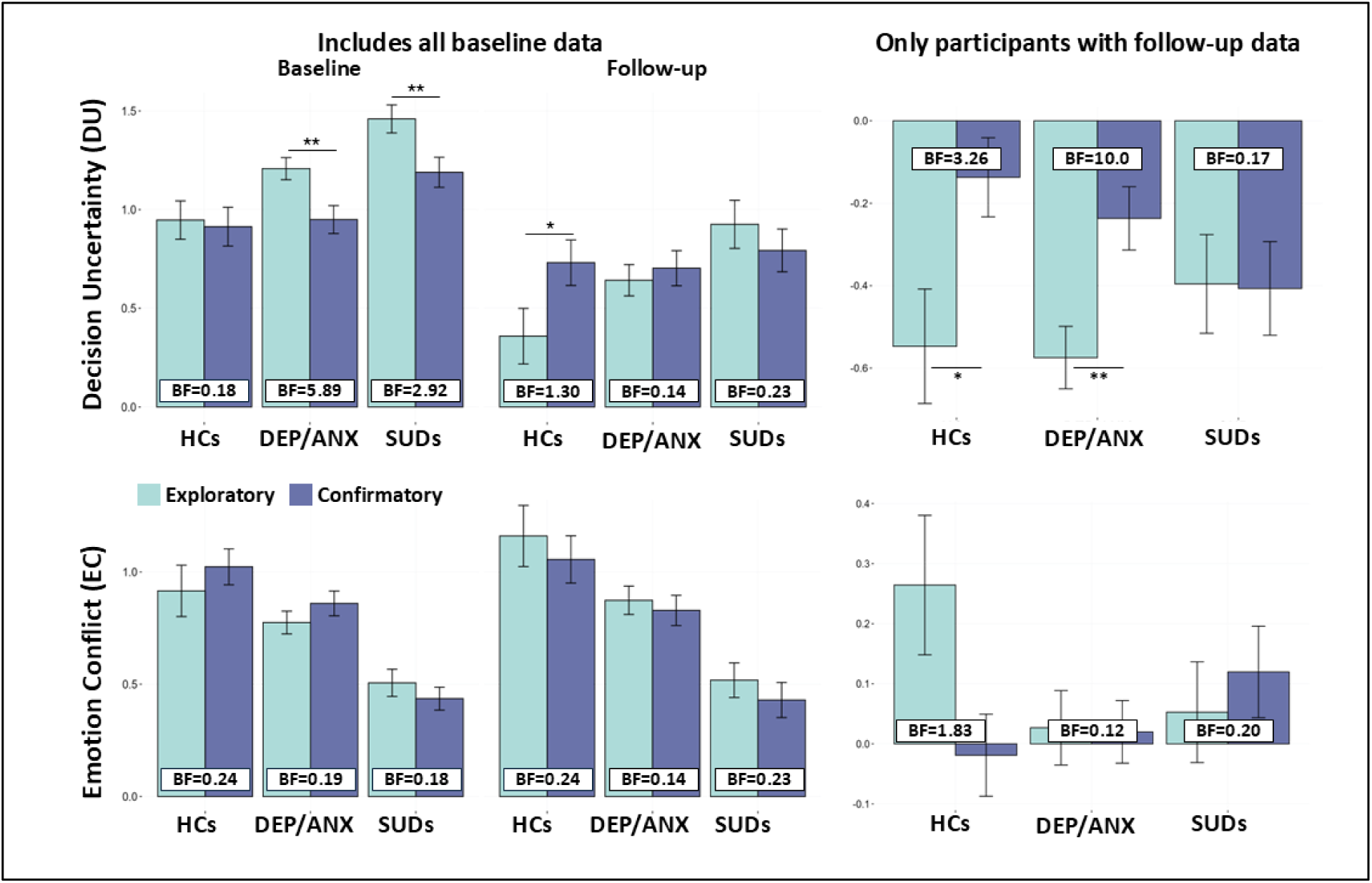
Comparison of exploratory and confirmatory samples. ***Top***: The left panels compare *DU* values between groups in each sample at baseline and follow-up. The right panel displays the associated change scores (i.e., follow-up scores minus baseline scores). Here, DEP/ANX and SUDs showed lower *DU* in the confirmatory sample compared to the exploratory sample at baseline, while HCs showed higher *DU* in the confirmatory sample compared to the exploratory sample at follow-up. Reductions in *DU* over time were greater in the exploratory sample than in the confirmatory sample for HCs and DEP/ANX, but not for SUDs. ***Bottom***: The left and right panels show equivalent group by sample comparisons for *EC* scores and change score comparisons, respectively. No sample differences in *EC* were observed for any groups at baseline or follow-up. HCs in the exploratory sample showed an increase in *EC* from baseline to follow-up, while participants in the confirmatory sample instead showed no change in *EC* values. \**p* < 0.05, \*\**p* < 0.01, \*\*\**p* < 0.001.

### Intra-class correlations between parameters at baseline and 1-year follow-up

Between baseline and 1-year follow-up, ICCs for *EC* and *DU* in the confirmatory sample were fair-to-good (ICCs=0.70 and 0.57, respectively). ICCs for response times (RTs) were fair across task conditions (ICCs between 0.43 and 0.51). For descriptive choice measures, ICCs across conditions were fair-to-good for average chosen runway position (ICCs between 0.54 and 0.71) and poor-to-fair for choice variability (ICCs between 0.37 and 0.51). These results were consistent with those in the exploratory sample. Consistency in clinical scale scores and dimensional measures was also fair-to-good across all participants (ICCs between 0.42 and 0.74). Detailed results of ICCs by group and condition for task measures, as well as for clinical scales and post-task surveys, are provided in **Supplementary Tables 5-6**.

### Model validation

Across all participants, the model accurately predicted behavior in 81.35% (SD = 1.4%) of trials (i.e., chance accuracy = 11%; 1/9 runway positions) with an average action probability of 0.69±0.28. There were no group differences in model performance (**Table 3**). As expected, higher values for each parameter predicted slower RTs for each task condition (*DU*: *r*s=0.31-0.55, *p*s<0.001; *EC*: *r*s=|0.14-0.30|, *p*s<0.024); see **Supplementary Figure 2**. This is consistent with results at baseline and with prior longitudinal results in the exploratory sample (Smith et al., 2021b; Smith et al., 2021c; Smith et al., 2023). Relationships between model parameters and post-task survey questions were also in expected directions, as detailed in **Table 5**. In particular, there were significant positive associations between *EC* and both self-reported anxiety (Q2) and avoidance motivation (Q5) during the task, and a positive association between *DU* and greater self-reported decision difficulty (Q3) on the task (**Supplementary Figure 3**). As the model was not fit to these RT and self-report measures, this provides external support for its validity. Supplementary within-group analyses for these measures are reported in **Supplementary Table 7**.

**Table 5.** Post-task self-report questionnaire items at baseline and follow-up, and correlations with computational model parameters at follow-up.

| Post-Task Self-Report Questions (Likert Scale: 1 = not at all; 7 = very much) | Mean (SD) Baseline |  | Mean (SD) 1-Year follow-up (N=287) | Emotion conflict parameter (EC) | Decision uncertainty parameter (DU) |
| --- | --- | --- | --- | --- | --- |
|  | All participants (N=480) | Returned for follow-up (N=287) |  |  |  |
| Q1. I found the positive pictures enjoyable | 5.02 (1.56) | 5.17 (1.56) | 4.97 (1.59) | 0.06 | -0.04 |
| Q2. The negative pictures made me feel anxious or uncomfortable | 4.01 (1.97) | 3.98 (1.99) | 4.00 (1.95) | **0.34\*\*\*** | **0.06** |
| Q3. I often found it difficult to decide which outcome I wanted | **2.32 (1.71)** | **2.26 (1.68)** | **2.07 (1.60)** | 0.04 | **0.42\*\*\*** |
| Q4. I always tried to move all the way towards the outcome with the largest reward points | 4.86 (2.35) | 4.84 (2.38)<sup>+</sup> | 4.98 (2.44) | **-0.77\*\*\*** | **-0.46\*\*\*** |
| Q5. I always tried to move all the way away from the outcome with the negative picture/sounds | 2.86 (2.15) | 2.92 (2.19) | 2.93 (2.32) | **0.72\*\*\*** | **0.31\*\*\*** |
| Q6. When a negative picture and sound were displayed, I kept my eyes open and looked at the picture | **5.43 (1.89)** | **5.39 (1.89)<sup>+</sup>** | **5.29 (1.97)** | **-0.42\*\*\*** | **-0.21\*\*\*** |
| Q7. When a negative picture and sound were displayed, I tried to think about something unrelated to the picture to distract myself | **2.84 (1.90)** | **2.93 (1.95)** | **2.95 (1.97)** | **0.32\*\*\*** | 0.04 |
| Q8. When a negative picture and sound were displayed, I tried other strategies to manage emotions triggered by the pictures | **3.04 (1.91)** | **3.11 (1.90)** | **3.22 (1.93)** | **0.34\*\*\*** | **0.05** |

**Note:** <sup>+</sup> $p < 0.05$ , <sup>++</sup> $p < 0.01$ , <sup>+++</sup> $p < 0.001$ (pre-post differences); \* $p < 0.05$ , \*\* $p < 0.01$ \*\*\* $p < 0.001$ (correlations at follow-up). Results that were also statistically significant in the exploratory sample are highlighted in bold.

### Diagnostic effects over time

To evaluate stability in model parameters, LMEs predicting each parameter were estimated including main effects of age, sex, group, and time, as well as interactions between group and sex and group and age. The LME predicting *DU* showed significant main effects of age (*F*(1, 462)=28.76, *p*<0.001), time (*F*(1, 346)=32.83, *p<*0.001), and group (*F*(2, 469)=3.08, *p=*0.047). This indicated an increase in *DU* with age, reduction in *DU* over time (EMM_Baseline_=1.03; EMM_Follow-up_=0.73; *t*(346.52)=5.73, *p*<0.001 *d*=0.46), and greater *DU* in SUDs (EMM=1.05) than the DEP/ANX group (EMM_DEP/ANX_=0.84; *t*(472.34)=-2.23, *p*=0.026, *d*=0.33). In the LME predicting *EC*, there was a main effect of age (positive association between age and *EC*; *F*(1, 462)=28.76, *p*<0.001) and sex (greater *EC* in females than males; *F*(1, 471)=6.24, *p*=0.013; EMM_Females_=0.80; EMM_Males_=0.66; *t*(475.87)=2.03, *p*=0.043, *d*=0.34). There was also a main effect of group (*F*(2, 470)=15.86, *p*<0.001), reflecting greater values in HCs (EMM=1.07) than the clinical groups (DEP/ANX: EMM =0.85; *t*(448.69)=-2.468, *p*=0.014, *d*=-0.49; SUDs: EMM=0.44; *t*(458)=6.793, *p*<0.001, *d*=1.41). Further, SUDs also had significantly lower *EC* than DEP/ANX (*t*(473.21)=5.44, *p*<0.001, *d*=0.92). A *group x sex* interaction (*F*(2, 472)=5.41, *p*=0.048) was also present. The interaction indicated that the group effect was primarily driven by females (Females: EMM_HCs_=1.21; EMM_DEP/ANX_=0.90; EMM_SUDs_=0.41; *t*s≥2.89, *p*s≤0.004; and Males: EMM_HCs_=0.72; EMM_DEP/ANX_=0.72; EMM_SUDs_=0.53; *t*s≤1.40, *p*s≥0.16; see Figure 3).

**Figure 3.**
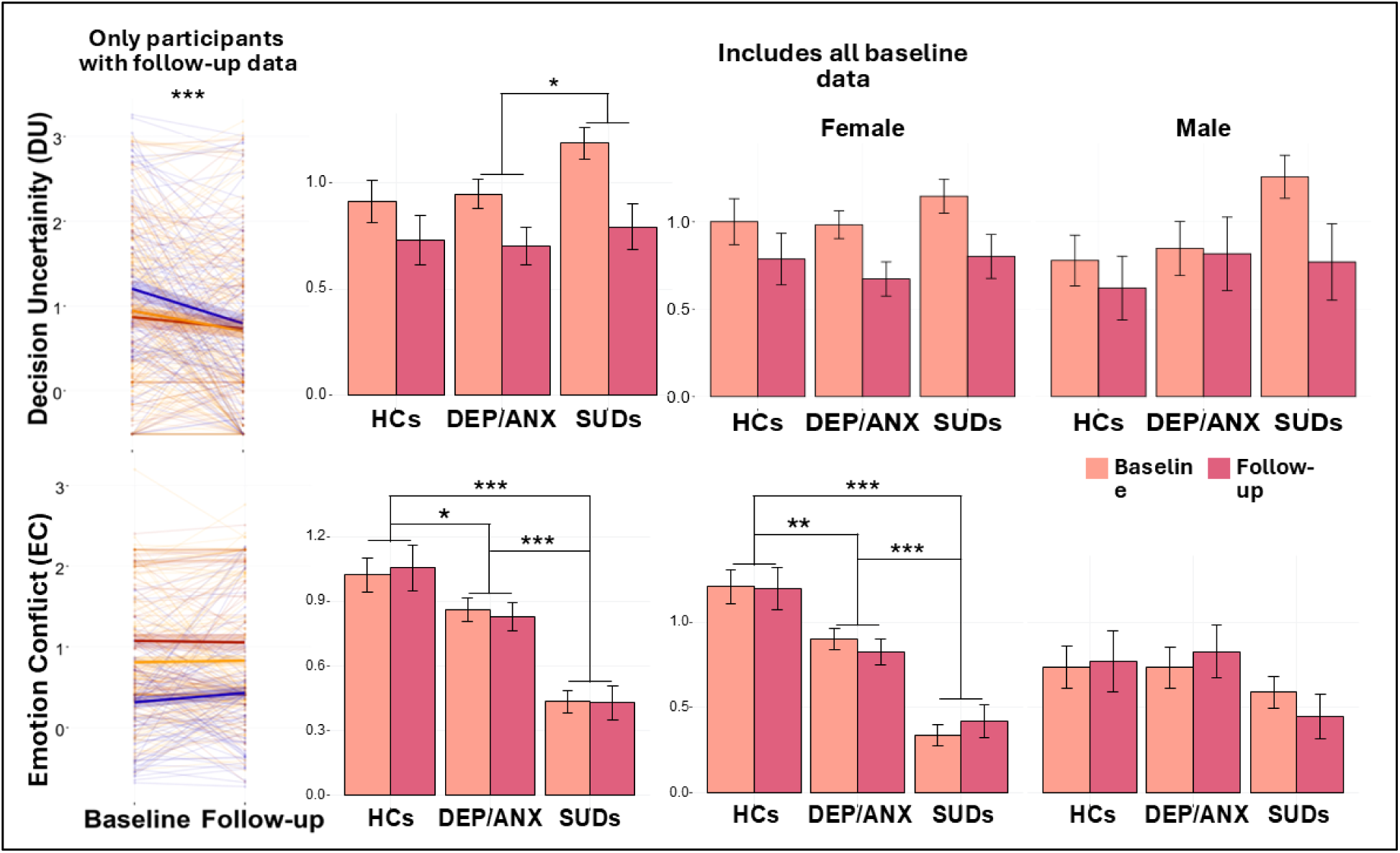
Effects of group, time, and sex on model parameters. Spaghetti plots (left) show change from baseline to follow-up and therefore only include participants who returned for follow-up. Bar plots (right) include all baseline participants, including those who did not return for follow-up. Spaghetti plots illustrate individual differences in the stability of parameter values over time. Here, thick lines indicate group means and lighter lines indicate individual values. The shaded area around each group mean line reflects the standard error. Sex comparisons in the lower bar graphs illustrate how observed group differences in *EC* were mainly driven by females. \**p* < 0.05, \*\**p* < 0.01, \*\*\**p* < 0.001.

These effects largely replicated prior results (replication summary provided in **Supplementary Figure 4**). Similar results were also found in pre-registered LMEs including only effects of group and time, and in further LMEs controlling for possible effects of premorbid cognitive ability (WRAT reading scores) in those with available data (**Supplementary Tables 8-9**). Analogous LMEs were also performed when only including data from participants who returned for the follow-up visit, with similar results (**Supplementary Figure 5**).

Identical analyses were also performed for the descriptive task measures: RTs, average chosen runway position, and choice variability (**Supplementary Tables 10-21; Supplementary Figure 6**). These results also replicated many of our previously reported findings (detailed in **Supplementary Figure 7).**

In contrast, when testing the ability of baseline model parameters or other behavioral task measures to predict change in symptoms over time, most previously reported findings did not replicate (**Supplementary Figure 1**). The only replicable finding was the negative relationship between average chosen runway position in the CONF2 condition at baseline and change in self-reported Behavioral Inhibition System (BIS) scores at follow-up (*r*=-0.14; *p*=0.047). Other significant associations were observed (i.e., present in the confirmatory sample but not in the exploratory sample), however, suggesting the presence of either false positives or false negatives in either sample. Lastly, LMEs were also performed for post-task self-report survey items assessing anxiety (Q2), decision-making difficulty (Q3), approach motivation (Q4) and avoidance motivation (Q5; **Supplementary Tables 22-24**). Results replicated previous findings in some cases, but not others (for details, see **Supplementary Figure 8**). Successful replications included main effects of group on approach (*F*(2, 468)=4.87, *p* = 0.008) and avoidance (*F*(2, 467)=4.18, *p*=0.016) motivation, where SUDs (EMM_Approach_=5.44; EMM_Avoid_=2.39) displayed greater approach motivation (EMM_SUDs_=5.44; EMM_HCs_=4.32; *t*(452.58)=-4.03, *p*<0.001, *d*=-0.73) and reduced avoidance motivation (EMM_SUDs_=2.34; EMM_HCs_=3.31; *t*(448.86)=3.81, *p*<0.001, *d*=0.63) compared to HCs. The previously observed *group x sex* interaction (*F*(2, 469)=4.32, *p*=0.014) in avoidance motivation was also observed here, suggesting that the lower avoidance motivation observed in SUDs (EMM_Female_=2.20; EMM_Male_=2.67) than HCs (EMM_Female_=3.64; EMM_Male_=2.56) was driven by females (Females: *t*(438.38)=4.49, *p*<0.001, *d*=0.93; Males: *t*(486.53)=-0.254, *p*=0.799, *d*=-0.07).

### Evaluation of diagnostic specificity

After carrying out all pre-registered analyses above, we then combined exploratory (N_Baseline_=478; N_Follow-up_=324) and confirmatory (N_Baseline_=480; N_Follow-up_=287) samples. This provided greater power to examine whether observed group differences were consistent across the more specific diagnostic categories present within the dataset (e.g., generalized anxiety disorder, social anxiety disorder, methamphetamine use disorder, opioid use disorder, etc.). To answer this question, separate LMEs were performed including only HCs and different subsets of participants from the combined clinical groups. Namely, one LME was performed including the subset of participants who met criteria for each specific disorder (e.g., all those with stimulant [cocaine or methamphetamine] use disorder, or all those with major depressive disorder, etc.). Note that, due to comorbidities, these subsets of participants had partial overlap (e.g. individuals with major depressive disorder may or may not have had other anxiety or substance use disorders). These LMEs then predicted each model parameter based on the specific diagnostic category in question (e.g., HCs versus generalized anxiety, HCs versus opioid use disorder, etc.); effects of time and the interaction term were also included. As a secondary check, these LMEs were also re-performed when including possible effects of age, sex, and their interaction with group. Here we found consistent main effects of group across specific disorders for each parameter (Figures 4-5). There were no effects of time, with the exception of increases in *EC* over time in those with sedative use disorders (EMM_Baseline_=0.78; EMM_Follow-up_=0.90) and major depression (EMM_Baseline_=0.76; EMM_Follow-up_=0.83). Overall, these results indicated that HCs had greater *EC* values than each specific diagnostic group (*F*s≥8.03, *p*s≤0.002). When accounting for effects of age and sex, these main effects of group remained in all cases (*F*s≥6.41, *p*s≤0.012). Females also displayed greater avoidance than males across all specific diagnostic groups (*F*s≥17.41, *p*s<0.001). Extending prior results, a significant *group x sex* interaction (*F*s≥5.16, *p*s<0.024) was also found in most cases. Specifically, the group differences between HCs and those with major depression, generalized anxiety, panic, cannabis, stimulant, and opioid use disorders were each driven by females (*t*s≥4.22, *p*s<0.001). In alcohol (*F*(1, 270) = 9.57, *p*=0.002) and sedative (*F*(1, 244)=8.20, *p*=0.005) use disorder, the group difference was present in both sexes, but the effect in females remained stronger (alcohol: *t*(268.68)=6.38, *p*<0.001, *d*=1.81; sedative: *t*(237.21)=6.97, *p*<0.001, *d*=1.89) than in males (alcohol: *t*(274.97)=2.37, *p*=0.018, *d*=0.62; sedative: *t*(250.48)=2.18, *p*=0.03, *d*=0.70).

**Figure 4.**
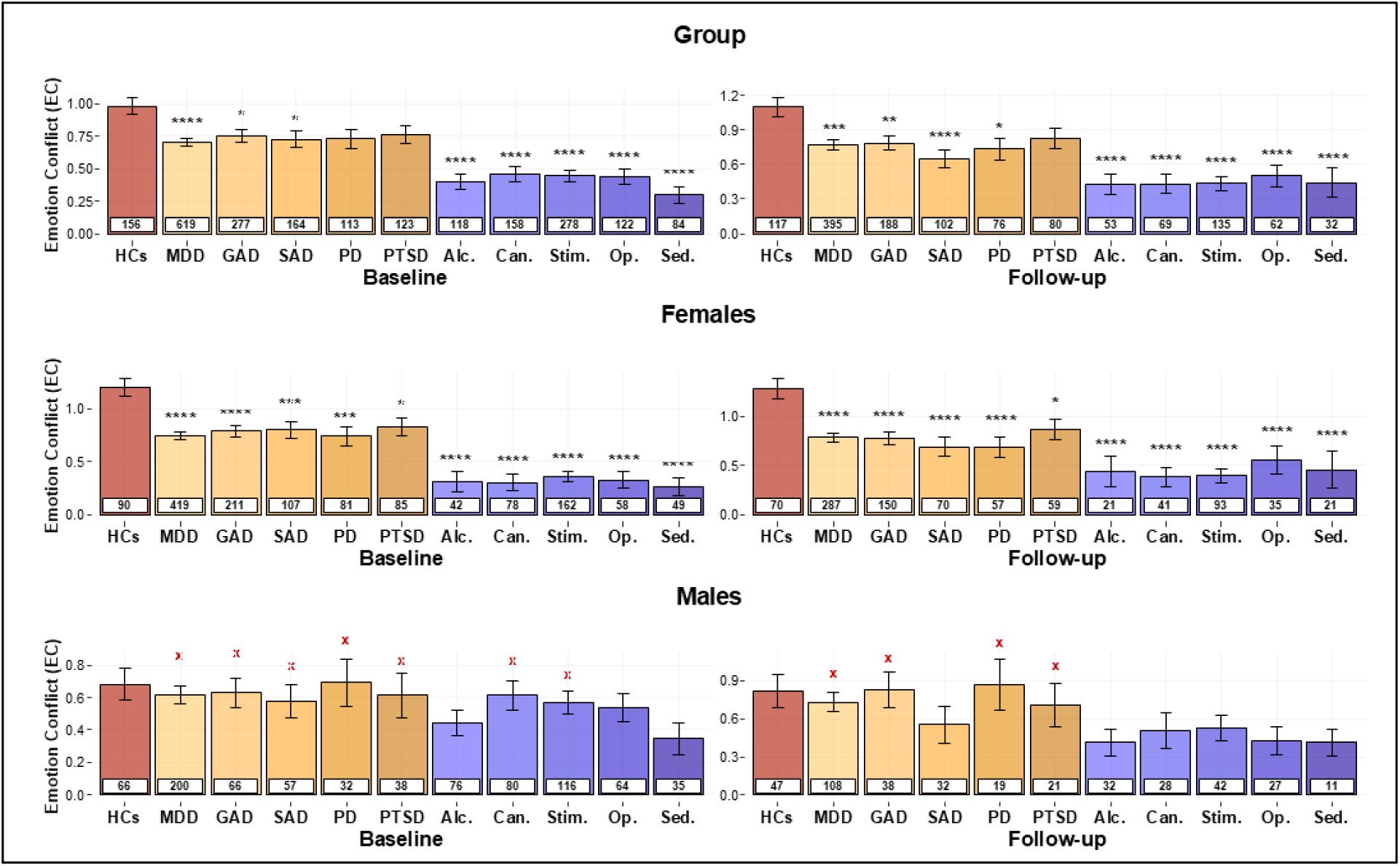
Effects of group and sex on emotion conflict (EC) across specific diagnostic categories in the combined sample. Values are shown separately for healthy comparisons (HCs) and each specific affective and substance use disorders. For each group, the number of participants is provided in the white box. **Top**: Differences between HCs and each specific diagnosis are consistent with differences seen in the broader clinical groupings. **Middle**: The overall group differences appear to be driven by females at both baseline and follow-up. **Bottom**: Males with affective and substance use disorders did not differ from HCs. MDD: Major Depressive Disorder; GAD: Generalized Anxiety Disorder; SAD: Social Anxiety Disorder; PD: Panic Disorder; PTSD: Post-Traumatic Stress Disorder; Alc.: Alcohol; Can.: Cannabis; Stim.: Stimulant; Op.: Opioid; Sed.: Sedative. For illustrative purposes, Bayesian t-tests were performed to evaluate evidence for the presence or absence of group differences with HCs at each time point. Bayes Factors (BFs) greater than 3 were taken to support the presence of an effect, denoted by black stars (*): *BF>3; **BF>10, ***BF>30, ****BF>100. BFs between 0.33 and 3 were considered equivocal null results. BFs less than 0.33 were taken to support the absence of a group difference with HCs, denoted by red Xs: ^x^BF<0.33, ^xx^BF<0.1, ^xxx^BF<0.033, ^xxxx^BF<0.001.

**Figure 5.**
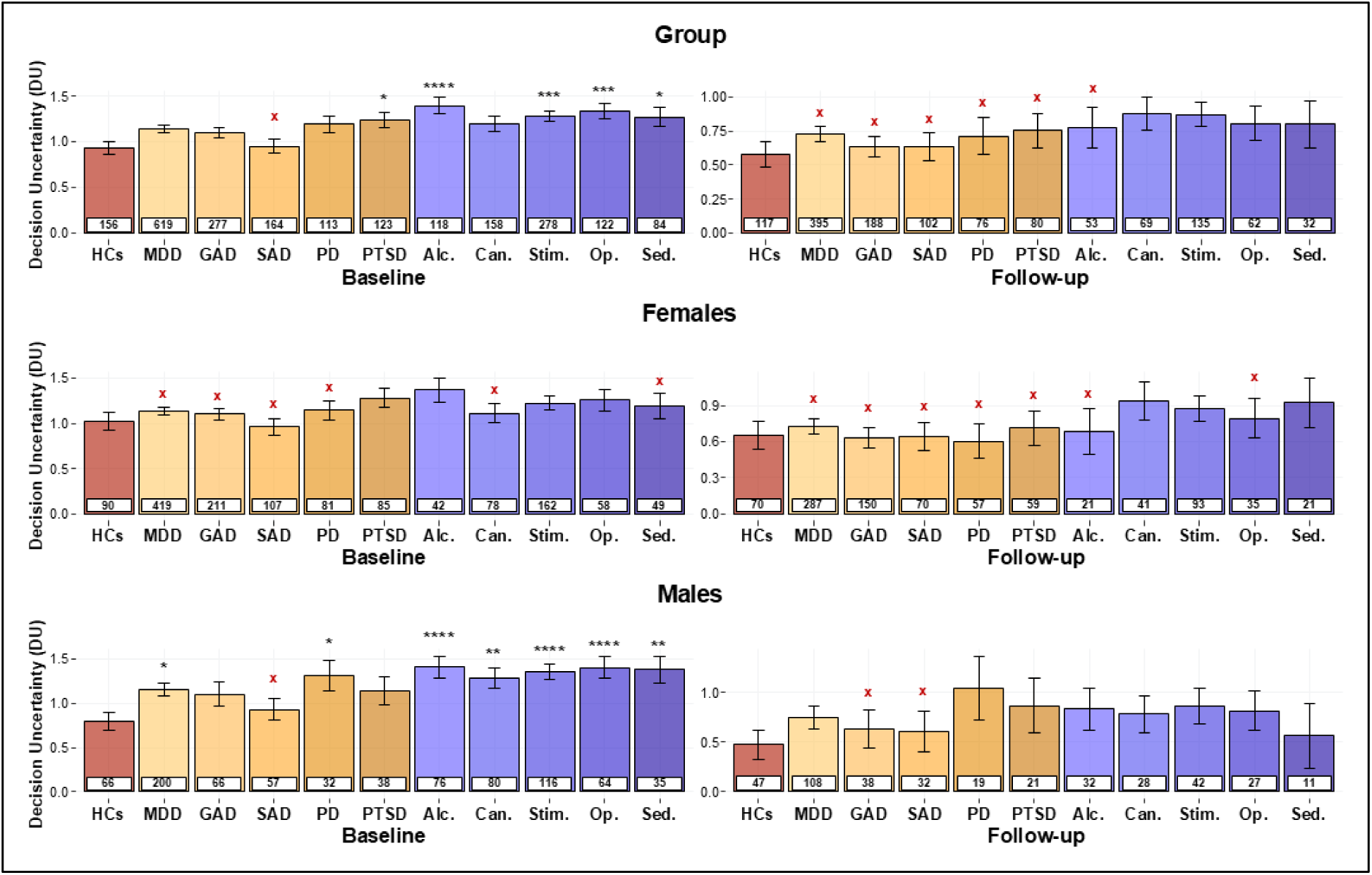
Effects of group and sex on decision uncertainty (*DU*) across specific diagnostic categories in the combined sample. Values are shown separately for healthy comparisons (HCs) and each specific affective and substance use disorders. For each group, the number of participants is provided in the white box. ***Top***: Group differences (between HCs and each specific diagnosis) were consistent with differences seen in DEP/ANX and SUDs more broadly. ***Middle*** and ***Bottom***: Male but not female SUDs displayed greater *DU* at baseline. MDD: Major Depressive Disorder; GAD: Generalized Anxiety Disorder; SAD: Social Anxiety Disorder; PD: Panic Disorder; PTSD: Post-Traumatic Stress Disorder; Alc.: Alcohol; Can.: Cannabis; Stim.: Stimulant; Op.: Opioid; Sed.: Sedative. For illustrative purposes, Bayesian t-tests were performed to evaluate evidence for the presence or absence of group differences with HCs at each time point. Bayes Factors (BFs) greater than 3 were taken to support the presence of an effect, denoted by black stars (*): *BF>3; **BF>10, ***BF>30, ****BF>100. BFs between 0.33 and 3 were considered equivocal null results. BFs less than 0.33 were taken to support the absence of a group difference with HCs, denoted by red Xs: ^x^BF<0.33, ^xx^BF<0.1, ^xxx^BF<0.033, ^xxxx^BF<0.001.

Group differences were also found in *DU*, whereby HCs had lower *DU* than most sub-groups (*F*s≥4.08, *p*s≤0.044), with the exception of generalized and social anxiety disorders, and a similar but non-significant pattern in panic disorder (*F*(1, 271)=3.71, *p*=0.055). A main effect of time (*F*s≥19.80, *p*s<0.001) was also found, with lower *DU* at follow-up in all sub-samples.

Comparable results were found when accounting for effects of age, sex, and their interaction with group. There was also a *group x age* interaction (*F*s≥6.52, *p*s≤0.011) in all groups with affective disorders, with the exception of individuals with PTSD (*F*(1, 274)=3.58, *p*=0.059). This interaction reflected a positive association between age and *DU* in affective disorders (depression, generalized anxiety, social anxiety and panic disorders; |*ts*|≥2.55, *ps*≤0.011) but not in HCs. No other significant effects were found. Notably, our previous paper reported that *DU* (but not *EC*) differentiated social anxiety from other affective disorders at baseline within a logistic regression model (Smith et al., 2023). As a supplementary test, we repeated this analysis in the follow-up data here to confirm whether this difference was stable over time (i.e., including the effects of *DU* and *EC*, along with their interaction with age and sex). This revealed a significant effect of age (Wald *z*=−3.7, *p*<0.001), but no other main effects or interactions were present (Wald *z*=−1.68 to 1.60, *p*s>0.093).

Descriptive statistics for model parameters in each specific diagnostic group are provided in **Supplementary Table 25**. Detailed results of the LMEs are provided in the **Supplementary Tables 26-29**.

### Model-based trial-by-trial response time prediction

As expected, trial-by-trial *choice uncertainty* predicted longer RTs across participants (Intercept: *F*(1,916)=441.48, *p*<0.001; **Supplementary Figure 9**). Notably, this association was significantly higher in SUDs (EMM=0.16) than in both DEP/ANX (EMM=0.11; *t*(943.433)=-4.67, *p*<0.001, *d*=-0.32) and HCs (EMM=0.11; *t*(898.45)=-4.12, *p*<0.001, *d*=-0.38).

These results prompted further post-hoc analyses to test for consistent group differences in the relationship between RTs and both choice behavior (average chosen runway position; **Supplementary Figure 10**) and model parameters. Here, LMEs showed a significant *group x RT* interaction in predicting choice of runway position (*F*(2, 1547.32) = 4.78, *p* = 0.009; ET_HCs_=-0.59; ET_DEP/ANX_=-1.34; ET_SUDs_=-1.94) and *EC* (*F*(2, 1554.12)=3.48, *p*=0.031; ET_HCs_=0.17; ET_DEP/ANX_=0.46; ET_SUDs_=0.69), but not *DU* (*F*(2, 1462.84)=0.29, *p*=0.752). Both significant interactions suggested that shorter RTs were associated with greater approach behavior to a greater extent in SUDs than HCs (choice: *t*(1533)=3.13, *p*=0.005; *EC*: *t*(1556)=-2.64, *p*=0.023).

### Predictive categorization in the combined sample

Our predictive (out-of-sample) categorization analyses using the stacked machine learning approach (Kuhn, 2008) first restricted the sample to individuals with one or more clinical diagnoses (i.e., removing HCs) and then classified individuals (Figure 6) in terms of those with affective disorders but no SUDs vs. SUDs but no affective disorders. Here we found that ENET showed the highest balanced accuracy (0.71; Sensitivity=0.76; Specificity=0.67) when model parameters (*EC* and *DU* at baseline and follow-up), age and sex were used as predictors. In comparison, when only age and sex were used as predictors, ENET showed a lower balanced accuracy of 0.47, comparable to a neutral threshold of 0.5 (Sensitivity=0.30; Specificity=0.65). Thus, adding model-based information led to improved out-of-sample classification. When further evaluating AUCs as a means of quantifying the ability of the model to distinguish between groups, we found an acceptable discrimination ability of 0.77, compared to poor discrimination with age and sex alone (AUC=0.50). In addition, the exclusion of either *DU* (balanced accuracy=0.59; AUC=0.65) or *EC* (balanced accuracy=0.51; AUC=0.52) from the best models caused a substantial reduction in model performance. Performance metrics for all predictive models are provided in **Supplementary Table 30**.

**Figure 6.**
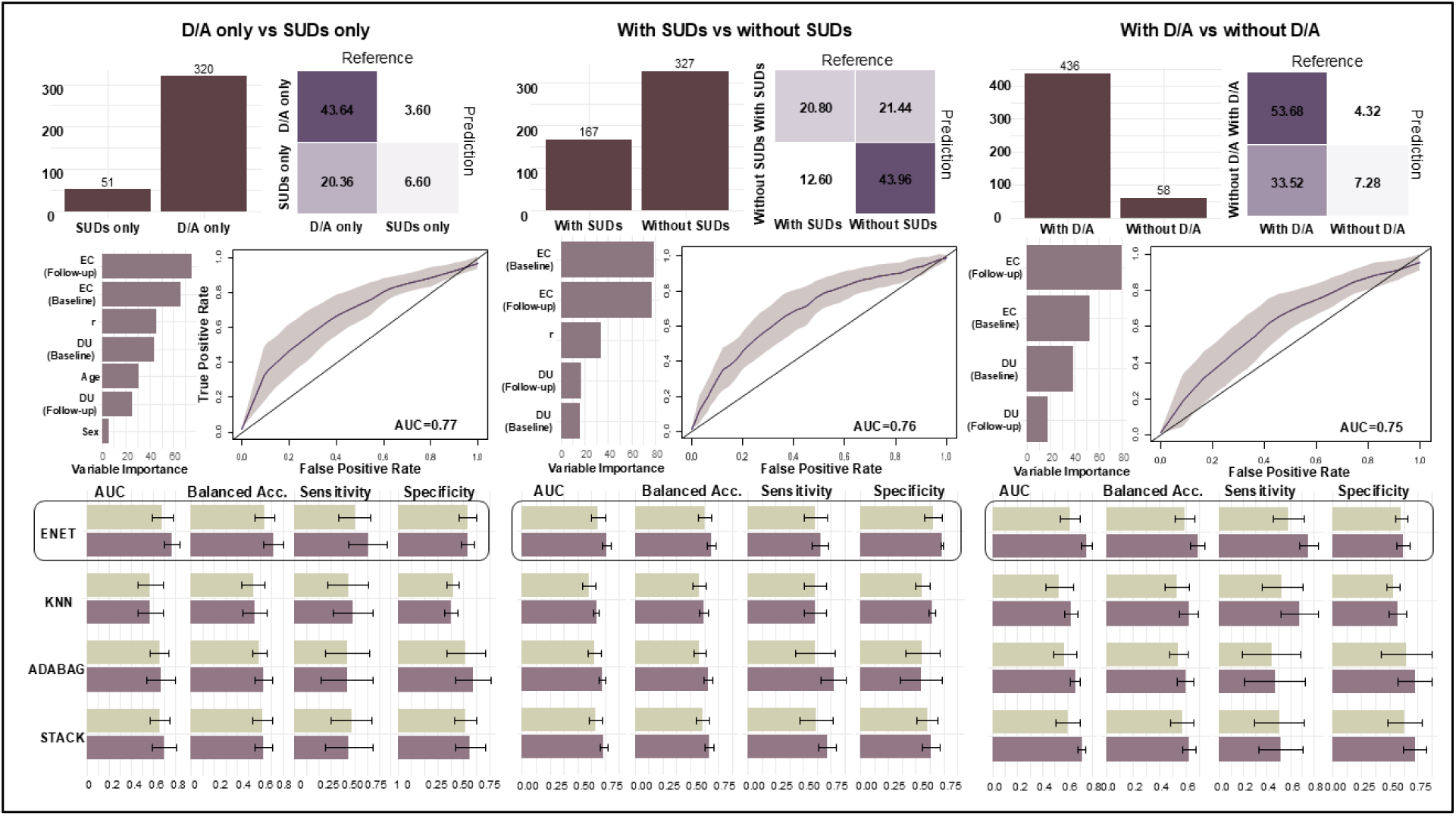
Predictive classification in the clinical sample. In the combined clinical sample (excluding HCs), multiple stacked classification algorithms predictively classified (*left panel*) individuals with only affective disorders (D/A) vs. only substance use disorder (SUDs), (*middle panel*) individuals with vs. without SUDs, and (*right panel*) individuals with vs. without D/A. ***Top-left***: Histograms show the number of participants in each group. Up-sampling was used to minimize model biases. ***Top-right***: Confusion matrices illustrate classifications accuracy for the winning model. ***Middle-left***: Bar plots indicate variable importance (VI), which is the relative contribution each predictor used in the winning model. This metric was computed using the STACK approach to reflect the contribution of predictors across algorithms. ***Middle-right***: Receiver operating characteristic curves (ROCs) are shown for the winning model. ROCs provide information about true positive and false positive rates at various categorization thresholds. Associated area-under-the-curve (AUC) values show the probability of a sample being assigned to the correct group. ***Bottom***: Model performance was evaluated using AUCs, balanced accuracy, sensitivity (true positive rate), and specificity (true negative rate). Balanced accuracy was used due to unbalanced sample sizes. Performance metrics were averaged across the five folds of cross validations. ENET had the highest balanced accuracy (highlighted) for each predictive classification and was chosen as the best performing algorithm. Algorithms: ENET = elastic net; KNN = k-nearest neighbors; ADABAG = Bagged AdaBoost.

The same approach was then repeated (still excluding HCs) to classify: 1) the presence vs. absence of SUDs (**Supplementary Table 31**), 2) the presence vs. absence of affective disorders (**Supplementary Table 33**), and 3) comorbid affective and substance use disorders vs. the presence of either disorder alone (**Supplementary Table 33**). In each case, ENET similarly showed the best performance, where incorporating model-based information led to improvement from chance (with age and sex alone) to above-chance balanced accuracy (0.69,0.69 and 0.64, respectively) and discrimination ability (AUCs: 0.76, 0.75 and 0.68, respectively). Exclusion of *DU* from the best models caused a reduction in balanced accuracy (0.61, 0.58 and 0.60, respectively) and discrimination ability (AUC: 0.64, 0.64 and 0.63, respectively). Similar reductions in balanced accuracy (0.60, 0.59 and 0.63, respectively) and discrimination ability (AUC: 0.63, 0.60 and 0.66, respectively) were found when *EC* was instead excluded from the best models.

When examining variable importance across the three groupings described above, *EC* at baseline (VI≥59.97) and follow-up (VI≥34.69) had the highest relative contribution in all cases when compared to *DU* at baseline (16.21≥ VI) or follow-up (29.30≥VI). However, as SUDs showed greater *DU* and lower *EC* than DEP/ANX in other results above, both parameters appear relevant for clinical differentiation.

## Discussion

The current study was designed to achieve two main goals. First, as pre-registered, we replicated a longitudinal computational modeling study in a new sample of individuals with depression/anxiety (DEP/ANX), substance use disorders (SUDs), and healthy comparisons (HCs) who completed an approach-avoidance conflict (AAC) task. Second, we combined the previous and new samples of participant data to answer more granular questions about how computational mechanisms may differentiate specific clinical disorders. Namely, we tested potential differentiations between HCs and each psychiatric disorder present in the sample. This was followed by a machine learning-based (out-of-sample) classification approach to evaluate whether computational modeling measures could predictively identify individuals as having affective or substance use disorders (or comorbidity of the two). Pre-registered replication analyses confirmed that some, but not all, prior findings (Smith et al., 2021c) were present in the new sample. Most centrally, the greater emotion conflict (*EC*; tracking avoidance motivation) values previously seen in HCs compared to the clinical groups showed longitudinal stability, most starkly in females and in those with SUDs. In contrast, decision uncertainty (*DU*) tended to reduce over time and group estimates converged to non-significant differences at 1-year follow-up. There was also a general pattern wherein specific diagnoses tended to differ from HCs in the same direction as the broader DEP/ANX and SUD groupings, suggesting transdiagnostic relevance. At the same time, model-based measures could also predictively classify whether individuals had DEP/ANX or an SUD, suggesting that some computational decision mechanisms may also differ between diagnoses. We discuss each of these findings below.

In our initial exploratory studies, the pattern of group differences in *EC* was surprising, as maladaptive avoidance is often observed clinically in anxiety and depression. However, given the confirmation of this result, both over time and in independent samples, it appears that higher *EC* may instead reflect adaptive responding, particularly in females. One possibility is that this could reflect a healthy regulation strategy in which anticipated emotional states are more strongly prioritized over small task rewards. If so, results might suggest that those with SUDs, and to a weaker extent DEP/ANX, may not prioritize their emotional states in this same way. A related factor could be that individuals in the clinical groups have less awareness or trust in their bodily/emotional signals (e.g., as suggested by Lavalley et al., 2024; Smith et al., 2021a; Smith et al., 2020), and/or that their elevated baseline negative affect attenuates the impact of unpleasant task stimuli during choice. Another possibility is that lower *EC* stems from blunted responses to negative stimuli, as suggested in some previous work in SUDs (Weinberg et al., 2016). This would be congruent with the broader SUD literature describing reduced sensitivity to punishments (Simons & Arens, 2007; Simons et al., 2008) as well as affective stimuli more generally (Hester et al., 2013; Stewart et al., 2014). Given that individuals with SUDs also reported greater approach motivation on the task, these findings could index heightened motivation to seek immediate rewards, which may outweigh the expected negative consequences of substance use (such as loss of career or social support).

The consistent sex differences we observe, in which it was primarily female HCs that showed greater avoidance than those in the clinical groups, may also relate to other known sex differences in affect and decision making. For example, studies have shown that females rely more on avoidant coping mechanisms in general, which in turn positively relates to anxiety (Panayiotou et al., 2017; Sinha & Latha, 2018). However, it should be kept in mind that we found proportionally higher dropout in males over time, which could have influenced results. On the other hand, there were no differences identified between males who did versus did not return for follow-up (e.g., in affective or substance use symptom severity) that would raise concerns in this regard.

Several novel results in the combined samples also offered insights of potential clinical relevance. For example, we evaluated the utility of our computational model to capture the deliberative aspects of decision-making captured by response times (RTs) and tested for evidence of associated clinical differences. Here we found that correlations between trial-by-trial choice uncertainty and RTs were significantly positive across all participants, but also that this relationship was stronger in SUDs than the other groups, suggesting potential differences in the deliberation process matching the observed choice differences. This was further supported by a stronger inverse relationship between RTs and approach behavior in SUDs relative to HCs (i.e., based on both average runway position and *EC*). This relationship to longer RTs appears congruent with the greater *DU* values found in SUDs, both suggesting a greater contribution of uncertainty in reaching approach decisions (recall that SUDs also showed greater RTs than the other groups in general). Machine learning analyses combining RT-uncertainty correlation values with model parameters (at both time points) could also discriminate between individuals who had only DEP/ANX from those with only SUDs in an out-of-sample manner (balanced accuracy = 0.71). Similarly, this machine learning approach could predict the presence or absence of SUDs (balanced accuracy = 0.69), and the presence or absence of DEP/ANX (balanced accuracy = 0.69) irrespective of comorbidities, as well as classify those with or without comorbidity in general (i.e., between affective and substance use disorders; balanced accuracy = 0.64), potentially reflecting overall illness severity. Thus, overall, the stability of these maladaptive choice patterns over time, and potential differences in deliberative processes, may go beyond the general presence of psychopathology. Instead, it appears they also differentiate between disorders both with and without comorbidity.

Before concluding, it is important to emphasize remaining limitations. First, as mentioned above, the sample was imbalanced with respect to clinical group membership and sex ratio within each group. There was also differential drop-out over time. That said, our intent-to-treat analysis approach aimed to minimize biasing effects of drop-out, and machine learning analyses made use of balanced accuracy metrics designed to compensate for imbalanced sample sizes. Next, as our community sample was not intentionally recruited to test the presence versus absence of some psychiatric comorbidities (e.g., generalized anxiety disorder without major depression), future work will be necessary to compare specific affective or substance use diagnoses more definitively. Finally, it should be noted that other modeling approaches could have been used. However, we followed prior work (Smith et al., 2021b; Smith et al., 2021c; Smith et al., 2023) and a pre-registered approach, and the model appeared to perform well in capturing decision dynamics.

In summary, these findings contribute to ongoing efforts to replicate and confirm the generalizability of prior results. They also extend the utility of previously observed transdiagnostic markers by demonstrating their further ability to predictively categorize different clinical groups. Not all results replicated, further supporting the importance of such confirmatory studies. In particular, the ability of the model parameters to predict symptom changes over time did not appear consistent between studies. In contrast, the results that did replicate, as well as the novel evidence supporting the potential utility of *EC* and *DU* as diagnostic predictors, should now motivate future work aimed at intervening on these mechanisms and evaluate their causal relevance. It should also motivate further work aimed at better understanding underlying neural mechanisms and potential relationships with changes in symptom severity.

## Author Contributions

M.M.M. and R.S. led the formal analysis and manuscript preparation. N.H contributed to the formal analysis. R.A. led task development and M.P.P. led the larger funded study in which these data were collected. All authors reviewed, revised, and approved the final manuscript.

## Data Availability

All data produced in the present study are available upon reasonable request to the authors.

## Supplementary Materials

### Intraclass correlations in model parameters by group

ICCs for *DU* by group were as follows:

**HCs**: ICC = 0.66 (*F*(68,68) = 4.81, *p* < 0.001)

**DEP/ANX**: ICC = 0.61 (*F*(134,134) = 4.10, *p* < 0.001)

**SUDs**: ICC = 0.44 (*F*(82,82) = 2.62, *p* < 0.001)

ICCs for *EC* by group were as follows:

**HCs**: ICC = 0.76 (*F*(68,68) = 7.16, *p* < 0.001)

**DEP/ANX**: ICC = 0.70 (*F*(134,134) = 5.78, *p* < 0.001)

**SUDs**: ICC = 0.45 (*F*(82,82) = 2.64, *p* < 0.001)

### Intraclass correlations in descriptive task measures

For all model-free task measures, ICCs were first calculated across all participants and then by group for the five trial types.

#### Response time

ICCs for all participants were as follows:

**All trials:** ICC = 0.56 (*F*(285,285) = 3.52, *p* < 0.001)

**AV:** ICC = 0.50 (*F*(284,284) = 2.98, *p* < 0.001)

**APP:** ICC = 0.51 (*F*(285,285) = 3.05, *p* < 0.001)

**CONF2:** ICC = 0.49 (*F*(285,285) = 2.91, *p* < 0.001)

**CONF4:** ICC = 0.51 (*F*(285,285) = 3.09, *p* < 0.001)

**CONF6:** ICC = 0.43 (*F*(285,285) = 2.54, *p* < 0.001)

ICCs for HCs were as follows:

**All trials:** ICC = 0.62 (*F*(68,68) = 4.28, *p* < 0.001)

**AV:** ICC = 0.62 (*F*(68,68) = 4.25, *p* < 0.001)

**APP:** ICC = 0.54 (*F*(68,68) = 3.35, *p* < 0.001)

**CONF2:** ICC = 0.47 (*F*(68,68) = 2.77, *p* < 0.001)

**CONF4:** ICC = 0.55 (*F*(68,68) = 3.47, *p* < 0.001)

**CONF6:** ICC = 0.51 (*F*(68,68) = 2.05, *p* < 0.001)

ICCs for DEP/ANX were as follows:

**All trials:** ICC = 0.59 (*F*(133,133) = 3.88, *p* < 0.001)

**AV:** ICC = 0.47 (*F*(133,133) = 2.75, *p* < 0.001)

**APP:** ICC = 0.53 (*F*(133,133) = 3.22, *p* < 0.001)

**CONF2:** ICC = 0.54 (*F*(133,133) = 3.32, *p* < 0.001)

**CONF4:** ICC = 0.52 (*F*(133,133) = 3.20, *p* < 0.001)

**CONF6:** ICC = 0.50 (*F*(133,133) = 3.03, *p* < 0.001)

ICCs for SUDs were as follows:

**All trials:** ICC = 0.48 (*F*(82,82) = 2.81, *p* < 0.001)

**AV:** ICC = 0.36 (*F*(82,82) = 2.11, *p* < 0.001)

**APP:** ICC = 0.46 (*F*(82,82) = 2.72, *p* < 0.001)

**CONF2:** ICC = 0.44 (*F*(82,82)= 2.60, *p* < 0.001)

**CONF4:** ICC = 0.47 (*F*(82,82) = 2.78, *p* < 0.001)

**CONF6:** ICC = 0.32 (*F*(82,82) = 1.93, *p* = 0.002)

#### Average chosen runway position

ICCs for all participants were as follows:

**All trials:** ICC = 0.70 (*F*(286,286) = 5.61, *p* < 0.001)

**AV:** ICC = 0.62 (*F*(286,286) = 4.24, *p* < 0.001)

**APP:** ICC = 0.54 (*F*(286,286) = 3.36, *p* < 0.001)

**CONF2:** ICC = 0.69 (*F*(286,286) = 5.53, *p* < 0.001)

**CONF4:** ICC = 0.70 (*F*(286,286) = 5.55, *p* < 0.001)

**CONF6:** ICC = 0.71 (*F*(286,286) = 5.95, *p* < 0.001)

ICCs for HCs were as follows:

**All trials:** ICC = 0.84 (*F*(68,68) = 11.44, *p* < 0.001)

**AV:** ICC = 0.80 (*F*(68,68) = 9.17, *p* < 0.001)

**APP:** ICC = 0.50 (*F*(68,68) = 3.03, *p* < 0.001)

**CONF2:** ICC = 0.80 (*F*(68,68) = 9.01, *p* < 0.001)

**CONF4:** ICC = 0.82 (*F*(68,68) = 10.04, *p* < 0.001)

**CONF6:** ICC = 0.88 (*F*(68,68) = 15.03, *p* < 0.001)

ICCs for DEP/ANX were as follows:

**All trials:** ICC = 0.70 (*F*(134,134) = 5.69, *p* < 0.001)

**AV:** ICC = 0.56 (*F*(134,134) = 3.59, *p* < 0.001)

**APP:** ICC = 0.45 (*F*(134,134) = 2.66, *p* < 0.001)

**CONF2:** ICC = 0.71 (*F*(134,134) = 5.85, *p* < 0.001)

**CONF4:** ICC = 0.69 (*F*(134,134) = 5.39, *p* < 0.001)

**CONF6:** ICC = 0.67 (*F*(134,134) = 5.13, *p* < 0.001)

ICCs for SUDs were as follows:

**All trials:** ICC = 0.43 (*F*(82,82) = 2.50, *p* < 0.001)

**AV:** ICC = 0.53 (*F*(82,82) = 3.25, *p* < 0.001)

**APP:** ICC = 0.69 (*F*(82,82) = 5.49, *p* < 0.001)

**CONF2:** ICC = 0.33 (*F*(82,82)= 1.97, *p* = 0.001)

**CONF4:** ICC = 0.30 (*F*(82,82) = 1.85, *p* = 0.003)

**CONF6:** ICC = 0.35 (*F*(82,82) = 2.05, *p* < 0.001)

#### Choice variability

ICCs for all participants were as follows:

**All trials:** ICC = 0.62 (*F*(286,286) = 4.22, *p* < 0.001)

**AV:** ICC = 0.51 (*F*(286,286) = 3.06, *p* < 0.001)

**APP:** ICC = 0.39 (*F*(286,286) = 2.25, *p* < 0.001)

**CONF2:** ICC = 0.37 (*F*(286,286) = 2.17, *p* < 0.001)

**CONF4:** ICC = 0.38 (*F*(286,286) = 2.20, *p* < 0.001)

**CONF6:** ICC = 0.37 (*F*(286,286) = 2.20, *p* < 0.001)

ICCs for HCs were as follows:

**All trials:** ICC = 0.77 (*F*(68,68) = 7.78, *p* < 0.001)

**AV:** ICC = 0.71 (*F*(68,68) = 5.95, *p* < 0.001)

**APP:** ICC = 0.58 (*F*(68,68) = 3.80, *p* < 0.001)

**CONF2:** ICC = 0.47 (*F*(68,68) = 2.78, *p* < 0.001)

**CONF4:** ICC = 0.52 (*F*(68,68) = 3.20, *p* < 0.001)

**CONF6:** ICC = 0.34 (*F*(68,68) = 2.04, *p* = 0.002)

ICCs for DEP/ANX were as follows:

**All trials:** ICC = 0.66 (*F*(134,134) = 4.85, *p* < 0.001)

**AV:** ICC = 0.47 (*F*(134,134) = 2.74, *p* < 0.001)

**APP:** ICC = 0.29 (*F*(134,134) = 1.80, *p* < 0.001)

**CONF2:** ICC = 0.50 (*F*(134,134) = 2.96, *p* < 0.001)

**CONF4:** ICC = 0.28 (*F*(134,134) = 1.79, *p* < 0.001)

**CONF6:** ICC = 0.44 (*F*(134,134) = 2.56, *p* < 0.001)

ICCs for SUDs were as follows:

**All trials:** ICC = 0.30 (*F*(82,82) = 1.86, *p* = 0.003)

**AV:** ICC = 0.34 (*F*(82,82) = 2.03, *p* < 0.001)

**APP:** ICC = 0.37 (*F*(82,82) = 2.16, *p* < 0.001)

**CONF2:** ICC = 0.12 (*F*(82,82)= 1.27, *p* = 0.141)

**CONF4:** ICC = 0.36 (*F*(82,82) = 2.14, *p* = 0.003)

**CONF6:** ICC = 0.33 (*F*(82,82) = 1.97, *p* = 0.001)

### Intraclass correlations in descriptive task measures

ICCs for post-task survey questions are provided in **Supplementary Table 6**. Across items, the reliability was poor to good for HCs (ICCs between 0.33 and 0.68) and fair to good for DEP/ANX (ICCs between 0.45 and 0.68). In SUDs, poor to fair reliability (ICCs between 0.31 and 0.53) was found for most post-task survey questions. However, ICCs for questions about the basic approach (Q4; ICC = 0.17, *p*=0.06) and avoidance (Q5; ICC = 0.12, *p* = 0.15) motivations were statistically non-significant.

### Relationship between model parameters and demographic variables at follow-up

Similar to our findings in the *exploratory* sample and the *confirmatory* baseline sample, no sex differences were found in *EC* (*t*(131.75) = 0.86, *p* = 0.39) or *DU* (*t*(125.24) = −0.06, *p* = 0.95) at 1-year follow-up. *DU* was also positively correlated with age (*r* = 0.14, *p* =0.017). We failed to replicate the relationship between the *EC* parameter and age (*r* = 0.04, *p* = 0.51).

### Relationship between model parameters and descriptive task measures at follow-up

We replicated the relationship between model parameters and RTs and choice variability (see **Supplementary Figure 2**). In addition, as we saw in the baseline sample, there was a positive correlation between *DU* and APP RTs (*r* = 0.51, *p* < 0.001) along with *EC* and variability in runway choices in APP condition (*r* = 0.12, *p*=0.35). The relationships between model parameters and the average chosen runway position were identical to those found at baseline (see **Supplementary Figure 2** for details).

### Relationship between model parameters and post-task surveys at follow-up

Here we report group-wise differences in post-task survey responses and their correlation with model parameters. The relationship between *EC* and post-task surveys was preserved in HCs and DEP/ANX. In individuals with SUDs, the correlation between *EC* and using distraction (Q7; *r* = 0.3, *p* = 0.07) and self-regulation (Q8; *r* = 0.09, *p* = 0.39) strategies during the presentation of aversive images was not statistically significant. For the within-group correlation of *DU* and post-task surveys, the relationships observed across participants were also seen in individuals with DEP/ANX. Relationships between *DU* and avoidance of negative stimuli (Q5; HCs: *r* = 0.13, *p* = 0.31), and between *DU* and keeping eyes open during presentation of negative stimuli (Q6; HCs: *r* = −0.11 *p* = 0.39; SUDs: *r* = −0.12, *p* = 0.30) were not statistically significant. The full results of the correlational analyses between model parameters and items in the post-task survey (Q1-Q8) are shown in **Supplementary Table 7**. Additional reports of specific relationships found significant at baseline are visualized in **Supplementary Figure 3**.

### Group difference in model parameters in the sample when only including participants who returned for follow-up

We perform LMEs identical to those in the main text in a sub-sample of participants who returned for the follow-up visit. The first set of LMEs examines the main effects of the group, time, and their interaction. Additional LMEs include the effects of age, sex, and their interactions with the group, along with the effects of group and time. Lastly, LMEs accounting for WRAT scores were also performed.

Similar to the results in the main text, there was a main effect of time (*F*(1, 283) = 21.46, *p* < 0.001) on *DU*, suggesting a reduction in *DU* over time. In an LME which also includes the effects of age and sex, the main effect of time was conserved (*F*(1, 292) = 26.39, *p* < 0.001), and a main effect of age was observed (*F*(1, 279) = 7.75, *p* < 0.01), suggesting that *DU* may increase with age. These effects were conserved when accounting for WRAT scores. A main effect of WRAT scores (*F*(1, 232) = 13.69, *p*<0.001) was also found. No other statistically significant effects or interactions were found.

When predicting *EC*, a main effect of group was observed (*F*(2, 283) = 20.29, *p* < 0.001), where SUDs (EMM = 0.37) displayed lower *EC* than DEP/ANX (EMM = 0.819; *t*(279)=4.645, *p* < 0.001, *d*=1.02) and HCs (EMM = 1.065; *t*(284)=6.158, *p* < 0.001, *d*=1.58). HCs displayed greater *EC* than DEP/ANX (*t*(284)=-2.402, *p* = 0.017, *d*=-0.56). In the follow-up LME, the main effect of group (*F*(2, 278) = 13.49, *p* < 0.001) was conserved, even when controlling for WRAT scores (*F*(2, 232) = 10.83, *p* < 0.001). A *group x WRAT* interaction (*F*(2, 233) = 4.59, *p* = 0.011) was also found. No other significant effects were observed.

As in our baseline study, we repeated the LMEs in female-only (N=213) and male-only (N=74) samples. The LMEs accounted for the effect of group, age (its interaction with group), and time. As expected, there was a main effect of group on *EC* (*F*(2,207.28) = 18.46, *p* < 0.001) in females, but not in males (*F*(2,68) = 2.33, *p* = 0.105; see **Supplementary Figure 5**). Post-hoc contrasts revealed females in the DEP/ANX (EMM=0.807; *t*(207.01)=-3,125, *p <* 0.01, *d* = −0.84) and SUDs (EMM = 0.351;*t*(207.03)=6.23, *p <* 0.001, *d* = 1.86) groups had lower *EC* values than HCs (EMM = 1.184), with the lowest value for SUDs (*t*(207.04)=4.14, *p <* 0.001, *d=*1.02). No other significant effects were found.

### Standard descriptive analyses including all baseline participants

The descriptive statistics and group differences in task performance measures are provided in **Supplementary Tables 10-12**. Distributions of the average chosen runway position during the longitudinal visit are shown in **Supplementary Figure 6.** An LME was also performed to evaluate the effect of group, time, and their interaction. Follow-up LMEs were performed to account for the effects of age, sex, and their interaction with group. A summary of the replication of these results is provided in **Supplementary Figure 7**. Similar to our baseline paper, subsequent LME also accounted for the effect of the WRAT score and its interaction with group. Analogous analyses were also performed in a sub-sample of participants, where the baseline data was limited to participants who returned for their follow-up visit. These results are described below. The LMEs were also performed in a sub-sample of participants who returned for the follow-up visit (see **Supplementary Tables 13-21)**.

#### Response time

When considering all trials, there was a main effect of time (*F*(1, 340) = 11.33, *p* < 0.001), whereby participants displayed faster RTs at 1-year follow-up. This was also seen in APP, CONF2, CONF4, and CONF6 trials (*ts ≥* 2.55, *ps ≤* 0.014). A main effect of group was observed in AV trails (*F*(2, 487) = 14.13, *p* < 0.001), where SUDs (EMM = 1.489) showed slower RTs than both HCs (EMM = 1.303; *t*(462.37)=-4.751, *p* < 0.001, *d* = −0.75) and DEP/ANX (EMM = 1.332; *t*(479.75)=-4.902, *p* < 0.001, *d* = −0.63). No other effects of group, time, or their interaction were found.

Next, we accounted for possible effects of age and sex. Across all trials, the main effect of time was conserved (*F*(1, 356.08) = 20.40, *p* < 0.001); a positive association with age was also present (*F*(1, 459.06) = 97.16, *p* < 0.001). When restricting to specific trial types, there was a main effect of the group in AV (*F*(2, 465) = 14.29, *p* < 0.001) and APP (*F*(2, 466) = 3.22, *p* = 0.041) trials. During AV trials, SUDs (EMM = 1.491) had slower response times than HCs (EMM = 1.326, *t*(442.01)=-5.389, *p* < 0.001, *d*=-0.78) and DEP/ANX (EMM = 1.326, *t*(468.92)=-5.671, *p* < 0.001, *d*=-0.67). In the APP condition, DEP/ANX (EMM = 1.187) had faster response times than SUDs (EMM = 1.255; *t*(470.06)=-2.216, *p* = 0.027, *d*=-0.28). A *group x sex* interaction (*F*(2, 467) = 7.64, *p* < 0.001) was present in the AV trials. In females, SUDs (EMM = 1.51) displayed the highest RTs, followed by DEP/ANX (EMM = 1.338) and HCs (EMM = 1.237; *ts≥*2.43, *ps ≤* 0.016). In males, DEP/ANX (EMM = 1.297) were significantly faster than HCs (EMM = 1.442; *t*(457.87)=-2.415, *p* = 0.016, *d*=-0.59) and SUDs (EMM = 1.448; *t*(506.08)=-2.792, *p* = 0.005, *d*=-0.61). Significant effects of age (*F*s ≥ 55.94, *p*s < 0.001) and time (*Fs* ≥ 9.18, *ps* ≤ 0.003) were observed within all trial types. No other significant main effects or interactions were observed. When accounting for the effect of WRAT scores, a significant *group x WRAT* interaction was found in all conflict trials and when all trials were considered together (*Fs* > 3.41, *ps ≤* 0.034). There was a main effect of WRAT (*F*s *>* 5.90, *ps* < 0.016) in all comparisons except for the CONF2 and CONF4 trials. In addition, a main effect of sex (*F*(1, 381) = 4.64, *p* = 0.032) was also present in AV trials, such that males had slower response times than females.

#### Chosen Runway Position

There was a main effect of group when all trials were considered and for each trial type (*Fs* ≥ 4.73, *ps* ≤ 0.009), except for APP trials (*F*(2, 494) = 1.22, *p* = 0.295). Post-hoc contrast revealed that SUDs consistently displayed lower avoidance than HCs (|*ts*| ≥ 4.34, *ps* < 0.001) and DEP/ANX (|*ts*| ≥ 3.19, *ps* ≤ 0.002) in the AV and CONF conditions.

Notably, there was also a main effect of time in APP (*F*(1, 334) = 5.91, *p* = 0.016) and AV trials (*F*(1, 336) = 11.16, *p* < 0.001), indicating a greater drive to approach the points (EMM_Baseline_ = 8.183; EMM_Follow-up_ = 8.396) and avoid the aversive image (EMM_Baseline_ = 7.648; EMM_Follow-up_ = 7.998) in the absence of conflict at 1-year follow-up. A *group x time* interaction was found in AV trials (*F*(2, 337) = 3.60, *p* = 0.028), whereby SUDs (EMM_Baseline_ = 6.782; EMM_Follow-up_ = 7.453) at both time points displayed lower avoidance than HCs (Baseline: EMM = 8.249, *t*(615.99)=6.568, *p* < 0.001, *d*=1.29; Follow-up: EMM = 8.27; *t*(754.42)=3.055, *p* = 0.002, *d* = 0.72) and DEP/ANX (Baseline: EMM = 8.009, *t*(615.99)=6.78, *p* < 0.001, *d* = 1.08; Follow-up: EMM = 8.267; *t*(760.71)=3.601, *p* < 0.001, d=0.72). No other significant main effects or interactions were observed.

The majority of these results were conserved when effects of age, sex, and their interactions with the group were included (*Fs* ≥ 7.59, *ps* < 0.001). There was a main effect of sex when all trials were considered (*F*(1, 472) = 4.66, *p* = 0.031) and in all CONF conditions (*Fs* ≥ 4.10, *ps* ≤ 0.043), whereby males displayed lower avoidance than females. *Group x sex* interaction was seen in CONF2 and CONF4 conditions, where we see the conservation of group effects in females (|*ts|* ≥ 2.942, *ps* ≤ 0.003) but not in males. The main effect of age was also present in all conditions (*Fs* ≥ 6.59, *ps* ≤ 0.008). Lastly, when controlling for the effects of the WRAT score and its interaction with group, *group x sex* interaction was found in CONF6 trials (*F*(2, 382) = 3.11, *p*<0.05). There was main effect of WRAT in APP (*F*(1, 364) = 15.78, *p*<0.001) and AV (*F*(1, 360) = 26.05, *p*<0.001) trials and *group x WRAT* interaction in CONF2 trials (*F*(2, 371) = 3.82, *p* = 0.023).

#### Choice Variance

As expected, the choice variance was lower at follow-up compared to baseline when all trials were considered and within each trial type (*Fs* < 5.90, *ps* ≤ 0.016). SUDs (EMM_APP_ = 1.006; EMM_AV_ = 1.543) also displayed higher choice variance than HCs (EMM_APP_ = 0.605; EMM_AV_ = 0.799) and DEP/ANX (EMM_APP_ = 0.708; EMM_AV_ = 0.844) group for the AV and APP trials (|*ts*| > 2.81, *ps* ≤ 0.005). We failed to replicate the group differences (*F*(2, 481) = 0.66, *p* = 0.518) in the CONF6 trial, which were also absent at baseline. There was also a *group x time* interaction in AV trials (*F*(2, 352) = 3.48, *p* = 0.032), indicating that SUDs (EMM_Baseline_ = 1.755; EMM_Follow-up_ = 1.189) also had higher choice inconsistencies than HCs (EMM_Baseline_ = 0.831; EMM_Follow-up_ = 0.746) and DEP/ANX (EMM_Baseline_ = 0.949; EMM_Follow-up_ = 0.668) at baseline and follow-up (|*ts*| < 2.40, *ps ≤* 0.017). These results were conserved when effects of age, sex, and their interactions with group were included (*Fs* < 6.62, *ps ≤* 0.001). When all trials were considered, there was a main effect of sex (*F*(1, 471) = 4.33, *p* = 0.038), whereby females (EMM = 2.01) displayed greater variability in their choices than males (EMM = 1.79). There was also a main effect of age (*Fs ≥* 13.52, *ps <* 0.001) for each trial type and when all trials were considered (*F*(1, 463) = 13.63, *p* < 0.001). A *group x* age interaction (*Fs ≥* 3.96, *ps* ≤ 0.02) was found for all CONF trials, and a main effect of group was also found for CONF4 trials (*F*(2, 464) = 3.91, *p* = 0.021). Lastly, additional LMEs were performed to account for the effects of WRAT score and its interaction with group. A main effect of WRAT scores was found in each individual trial type (*Fs ≥* 12.57, *ps <* 0.001). Once included, the *group x age* interaction in CONF trials (*Fs* ≤ 2.64, *ps*≥0.073) and group effect in APP trials (*F*(2, 372) = 0.45, *p* = 0.641) were no longer significant.

### Group differences in post-task self-report questionnaire

Additional LMEs were performed to assess differences in select self-report questionnaire items (**Table 5**, main text) measuring anxiety (Q2), difficulty in decision-making during the task (Q3), approach motivation (Q4) and avoid motivation (Q5). A summary of the replication of these results is provided in **Supplementary Figure 8**. Analogous analyses were also performed in a sub-sample of participants, where the baseline data was limited to participants who returned for their follow-up visit. These results are provided in **Supplementary Tables 22-24**.

For self-reported anxiety (Q2), a main effect of group (*F*(2, 489) = 3.99, *p* = 0.019) was observed. SUDs (EMM = 3.72) displayed lower self-reported anxiety than DEP/ANX (EMM = 4.22; *t*(480.34) = 2.685, *p* = 0.007, *d* = 0.37). When effects of age and sex were considered, sex effect (F(1, 468) = 11.57, *p* < 0.001) and *group x sex* interaction (*F*(2, 469) = 5.75, *p* < 0.001) better explained the group effects (*F*(2, 273) = 1.06, *p* = 0.349). In female participants SUDs (EMM = 3.68) reported lower anxiety than DEP/ANX (EMM = 4.36; *t*(457.41)=3.103, *p* = 0.002, *d* = 0.51) and HCs (EMM = 4.52, *t*(438.16)=2.988, *p* = 0.003, *d* = 0.62). No significant group effects were found in males. No other main effects or interactions were found. However, when accounting for WRAT scores, only a main effect of sex was found (*F*(1, 382) = 11.08, *p* < 0.001). Females (EMM =4.15) reported greater anxiety (*t*(387.68)=3.115, *p* = 0.002, *d* = 0.51) than males (EMM = 3.46).

For self-reported difficulty in decision-making during the task (Q3), there was a main effect of time (*F*(1, 363) = 4.72, *p* = 0.03), suggesting participants had less difficulty making decisions during the task at follow-up (EMM_Baseline_ = 2.31; EMM_Follow-up_ = 2.10; *t*(367.49) = 2.14, *p* < 0.05, *d* = 0.17). When the effects of age and sex were considered, the main effect of time (*F*(1, 368) = 5.51, *p* = 0.019) persisted. In addition, a main effect of age (*F*(1, 455) = 6.63, *p* = 0.010) and *group x age* interaction was found (*F*(2, 457) = 4.55, *p* = 0.011). No additional main effects or interactions were found. However, when accounting for WRAT scores, no significant main effects or interactions were found (*Fs* ≤ 3.38, *ps* ≥ 0.067).

For self-reported approach motivation (Q4), main effect of group (*F*(2, 492) = 6.84, *p* < 0.001) was observed. SUDs (EMM = 5.40) displayed lower self-reported approach motivation than DEP/ANX (EMM = 4.76; *t*(482.32)=-2.807, *p* = 0.005, *d* = −0.41) and HCs (EMM = 4.41; *t*(467.38)=-3.598, *p* < 0.001, *d* = −0.65). These effects (*F*(2, 468) = 4.87, *p* = 0.008) were conserved after accounting for the effects of age and sex. A main effect of age was also found (*F*(1, 461) = 5.67, *p* = 0.018). No other main effects or interactions were found. Identical results were found when accounting for the effects of WRAT scores.

For self-reported avoid motivation (Q5), main effect of group (*F*(2, 488) = 6.27, *p* = 0.002) was observed. SUDs (EMM = 2.41) displayed lower self-reported avoidance motivation than DEP/ANX (EMM = 3.08; *t*(479.91)=3.248, *p* = 0.001, *d* = 0.44) and HCs (EMM = 3.24; *t*(463.81)=3.29, *p* = 0.001, *d* = −0.54). These effects (*F*(2, 467) = 4.18, *p* = 0.016) were conserved after accounting for the effects of age and sex. A *group x sex* interaction (*F*(2, 469) = 4.38, *p* = 0.014) was found, where these group differences (Females: EMM_HCs_ = 3.64; EMM_DEP/ANX_ = 3.14, EMM_SUDs_ = 2.204; Male: EMM_HCs_ = 2.56; EMM_DEP/ANX_ = 2.90; EMM_SUDs_ = 2.67) were driven by female participants (DEP/ANX - SUDs: *t*(457.54)=3.717, *p* < 0.001, *d* = 0.61; HCs - SUDs: *t*(438.38)=4.489, *p* < 0.001, *d* = 0.93). However, when we controlled for WRAT scores, the group effect (*F*(2, 375) = 4.41, *p* = 0.013) was conserved, and the *group x sex* interaction (*F*(2, 381) = 2.99, *p*=0.052) was marginal. In addition, a main effect of age (*F*(1, 365) = 4.69, *p* = 0.031) was found.

### Group differences in the relationship between response times and chosen runway position

An LME was performed to examine potential group differences in the relationship between RTs and chosen runway position in the combined exploratory and confirmatory sample: *chosen runway position ∼ group + time + RTs + group*time + group*RTs + group*time*RTs*. There was a main effect of group (*F*(2, 936.06) = 10.93, *p* < 0.001), reflecting greater approach behavior in DEP/ANX (EMM = 7.23; *t*(915) = −2.52, *p* = 0.025) and SUDs (EMM = 7.58; *t*(946) = −4.46, *p* < 0.001) than in HCs (EMM = 6.91) overall. SUDs also had greater approach behavior than DEP/ANX (*t*(977) = −3.14, *p* = 0.002). An overall negative relationship was found between RTs and chosen runway position (*F*(1, 1553.42) = 141.32, *p* < 0.001). This LME also revealed a *group x RT* interaction (*F*(2, 1547.32) = 4.78, *p* = 0.009), suggesting a stronger negative association between RTs and approach behavior in SUDs (ET = −1.94) than in HCs (ET = −0.59; *t*(1533) = 3.13, *p* = 0.005). Lastly, there was a *group x time* x *RT* interaction (*F*(3, 831.67) = 3.43, *p* = 0.017). This interaction was due to a decrease in the magnitude of the inverse relationship between RTs and runway position over time in the DEP/ANX group (ET_Baseline_ = −1.75; ET_Follow-up_ = −0.93; *t*(812) = −2.95, *p* = 0.003), while the relationship between RTs and runway position remained relatively stable over time in the other two groups (HC: ET_Baseline_ = −0.91; ET_Follow-up_ = −0.28; *t*(779) = −1.20, *p* = 0.230; SUDs: ET_Baseline_ = −1.87; ET_Follow-up_ = −2.00; *t*(911) = 0.37, *p*=0.713). This led to a stronger negative relationship between RTs and runway position at follow-up in SUDs compared to DEP/ANX at follow-up (*t*(1200) = 2.59, *p*=0.010), which was not present at baseline (*t*(1539) = 0.37, *p*=0.715).

### Group differences in the relationship between response times and model parameters

LMEs were performed to examine potential group differences in the relationship between RTs and model parameters in the combined exploratory and confirmatory samples: *model parameter ∼ group + time + RTs + group*time + group*RTs + group*time*RTs*. There was a reduction in *DU* over time (*F*(1, 768.8) = 79.25, *p* < 0.001; EMM_Baseline_=1.08; EMM_Follow-up_=0.78; *t*(767) = 7.75, *p* < 0.001), and a positive relationship between RTs and *DU* (*F*(1, 1487.59) = 643.36, *p* < 0.001) was found. As expected, a main effect of group (*F*(2, 920.80)=5.38, *p*=0.005) was found, whereby SUDs (EMM = 1.08) had greater *DU* than DEP/ANX (EMM = 0.94; *t*(963) = −2.51, *p* = 0.012) and HCs (EMM = 0.87; *t*(927) = −2.93, *p* = 0.003.

The LME was repeated for *EC*. There was also a reduction in *EC* over time (*F*(1, 712.07) = 8.14, *p* = 0.005; EMM_Baseline_ = 0.74; EMM_Folllow-up_ = 0.81; *t*(709) = −2.49, *p* = 0.013), and a positive relationship between RTs and *EC* (*F*(1, 1555.99) = 83.77, *p* < 0.001) was also present. There was a main effect of group (*F*(2, 938.56) = 41.92, *p* < 0.001), reflecting lower *EC* in the clinical sample (DEP/ANX: EMM = 0.82; *t*(919) = 3.11, *p* = 0.002; SUDs: EMM = 0.45; *t*(949) = 8.16, *p* < 0.001) when compared to HCs (EMM = 1.03). Within the clinical sample, SUDs had lower *EC* than DEP/ANX (*t*(979) = 7.01, *p* < 0.001). This LME also revealed a *group x RT* interaction (*F*(2, 1554.12) = 3.48, *p* = 0.031), suggesting a stronger positive association between RTs and *EC* in SUDs (ET = 0.69) than in HCs (ET = 0.17; *t*(1556) = −2.64, *p* = 0.023). Lastly, there was a *group x time* x *RT* interaction (*F*(3, 812.34) = 3.41, *p* = 0.017). This interaction was due to a decrease in the magnitude of the relationship between RTs and runway position over time in the DEP/ANX group (ET_Baseline_ = 0.64; ET_Follow-up_ = 0.28; *t*(793) = −2.89, *p* = 0.004), while the relationship between RTs and runway position remained relatively stable over time in the other two groups (HC: ET_Baseline_ = 0.33; ET_Follow-up_ = 0.01; *t*(764) = 1.38, *p* = 0.17; SUDs: ET_Baseline_ = 0.69; ET_Follow-up_ = 0.70; *t*(887) = −0.04, *p*=0.97). This led to a stronger positive relationship between RTs and *EC* at follow-up in SUDs compared to DEP/ANX at follow-up (*t*(1167) = −2.20, *p*=0.028), which was not present at baseline (*t*(1531) = −0.31, *p*=0.75).

## Figures

**Supplementary Figure 1.**
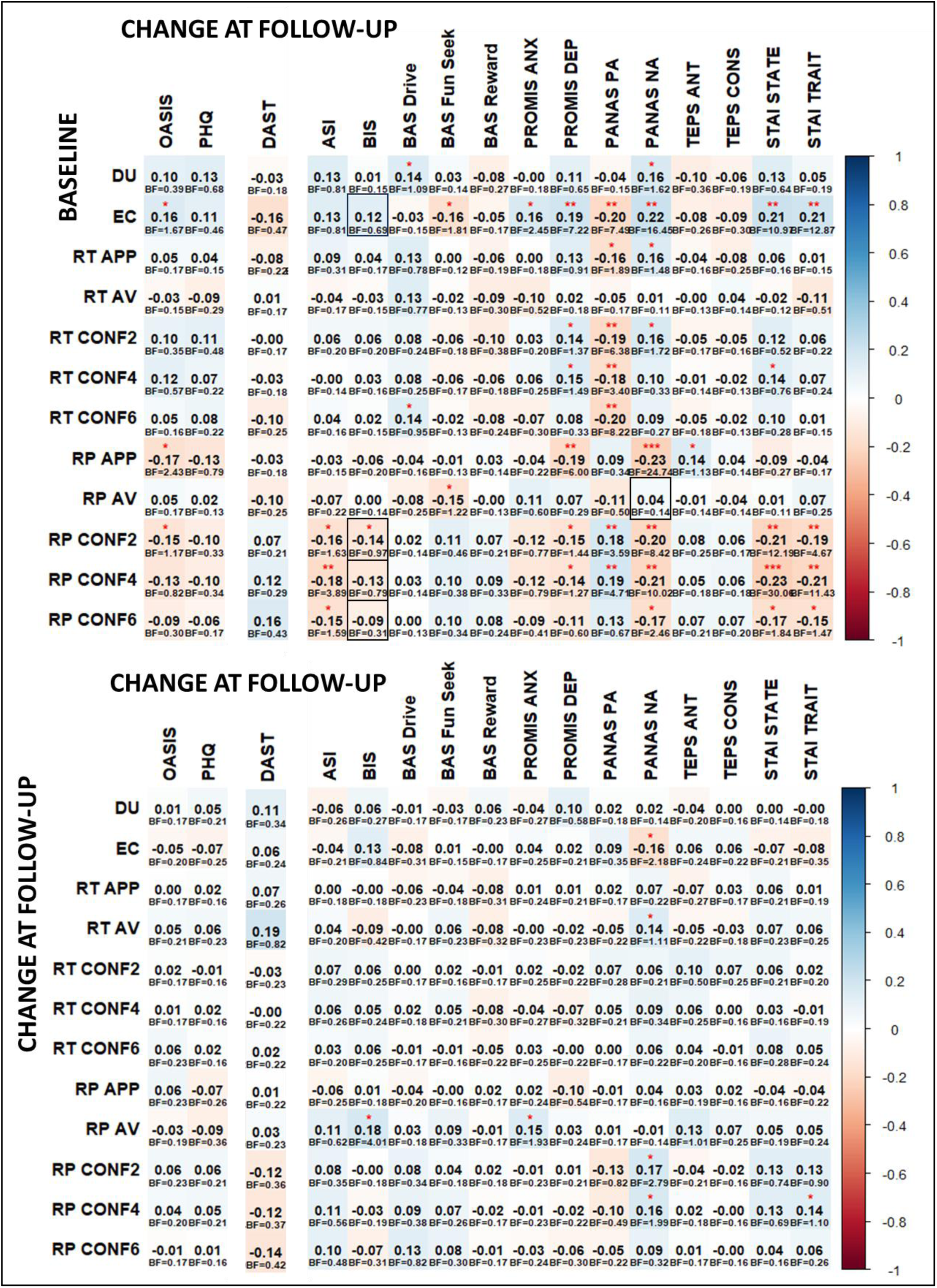
(*Top*) Partial correlations between task measures at baseline and symptom change at follow-up, accounting for baseline symptom severity. (*Bottom*) Partial correlations between changes over time in task measures and symptom severity, accounting for baseline symptom severity and baseline task performance *Left*: relationship with clinical symptoms. *Right*: relationship with dimensional measures as visualized in the *exploratory* paper. RT=response time; RP=average chosen runway position; Boxes represent significant partial correlations in the exploratory dataset. Uncorrected p-values: \**p* < 0.001, \*\**p* < 0.01, \**p* < 0.05 **Note:** For analyses involving the DAST scores, data was included only for individuals who met the criteria for substance use.

**Supplementary Figure 2.**
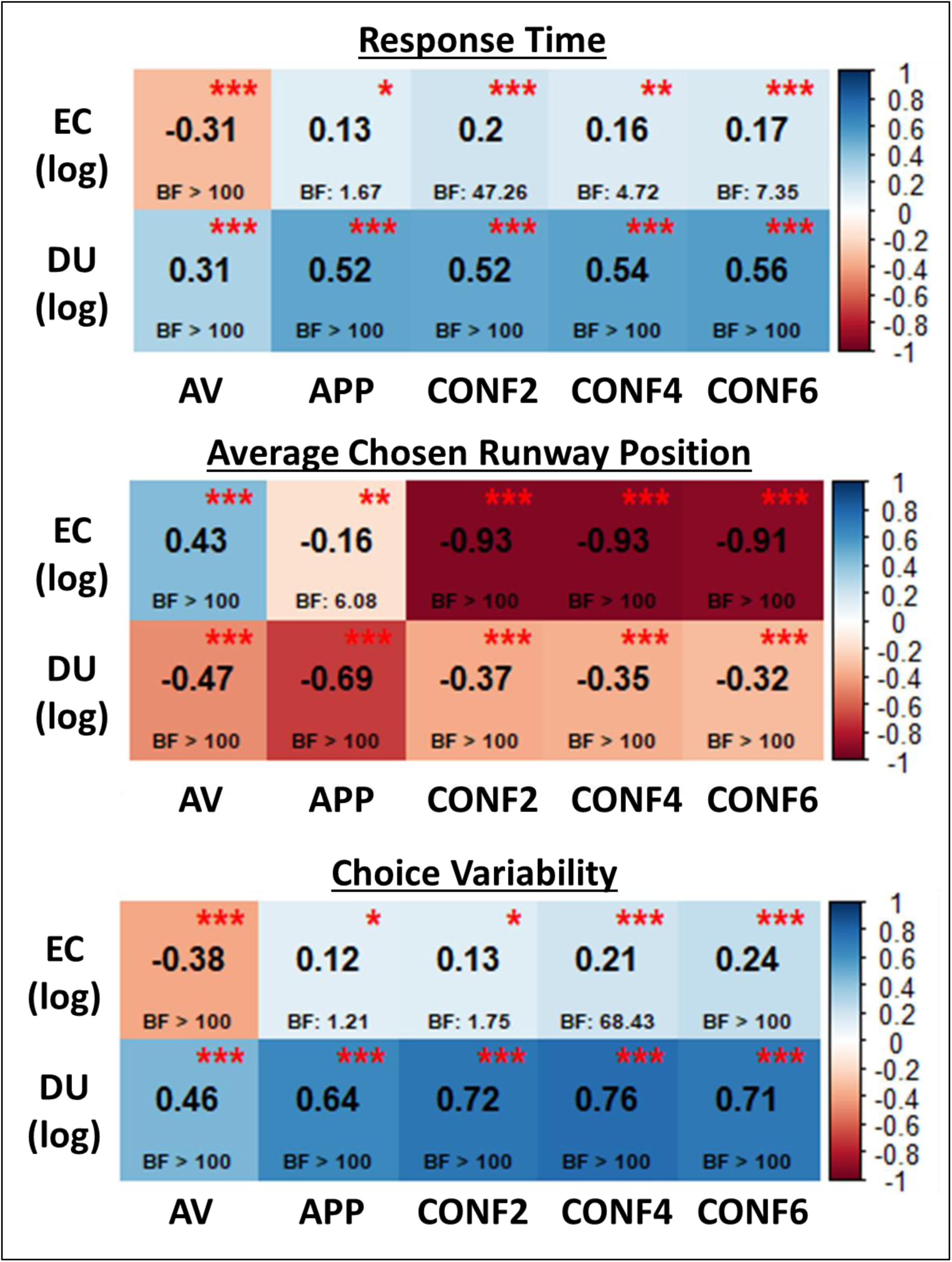
Pearson correlations and Bayes factor values between model parameters and descriptive task measures (top: reaction time, middle: average chosen runway position, and bottom: choice variability). Uncorrected *p*-values: \*\*\**p* < 0.001, \*\**p* < 0.01, \**p* < 0.05.

**Supplementary Figure 3.**
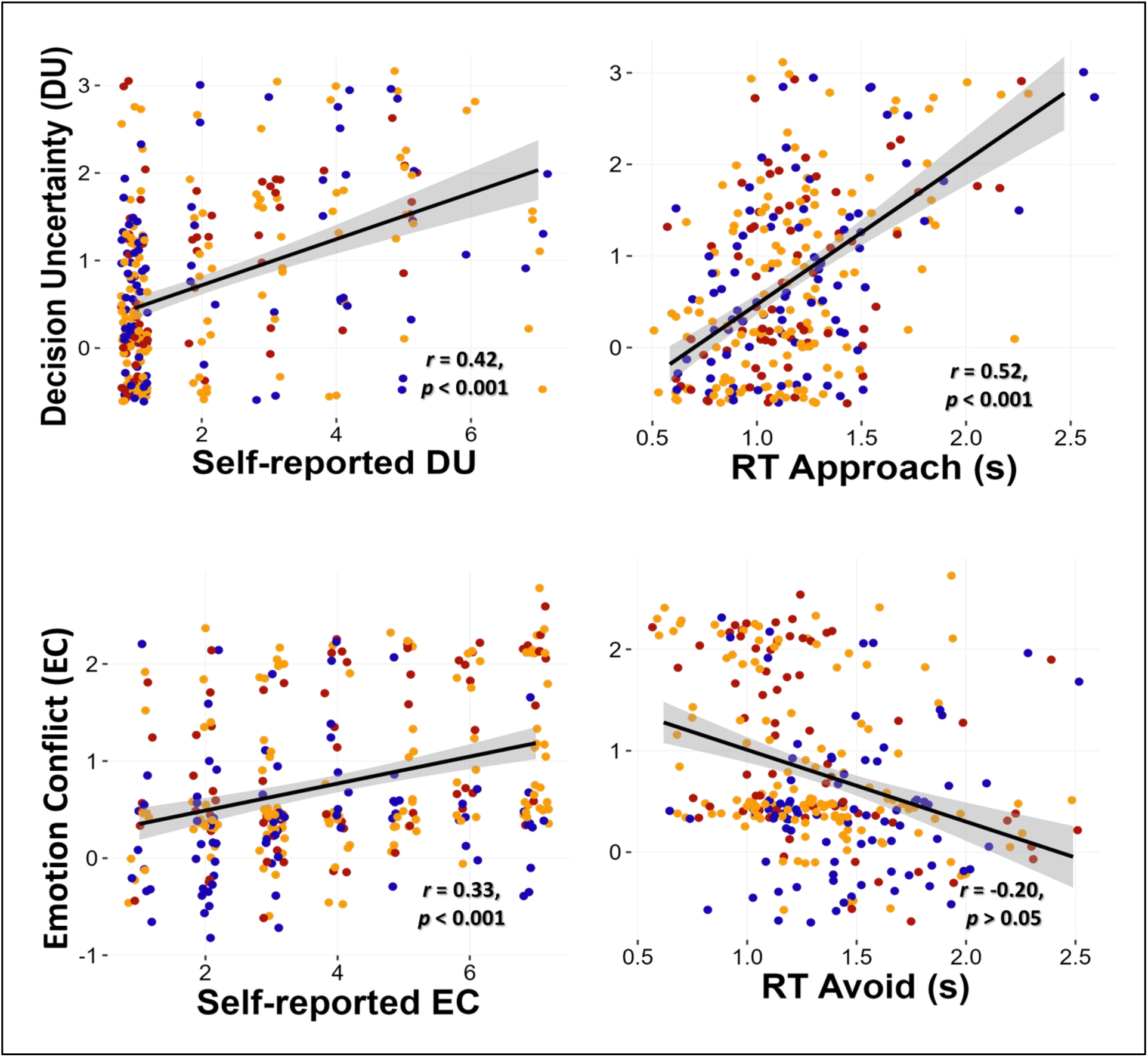
Association between model parameters (*top*: *DU* and *bottom*: *EC*) and (*left*) select self-report questionnaire items (Q2 and Q3 in **Table 4**, main text) and (*right*) response time (RTs) in approach (*top*) and avoid (*bottom*) conditions.

**Supplementary Figure 4.**
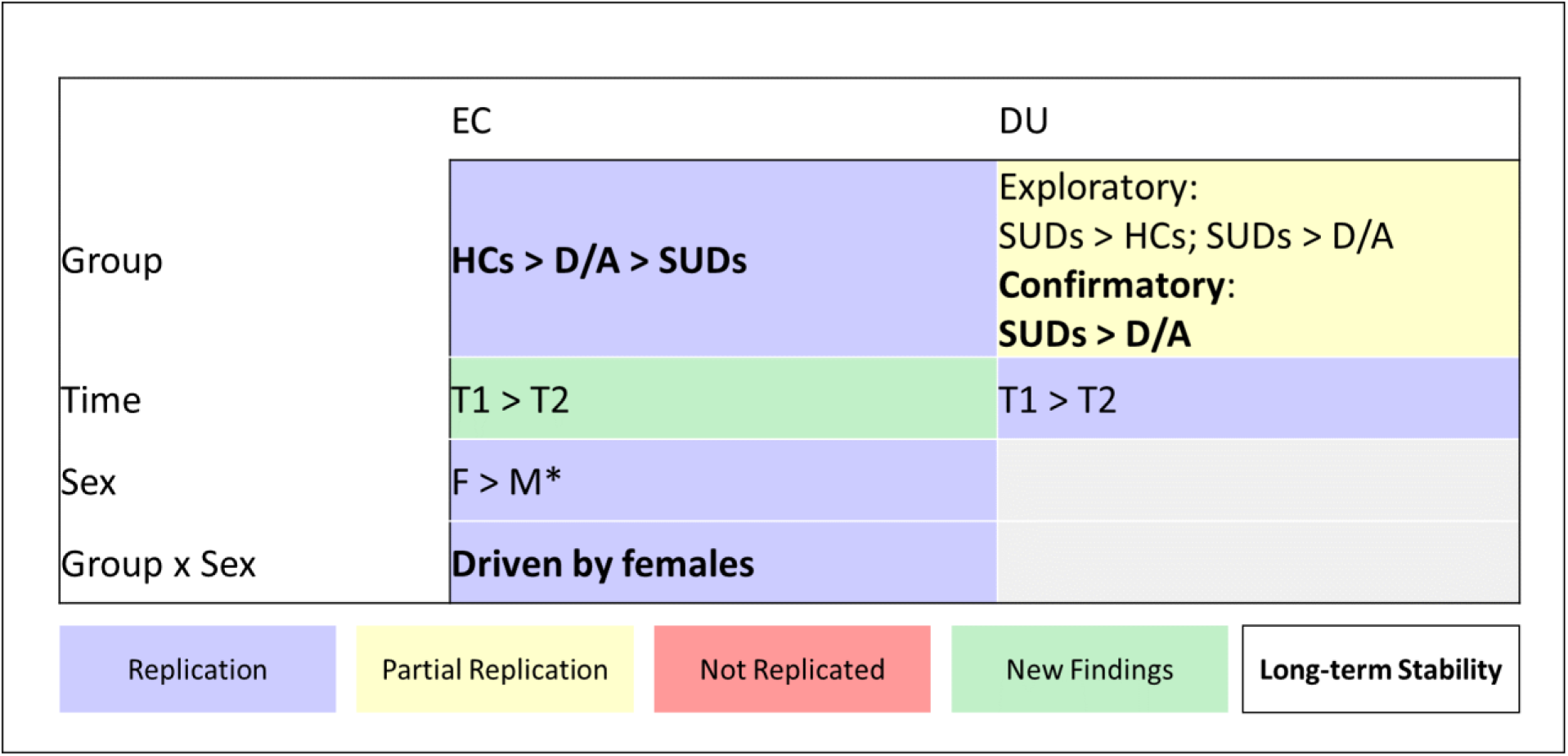
Summary of replication and long-term stability of the model parameters. HCs = Healthy Comparisons; D/A = Depression and Anxiety; SUDs = Substance use disorder; T1 = Baseline (including all participants); T2 = Follow-up; F = Females; M = Males <u>Replication</u> = congruent results in the exploratory and confirmatory samples <u>Partial Replication</u> = replication of main effect or interactions, not all post-hoc contrasts were replicated <u>Not Replicated</u> = statistically significant results in the exploratory sample, which were not significant in the confirmatory sample <u>New Findings</u> = statistically significant results in the confirmatory sample, which were not significant in the exploratory sample <u>Long-term Stability</u> = statistically significant results at baseline (including all participants) which were also significant at follow-up in the exploratory sample **Note**: The figure is an illustrative representation of comparable results and not a reflection of a complete model. *Replication of main effect only. Specific post-hoc contrast is from the present study.

**Supplementary Figure 5.**
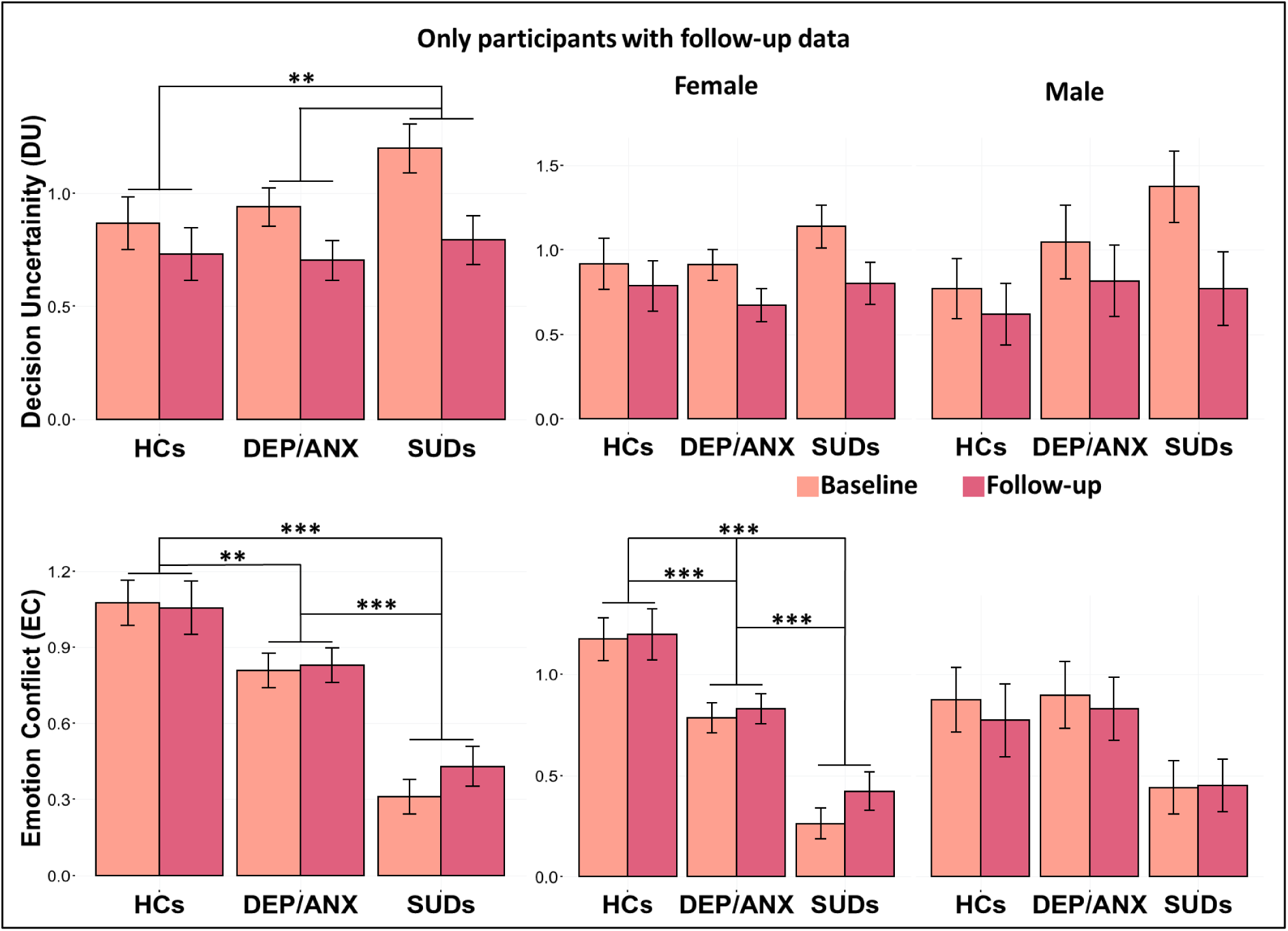
Group differences *are here* visualized as bar plots for only those participants who returned for the*ir* follow-up. Top: All participants display lower decision uncertainty at their follow-up visit. Bottom: SUDs display significantly lower emotion conflict and the observed group differences appear*ed* to be driven by females. \*\*\**p* < 0.001, \*\**p* < 0.01, \**p* < 0.05.

**Supplementary Figure 6.**
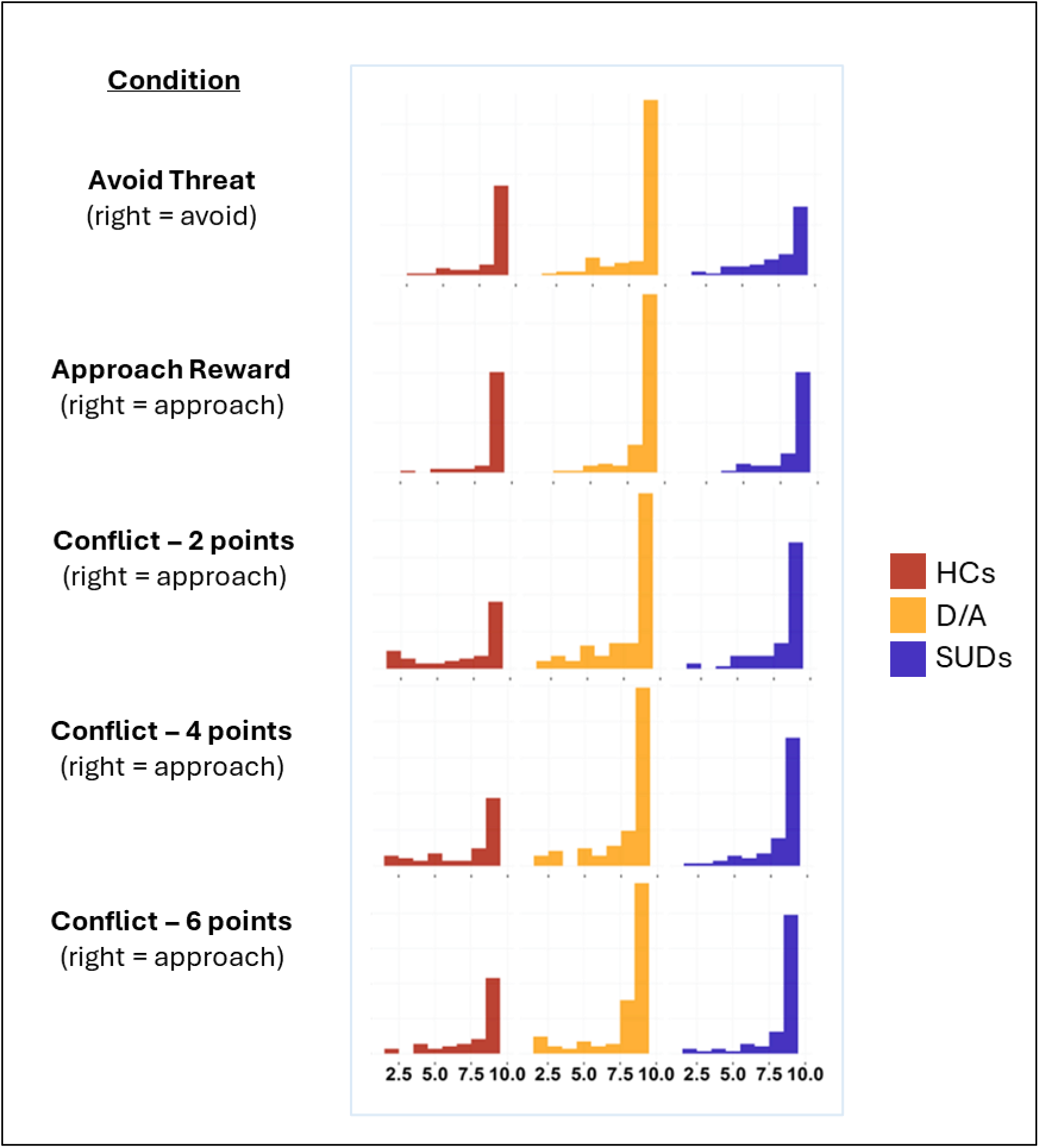
Histograms depicting average chosen runway position separated by group and condition. The x-axis represents the frequency of chosen runway positions. In all but *avoid* trials, the right most position (position 10) indicates maximum approach behavior, while the left-most position (position 2) indicates maximum avoidance (*note*: position 1 was coded as the “undecided” position and therefore not included here). HCs appeared to prefer either maximum approach or maximum avoidance depending on trial type. In conflict trials, there appeared to be some preference for avoidance, which decreased with increasing point values. For D/As, as the conflict gradient increased, participants appeared to be more confident in their approach/avoidance preferences. Lower avoidance was observed for SUDs across all conditions relative to the other two groups.

**Supplementary Figure 7.**
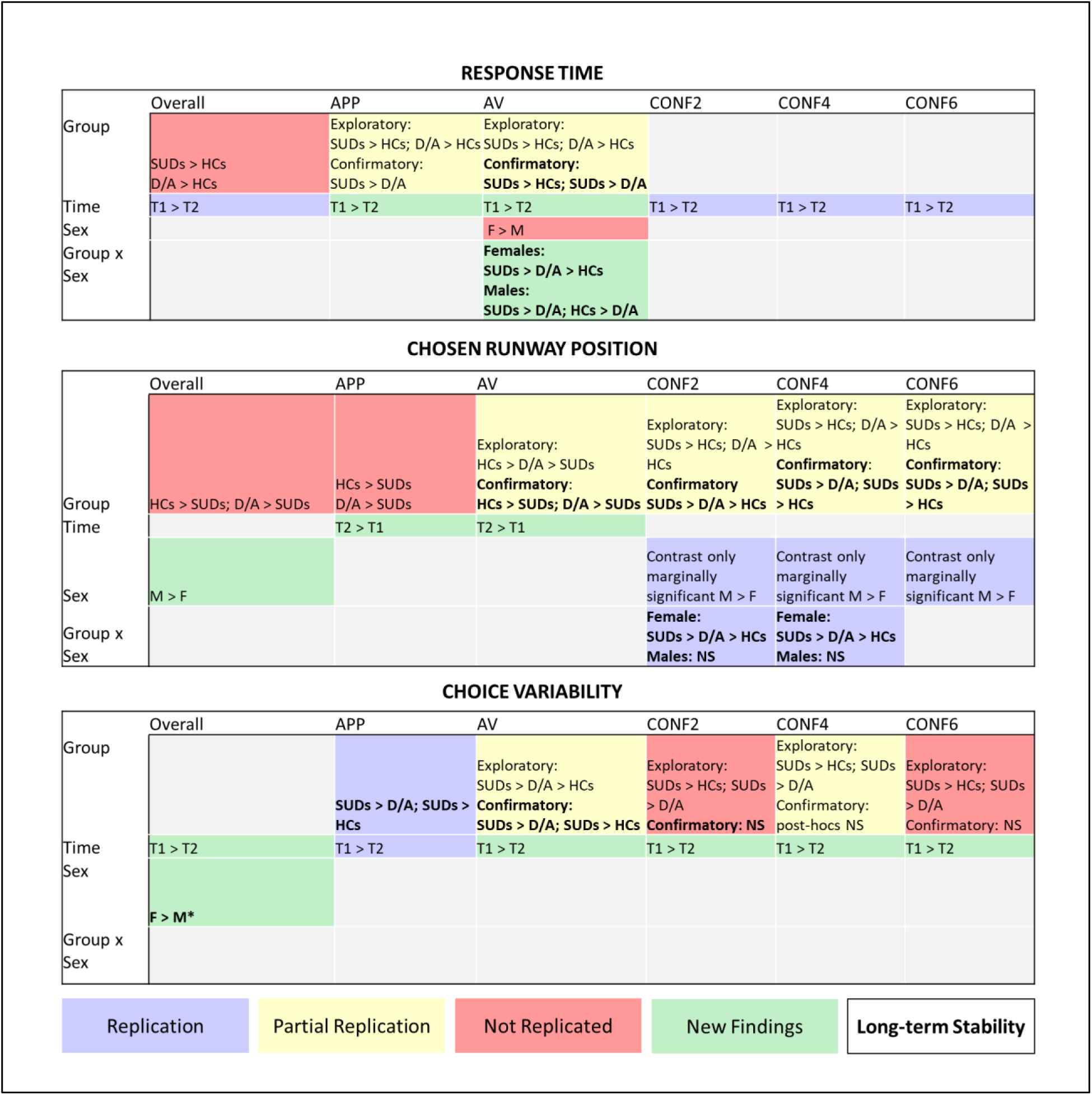
Summary of replication and long-term stability of descriptive task measures. HCs = Healthy Comparisons; D/A = Depression and Anxiety; SUDs = Substance Use Disorder; T1 = Baseline (including all participants); T2 = Follow-up; M = Males; F = Females <u>Replication</u> = congruent results in the exploratory and confirmatory samples <u>Partial Replication</u> = replication of main effect or interactions, not all post-hoc contrasts were replicated <u>Not Replicated</u> = statistically significant results in the exploratory sample, which were not significant in the confirmatory sample <u>New Findings</u> = statistically significant results in the confirmatory sample, which were not significant in the exploratory sample <u>Long-term Stability</u> = statistically significant results at baseline (including all participants) which were also significant at follow-up in the exploratory sample **Note**: The figure is an illustrative representation of comparable results and not a reflection of a complete model. *Significance found only at the main effects level.

**Supplementary Figure 8.**
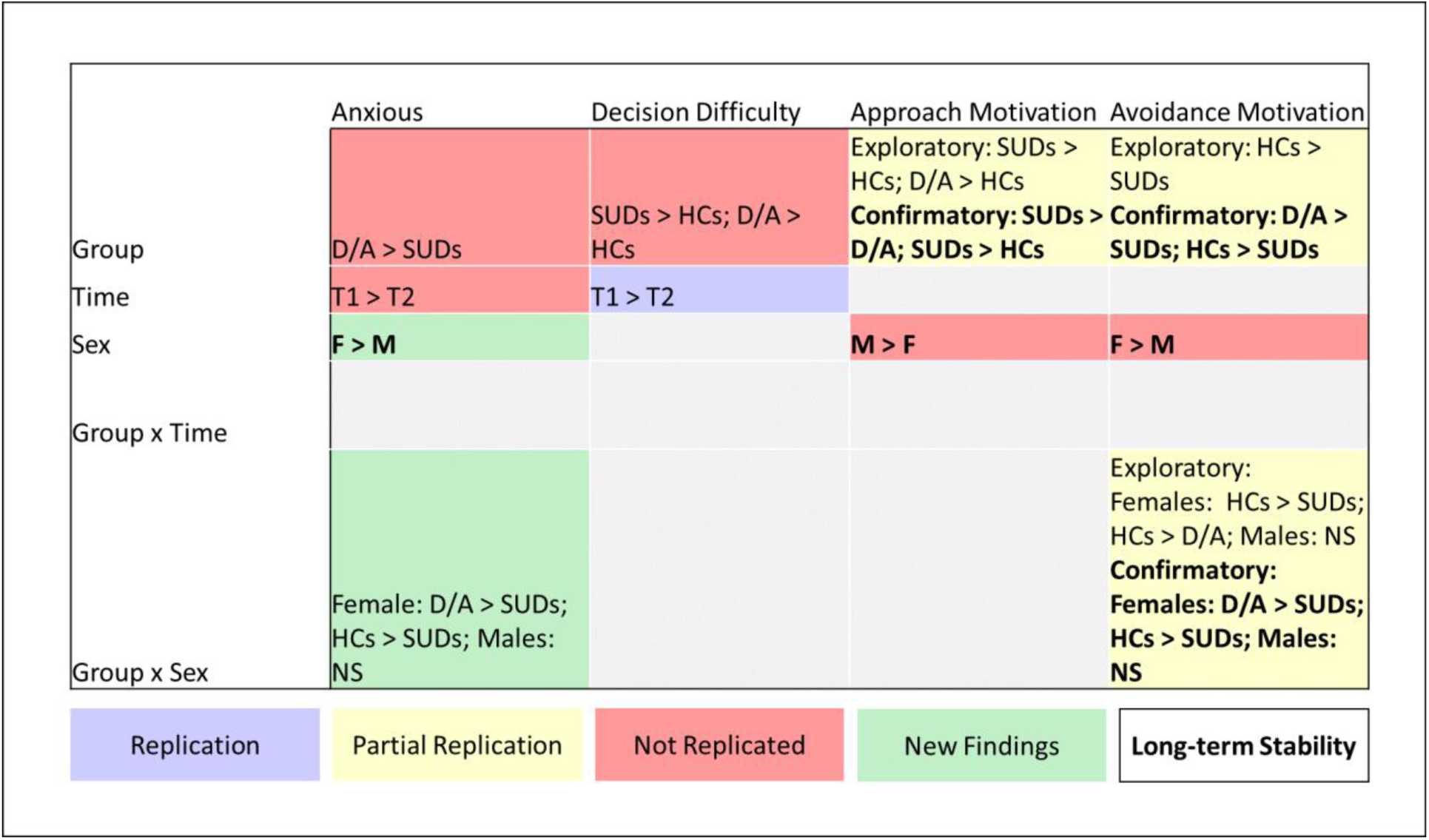
Summary of replication and long-term stability of self-reported post-task questionnaire items. HCs = Healthy Comparisons; D/A = Depression and Anxiety; SUDs = Substance Use Disorder; T1 = Baseline (including all participants); T2 = Follow-up; M = Males; F = Females; NS = Non-significant <u>Replication</u> = congruent results in the exploratory and confirmatory samples <u>Partial Replication</u> = replication of main effect or interactions, not all posthoc contrasts were replicated <u>Not Replicated</u> = statistically significant results in the exploratory sample, which were not significant in the confirmatory sample <u>New Findings</u> = statistically significant results in the confirmatory sample, which were not significant in the exploratory sample <u>Long-term Stability</u> = statistically significant results at baseline (including all participants) which were also significant at follow-up in the exploratory sample Note: The figure is an illustrative representation of comparable results and not a reflection of a complete model.

**Supplementary Figure 9.**
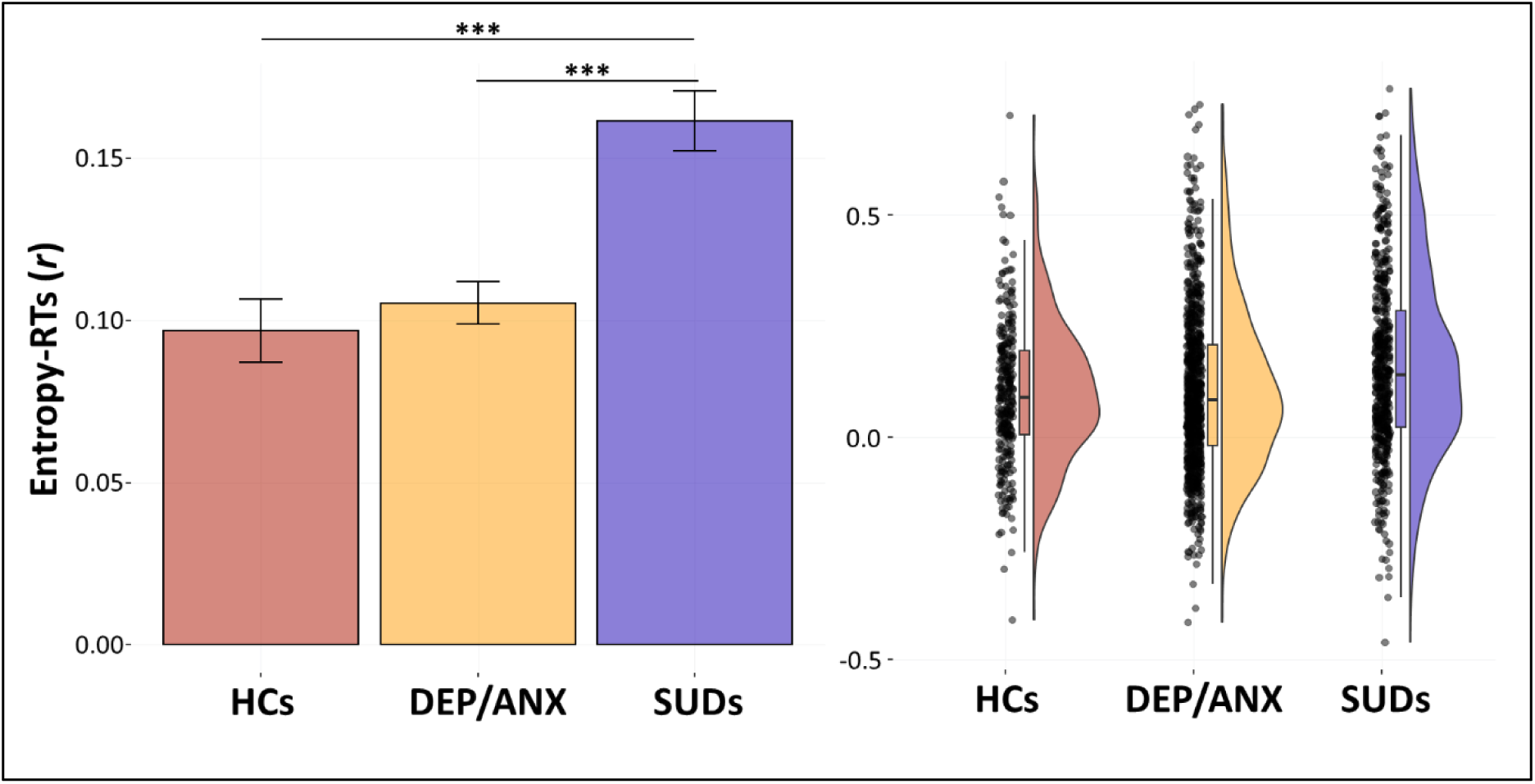
Trial-by-trial measure of choice uncertainty (entropy) predicts response time. HCs = Healthy Comparisons; DEP/ANX = Depression and Anxiety; SUDs = Substance Use Disorder; \*\*\**p* < 0.001, \*\**p* < 0.01, \**p* < 0.05.

**Supplementary Figure 10.**
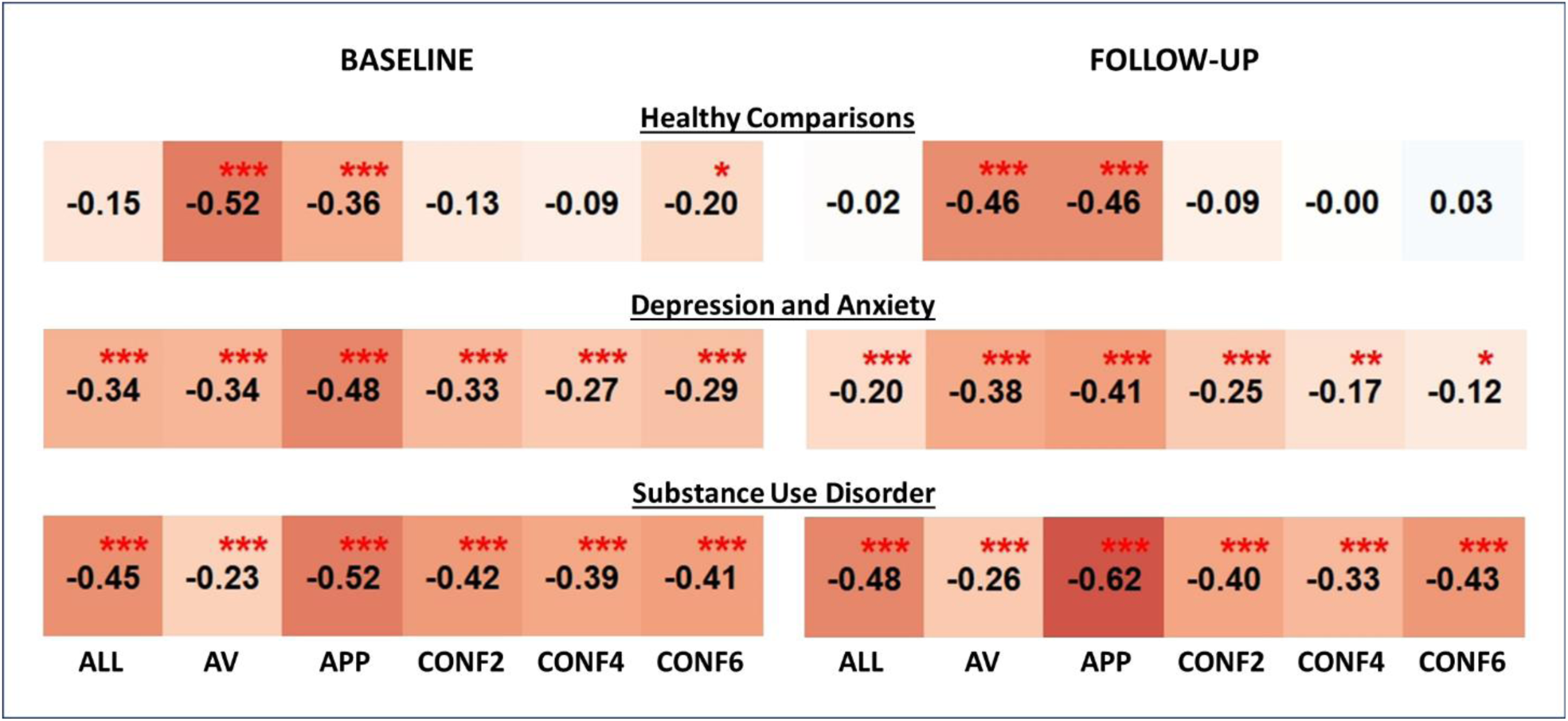
Correlations between response time and chosen runway position were performed at baseline (*left*) and follow-up (*right*) for Healthy Comparisons (*top*), Depression and Anxiety (*middle*) and Substance Use Disorder (*bottom*) group. Each cell represents a correlation between response time and chosen runway position either across all trials (ALL) or for a specific trial type (AV: avoid trials; APP: approach trials; CONF2: Conflict 2 trials; CONF4: Conflict 4 trials; CONF6: Conflict 6 trials). Uncorrected p-values: \*\*\**p* < 0.001, \*\**p* < 0.01, \**p* < 0.05.

## Tables

**Supplementary Table 1.**
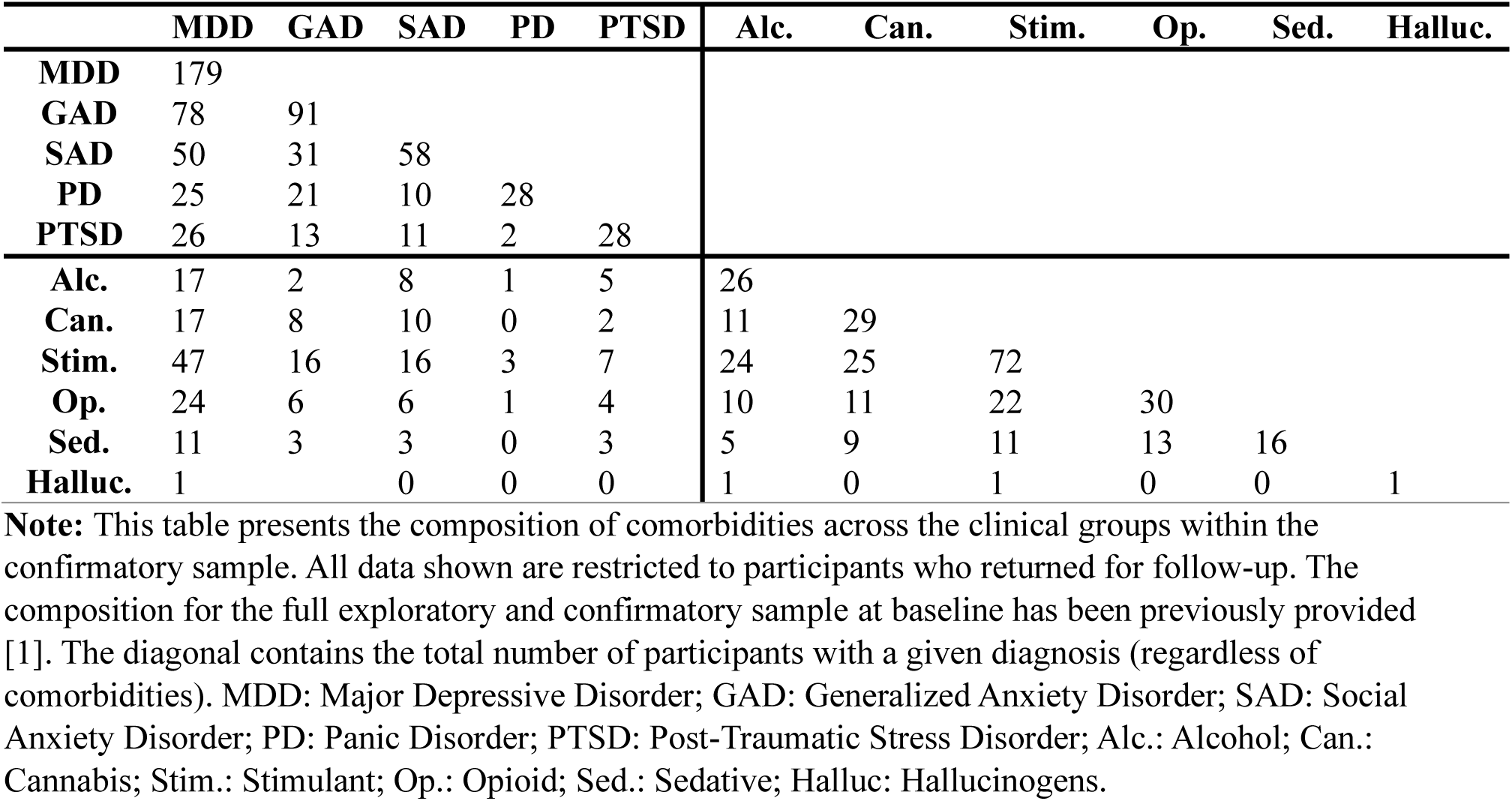
Comorbidity table for participants who returned for the follow-up visit.

**Supplementary Table 2.** Differences in participant characteristics at Baseline, between participants who returned at follow-up and those who did not.

|  | Returned<br>(N = 287) | Did Not Return<br>(N = 193) | *p* |
| --- | --- | --- | --- |
| Age | 33.50 (9.96) | 33.28 (9.53) | 0.806 |
| Sex (Male) | 74 (25.80) | 83 (43) | <0.001*** |
| PHQ | 8.14 (6.73) | 7.99 (6.20) | 0.801 |
| OASIS | 6.49 (4.89) | 6.52 (4.85) | 0.950 |
| DAST-10 | 2.35 (3.44) | 3.81 (4.01) | <0.001*** |
| WRAT reading score | 61.41 (6.40) | 60.05 (6.14) | 0.036* |

**Note:** PHQ = Patient Health Questionnaire; OASIS = Overall Anxiety Severity and Impairment Scale; DAST = Drug Abuse Screening Test
\**p*-values are based on t-tests for significant differences between the three groups (\*\*\**p* < 0.001; \*\**p* < 0.01; \**p* < 0.05). Sex is reported in terms of counts and percentages. *p*-values are based on associated Chi-Squared statistics.

**Supplementary Table 3.** Baseline symptom and demographic characteristics by group in those who did vs. did not return for follow-up.

|  | HCs |  | DEP/ANX |  | SUDs |  |
| --- | --- | --- | --- | --- | --- | --- |
|  | Returned | Did Not Return | Returned | Did Not Return | Returned | Did Not Return |
| N | 69 | 28 | 135 | 73 | 83 | 92 |
| Age | 33.45<br>(11.29) | 28.75<br>(10.06) | 33.63 (10.13) | 33.94<br>(10.32) | 33.34<br>(8.52) | 34.13 (8.37) |
| Sex (Male) | 23 (33.3) | 15 (53.6) | 29 (21.5) | 22 (30.1) | 22 (26.5)<sup>+</sup> | 46 (50.0)<sup>+</sup> |
| PHQ | 1.07 (1.75) | 1.61<br>(1.97) | 12.74 (5.08) | 12.14 (4.86) | 6.54 (5.81) | 6.64 (5.75) |
| OASIS | 0.96 (1.49) | 1.46<br>(1.88) | 9.77 (3.46) | 9.42 (3.82) | 5.77 (4.24) | 5.76 (4.67) |
| DAST | 0.19 (0.52) | 0.18<br>(0.39) | 0.44 (0.98) | 0.37 (0.89) | 7.25 (2.28) | 7.65 (2.18) |
| WRAT reading<br>score | 63.25<br>(5.63) | 61.76<br>(4.38) | 63.02 (5.03) | 62.31 (5.11) | 57.12<br>(7.12) | 56.38 (6.36) |

**Note:** PHQ = Patient Health Questionnaire; OASIS = Overall Anxiety Severity and Impairment Scale; DAST = Drug Abuse Screening Test;
<sup>+</sup>Sex is reported in terms of counts and percentages. *p*-values are based on associated Chi-Squared statistics.

**Supplementary Table 4.** Group-wise differences in participant characteristics at 1-year follow-up in the exploratory and confirmatory sample.

|  | HCs |  | DEP/ANX |  | SUDs |  |
| --- | --- | --- | --- | --- | --- | --- |
|  | *Exploratory* | *Confirmatory* | *Exploratory* | *Confirmatory* | *Exploratory* | *Confirmatory* |
| N | 48 | 69 | 192 | 135 | 84 | 83 |
| Age | 32.94<br>(11.29) | 34.50 (11.30) | 37.15<br>(11.40)* | 34.69<br>(10.17)* | 36.28 (9.18) | 34.41 (8.56) |
| Sex<br>(Male) | 24 (50.0) | 23 (33.3) | 50 (26.0) | 29 (21.5) | 34 (40.5) | 22 (26.5) |
| PHQ | 1.14 (1.84) | 0.91 (1.73) | 8.27 (6.10) | 8.47 (6.10) | 3.10 (4.57) | 3.30 (4.44) |
| OASIS | 1.47 (2.31) | 0.86 (1.75) | 7.56 (4.58) | 7.11 (4.) | 3.54 (4.36) | 3.26 (3.73) |
| DAST | 0.47 (0.56) | 0.30 (0.55) | 0.56 (1.05) | 0.37 (0.6) | 2.46 (2.89) | 2.27 (2.53) |
| WRAT<br>reading<br>score | 63.89 (4.54) | 63.25 (5.63) | 63.03 (4.64) | 63.02 (5.03) | 58.49 (6.00) | 57.12 (7.12) |

**Note:** PHQ = Patient Health Questionnaire; OASIS = Overall Anxiety Severity and Impairment Scale; DAST = Drug Abuse Screening Test;
\**p* < 0.05 based on t-test for significant differences between samples within each group.
<sup>+</sup> Sex is reported in terms of counts and percentages. *p*-values are based on associated Chi-Squared statistics.

**Supplementary Table 5.**
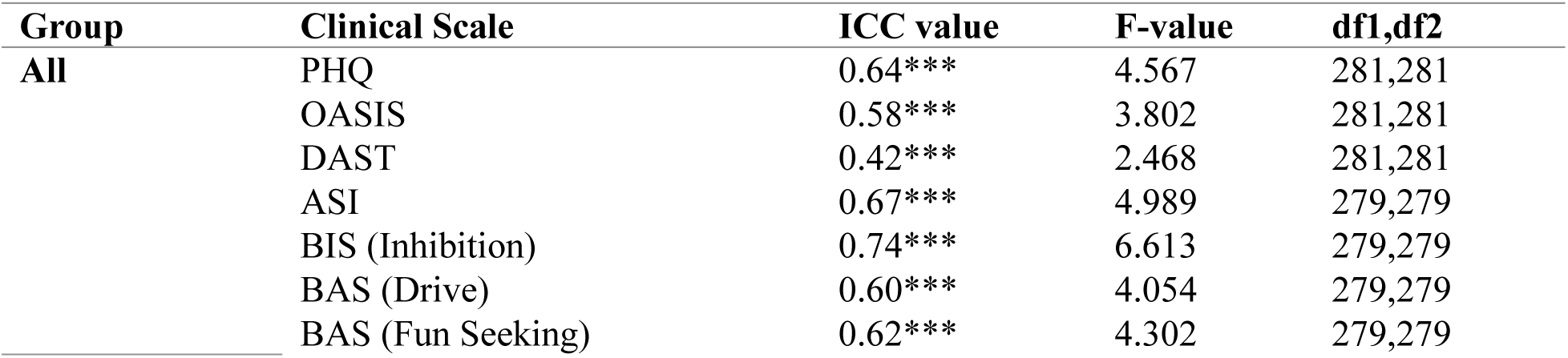
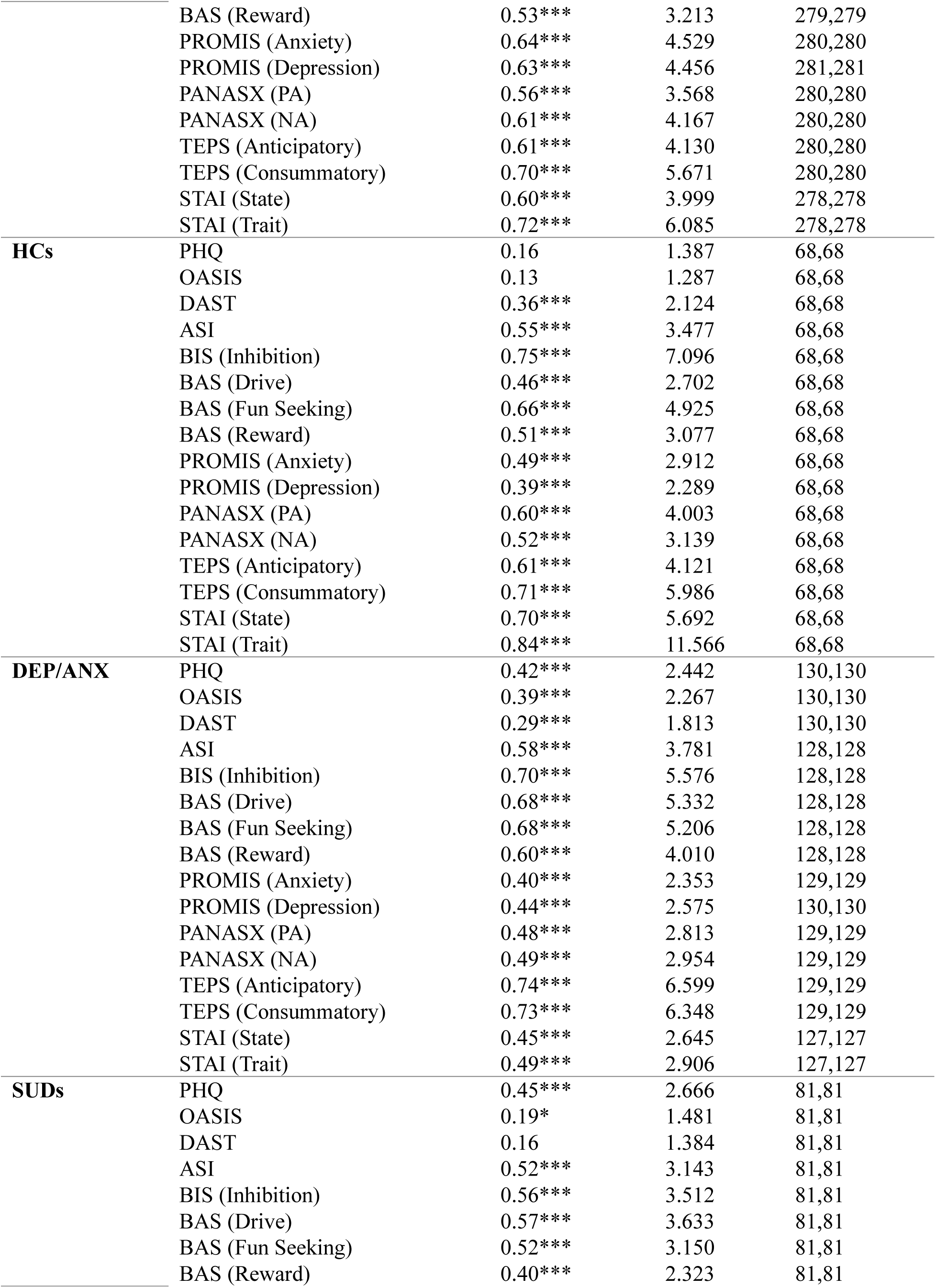
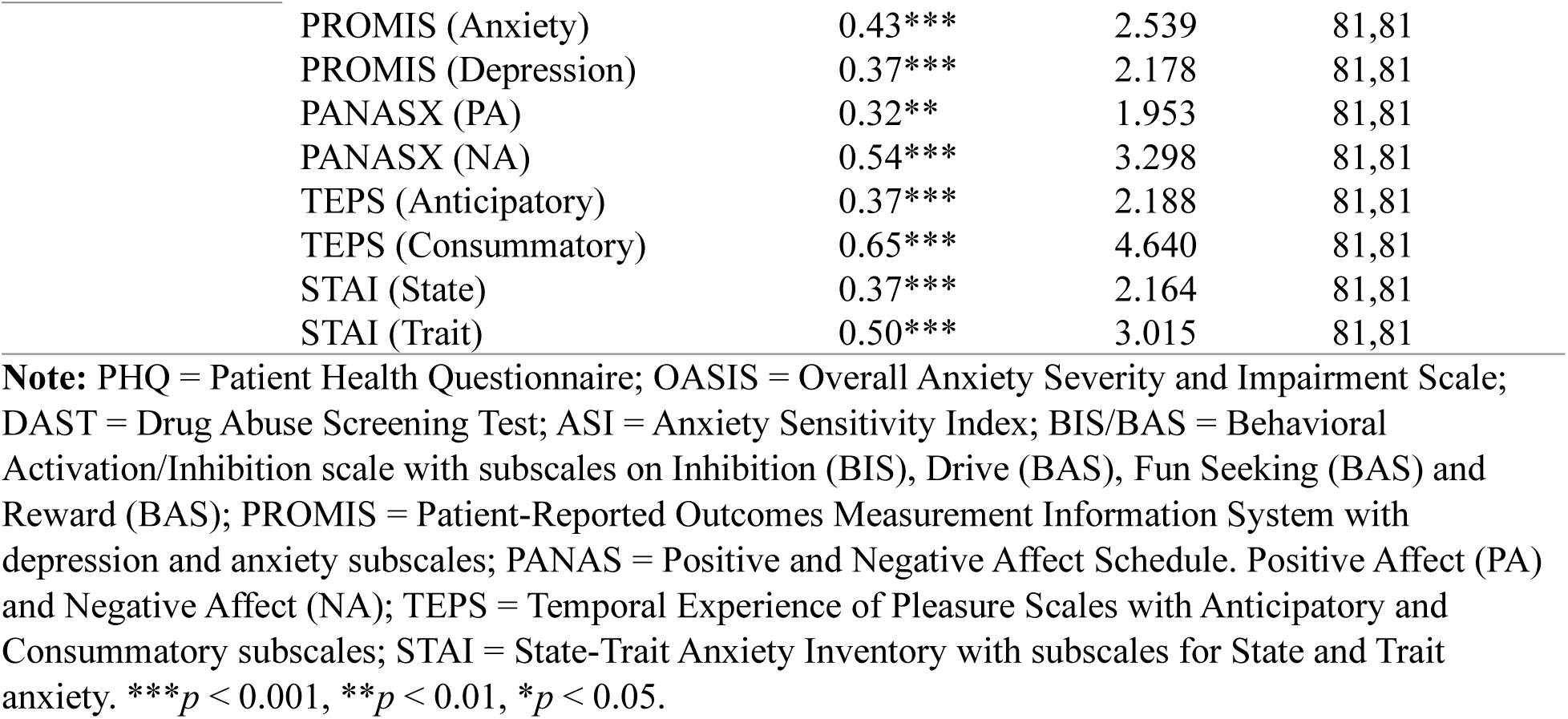
Group-wise interclass correlations for clinical measures at baseline and 1-year follow-up.

| Group | Clinical Scale | ICC value | F-value | df1,df2 |
| --- | --- | --- | --- | --- |
| All | PHQ | 0.64*** | 4.567 | 281,281 |
|  | OASIS | 0.58*** | 3.802 | 281,281 |
|  | DAST | 0.42*** | 2.468 | 281,281 |
|  | ASI | 0.67*** | 4.989 | 279,279 |
|  | BIS (Inhibition) | 0.74*** | 6.613 | 279,279 |
|  | BAS (Drive) | 0.60*** | 4.054 | 279,279 |
|  | BAS (Fun Seeking) | 0.62*** | 4.302 | 279,279 |
|  | BAS (Reward) | 0.53*** | 3.213 | 279,279 |
|  | PROMIS (Anxiety) | 0.64*** | 4.529 | 280,280 |
|  | PROMIS (Depression) | 0.63*** | 4.456 | 281,281 |
|  | PANASX (PA) | 0.56*** | 3.568 | 280,280 |
|  | PANASX (NA) | 0.61*** | 4.167 | 280,280 |
|  | TEPS (Anticipatory) | 0.61*** | 4.130 | 280,280 |
|  | TEPS (Consummatory) | 0.70*** | 5.671 | 280,280 |
|  | STAI (State) | 0.60*** | 3.999 | 278,278 |
|  | STAI (Trait) | 0.72*** | 6.085 | 278,278 |
| **HCs** | PHQ | 0.16 | 1.387 | 68,68 |
|  | OASIS | 0.13 | 1.287 | 68,68 |
|  | DAST | 0.36*** | 2.124 | 68,68 |
|  | ASI | 0.55*** | 3.477 | 68,68 |
|  | BIS (Inhibition) | 0.75*** | 7.096 | 68,68 |
|  | BAS (Drive) | 0.46*** | 2.702 | 68,68 |
|  | BAS (Fun Seeking) | 0.66*** | 4.925 | 68,68 |
|  | BAS (Reward) | 0.51*** | 3.077 | 68,68 |
|  | PROMIS (Anxiety) | 0.49*** | 2.912 | 68,68 |
|  | PROMIS (Depression) | 0.39*** | 2.289 | 68,68 |
|  | PANASX (PA) | 0.60*** | 4.003 | 68,68 |
|  | PANASX (NA) | 0.52*** | 3.139 | 68,68 |
|  | TEPS (Anticipatory) | 0.61*** | 4.121 | 68,68 |
|  | TEPS (Consummatory) | 0.71*** | 5.986 | 68,68 |
|  | STAI (State) | 0.70*** | 5.692 | 68,68 |
|  | STAI (Trait) | 0.84*** | 11.566 | 68,68 |
| **DEP/ANX** | PHQ | 0.42*** | 2.442 | 130,130 |
|  | OASIS | 0.39*** | 2.267 | 130,130 |
|  | DAST | 0.29*** | 1.813 | 130,130 |
|  | ASI | 0.58*** | 3.781 | 128,128 |
|  | BIS (Inhibition) | 0.70*** | 5.576 | 128,128 |
|  | BAS (Drive) | 0.68*** | 5.332 | 128,128 |
|  | BAS (Fun Seeking) | 0.68*** | 5.206 | 128,128 |
|  | BAS (Reward) | 0.60*** | 4.010 | 128,128 |
|  | PROMIS (Anxiety) | 0.40*** | 2.353 | 129,129 |
|  | PROMIS (Depression) | 0.44*** | 2.575 | 130,130 |
|  | PANASX (PA) | 0.48*** | 2.813 | 129,129 |
|  | PANASX (NA) | 0.49*** | 2.954 | 129,129 |
|  | TEPS (Anticipatory) | 0.74*** | 6.599 | 129,129 |
|  | TEPS (Consummatory) | 0.73*** | 6.348 | 129,129 |
|  | STAI (State) | 0.45*** | 2.645 | 127,127 |
|  | STAI (Trait) | 0.49*** | 2.906 | 127,127 |
| **SUDs** | PHQ | 0.45*** | 2.666 | 81,81 |
|  | OASIS | 0.19* | 1.481 | 81,81 |
|  | DAST | 0.16 | 1.384 | 81,81 |
|  | ASI | 0.52*** | 3.143 | 81,81 |
|  | BIS (Inhibition) | 0.56*** | 3.512 | 81,81 |
|  | BAS (Drive) | 0.57*** | 3.633 | 81,81 |
|  | BAS (Fun Seeking) | 0.52*** | 3.150 | 81,81 |
|  | BAS (Reward) | 0.40*** | 2.323 | 81,81 |

|  |  |  |  |
| --- | --- | --- | --- |
| PROMIS (Anxiety) | 0.43*** | 2.539 | 81,81 |
| PROMIS (Depression) | 0.37*** | 2.178 | 81,81 |
| PANASX (PA) | 0.32** | 1.953 | 81,81 |
| PANASX (NA) | 0.54*** | 3.298 | 81,81 |
| TEPS (Anticipatory) | 0.37*** | 2.188 | 81,81 |
| TEPS (Consummatory) | 0.65*** | 4.640 | 81,81 |
| STAI (State) | 0.37*** | 2.164 | 81,81 |
| STAI (Trait) | 0.50*** | 3.015 | 81,81 |

**Note:** PHQ = Patient Health Questionnaire; OASIS = Overall Anxiety Severity and Impairment Scale; DAST = Drug Abuse Screening Test; ASI = Anxiety Sensitivity Index; BIS/BAS = Behavioral Activation/Inhibition scale with subscales on Inhibition (BIS), Drive (BAS), Fun Seeking (BAS) and Reward (BAS); PROMIS = Patient-Reported Outcomes Measurement Information System with depression and anxiety subscales; PANAS = Positive and Negative Affect Schedule. Positive Affect (PA) and Negative Affect (NA); TEPS = Temporal Experience of Pleasure Scales with Anticipatory and Consummatory subscales; STAI = State-Trait Anxiety Inventory with subscales for State and Trait anxiety. \*\*\* $p < 0.001$ , \*\* $p < 0.01$ , \* $p < 0.05$ .

**Supplementary Table 6.**
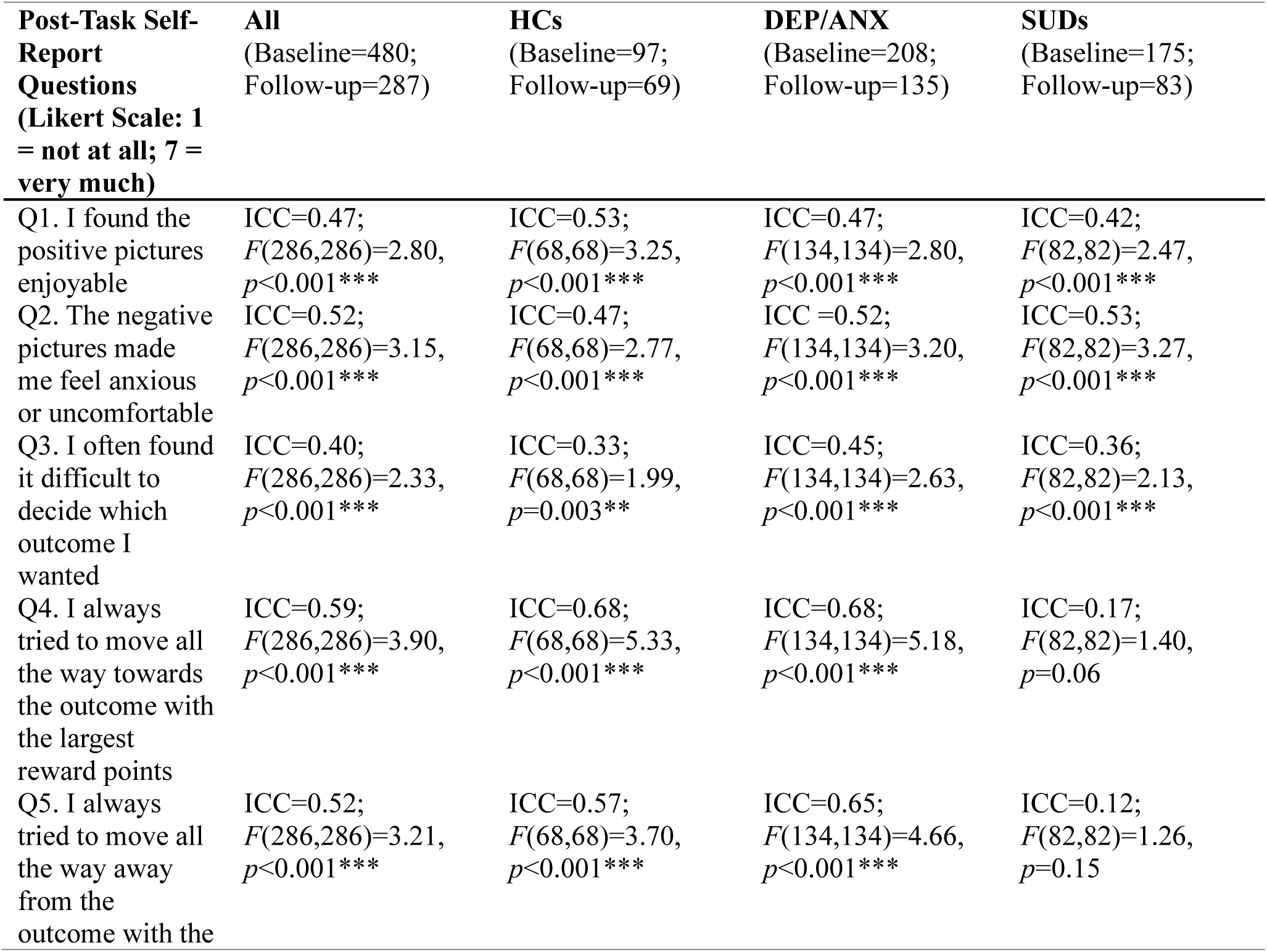
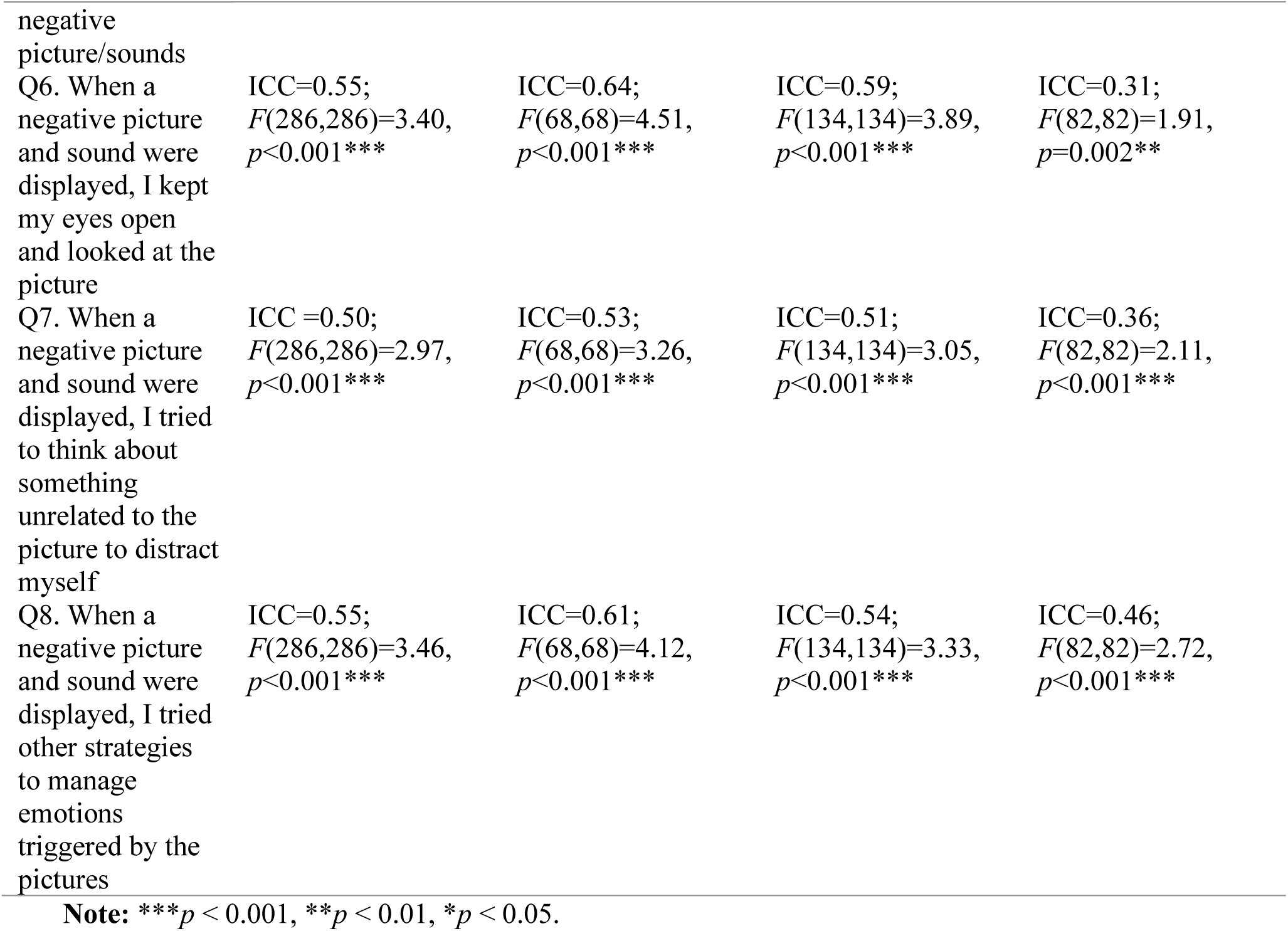
Group-wise intra-class correlations for post-task survey at baseline and 1-year follow-up.

**Supplementary Table 7.** Group-wise post-task self-report questionnaire items at baseline and follow-up, and correlations with computational model parameters at follow-up.

| Post-Task Self-Report<br>Questions (Likert Scale: 1 = not<br>at all; 7 = very much) | Mean (SD) Baseline |  | Mean (SD)<br>1-Year<br>follow-up | Emotion<br>conflict<br>(EC) | Decision<br>uncertainty<br>(β) |
| --- | --- | --- | --- | --- | --- |
|  | All<br>participants | Returned for<br>follow-up |  |  |  |
| HCs (N<sub>All</sub>= 97; N<sub>Follow-up</sub>=69) |  |  |  |  |  |
| Q1. I found the positive pictures<br>enjoyable | 5.31 (1.39) | 5.51 (1.30) | 5.20 (1.62) | 0.23 | -0.01 |
| Q2. The negative pictures made<br>me feel anxious or uncomfortable | 4.01 (2.01) | 3.99 (1.94) | 4.04 (1.92) | 0.47*** | -0.21 |
| Q3. I often found it difficult to<br>decide which outcome I wanted | 2.11 (1.66) | 2.16 (1.63) | 1.86 (1.29) | -0.05 | 0.51*** |
| Q4. I always tried to move all the<br>way towards the outcome with the<br>largest reward points | 4.34 (2.55) | 4.10 (2.51) | 4.38 (2.56) | -0.89*** | -0.33** |
| Q5. I always tried to move all the<br>way away from the outcome with<br>the negative picture/sounds | 3.24 (2.37) | 3.36 (2.35) | 3.30 (2.51) | 0.75*** | 0.12 |
| Q6. When a negative picture and sound were displayed, I kept my eyes open and looked at the picture | 5.13 (2.01) | 5.16 (1.96) | 4.91 (2.09) | -0.42*** | -0.08 |
| Q7. When a negative picture and sound were displayed, I tried to think about something unrelated to the picture to distract myself | 3.14 (2.07) | 3.16 (2.06) | 3.41 (2.00) | 0.46*** | 0.10 |
| Q8. When a negative picture and sound were displayed, I tried other strategies to manage emotions triggered by the pictures | 3.28 (1.92) | 3.45 (1.91) | 3.62 (1.93) | 0.46*** | -0.05 |
| DEP/ANX (N<sub>All</sub>= 208; N<sub>Follow-up</sub>=135) |  |  |  |  |  |
| Q1. I found the positive pictures enjoyable | 5.05 (1.53) | 5.12 (1.56) | 5.09 (1.51) | 0.09 | -0.13 |
| Q2. The negative pictures made me feel anxious or uncomfortable | 4.18 (1.96) | 4.13 (2.03) | 4.25 (1.94) | 0.32*** | 0.04 |
| Q3. I often found it difficult to decide which outcome I wanted | 2.31 (1.71) | 2.27 (1.70) | 2.12 (1.67) | 0.01 | 0.44*** |
| Q4. I always tried to move all the way towards the outcome with the largest reward points | 4.70 (2.40) | 4.73 (2.43) | 4.90 (2.52) | -0.83*** | -0.48*** |
| Q5. I always tried to move all the way away from the outcome with the negative picture/sounds | 3.12 (2.25) | 3.10 (2.27) | 2.99 (2.35) | 0.79*** | 0.33*** |
| Q6. When a negative picture and sound were displayed, I kept my eyes open and looked at the picture | 5.25 (1.92) | 5.23 (2.00) | 5.28 (1.97) | -0.43*** | -0.29*** |
| Q7. When a negative picture and sound were displayed, I tried to think about something unrelated to the picture to distract myself | 3.15 (1.93) | 3.13 (1.93) | 3.14 (2.03) | 0.32*** | 0.07 |
| Q8. When a negative picture and sound were displayed, I tried other strategies to manage emotions triggered by the pictures | 3.37 (1.94) | 3.30 (1.91) | 3.32 (1.93) | 0.32*** | -0.01 |
| SUDs (N<sub>All</sub>= 175; N<sub>Follow-up</sub>=83) |  |  |  |  |  |
| Q1. I found the positive pictures enjoyable | 4.83 (1.66) | 4.99 (1.71) | 4.59 (1.64) | -0.14 | -0.12 |
| Q2. The negative pictures made me feel anxious or uncomfortable | 3.80 (1.96) | 3.73 (1.94) | 3.54 (1.94) | 0.16 | -0.18 |
| Q3. I often found it difficult to decide which outcome I wanted | 2.45 (1.75) | 2.34 (1.71) | 2.17 (1.73) | 0.02 | 0.36*** |
| Q4. I always tried to move all the way towards the outcome with the largest reward points | 5.36 (2.08) | 5.63 (1.94) | 5.63 (2.05) | -0.73*** | -0.6*** |
| Q5. I always tried to move all the way away from the outcome with the negative picture/sounds | 2.33 (1.79) | 2.27 (1.76) | 2.51 (2.06) | 0.53*** | 0.24* |
| Q6. When a negative picture and sound were displayed, I kept my eyes open and looked at the picture | 5.82 (1.72) | 5.86 (1.59) | 5.61 (1.83) | -0.26* | -0.12 |
| Q7. When a negative picture and sound were displayed, I tried to think about something unrelated to the picture to distract myself | 2.29 (1.65) | 2.41 (1.80) | 2.27 (1.65) | 0.2 | 0 |
| Q8. When a negative picture and sound were displayed, I tried other strategies to manage emotions triggered by the pictures | 2.51 (1.76) | 2.53 (1.79) | 2.71 (1.84) | 0.09 | -0.09 |

**Note:** \*\*\* $p < 0.001$ , \*\* $p < 0.01$ , \* $p < 0.05$ .

**Supplementary Table 8.** Results of linear mixed effects models predicting DU and EC in data including all baseline participants, when accounting for effects of group and time.

| Predictor* | Result | EMM | Posthoc |
| --- | --- | --- | --- |
| **Decision Uncertainty (DU)** |  |  |  |
| Group | $F(2, 493)=1.44, p=0.239$ | | NS |
| Time | $F(1, 337)=24.16, p<0.001$ | T1=1.022;<br>T2=0.745 | T1 - T2: $t(340.3)=5.296, p<0.001, d=0.43$ |
| Group x Time | $F(2, 337)=1.60, p=0.203$ | | NS |
| **Emotion Conflict (EC)** |  |  |  |
| Group | $F(2, 495)=20.64, p<0.001$ | D/A=0.861;<br>HCs=1.022;<br>SUDs=0.465 | D/A - HCs: $t(458.3)=-1.826, p=0.068, d=-0.36$<br>D/A - SUDs: $t(485.37)=5.28, p<0.001, d=0.9$<br>HCs - SUDs: $t(472.44)=6.072, p<0.001, d=1.26$ |
| Time | $F(1, 325)=0.50, p=0.478$ | | NS |
| Group x Time | $F(2, 325)=0.48, p=0.620$ | | NS |

**Note:** HCs = Healthy Comparisons; D/A = Depression and Anxiety; SUDs = Substance Use Disorder; T1 = Baseline; T2 = Follow-up; NS = nonsignificant.
For interpretability, sum coding was used for time (baseline = -1; follow-up = 1), and group (with HCs coded as -1).

**Supplementary Table 9.**
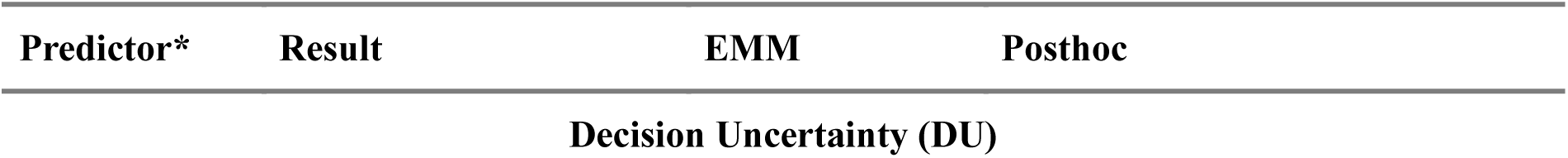
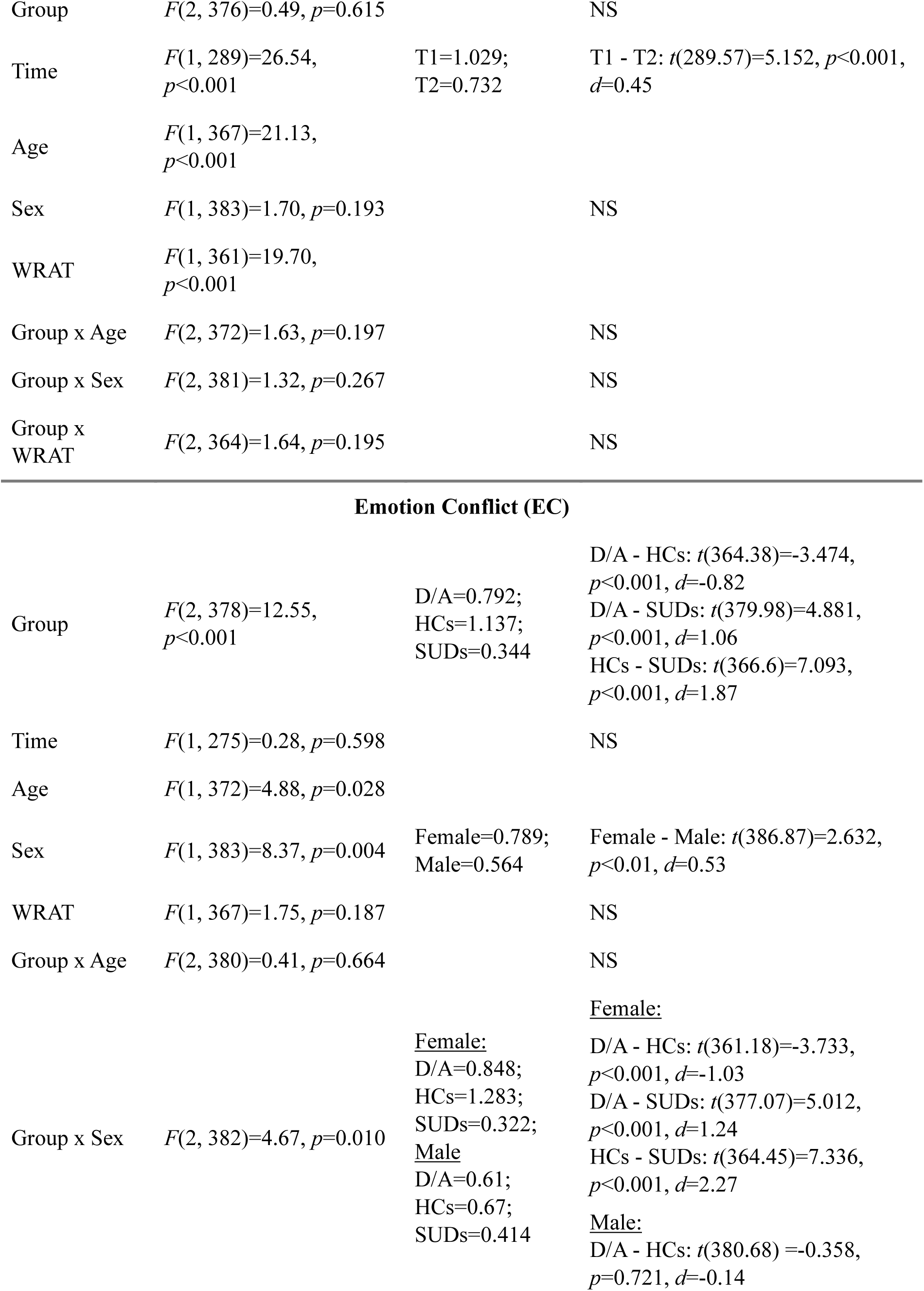
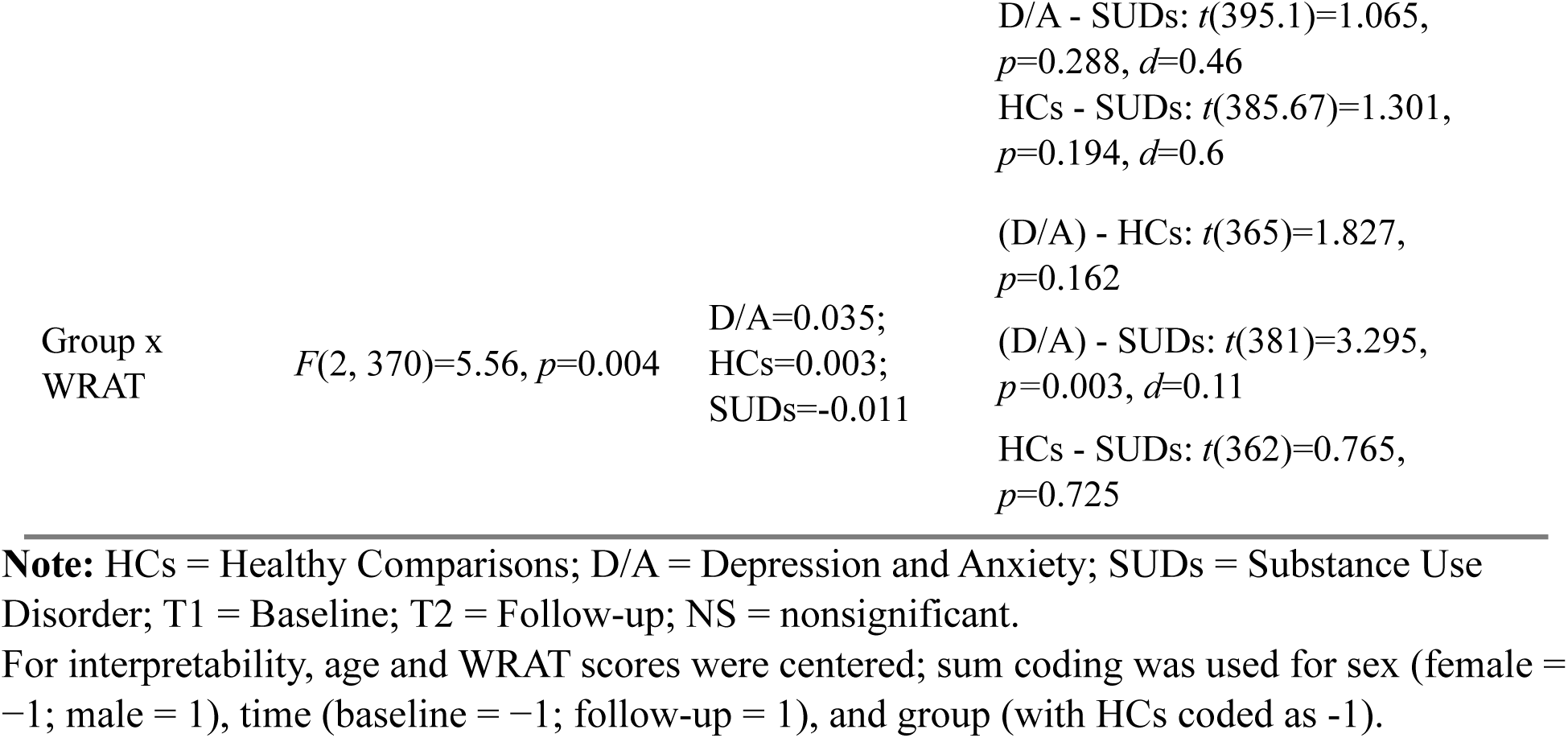
Results of Linear mixed effects models predicting DU and EC in data including all baseline participants, when accounting for effects of group, time, age, sex and WRAT scores.

**Supplementary Table 10.** Summary statistics for response times (Mean (SD)) at baseline and follow-up.

| Full Sample | HCs<br>(N = 97) | DEP/ANX<br>(N = 208) | SUDs<br>(N = 175) | $p^*$ |
| --- | --- | --- | --- | --- |
| **All trials** |  |  |  |  |
| Baseline | 1.24 (0.27) | 1.25 (0.29) | 1.30 (0.33) | 0.169 |
| Follow-up | 1.19 (0.30) | 1.19 (0.29) | 1.23 (0.32) | 0.598 |
| **Approach** |  |  |  |  |
| Baseline | 1.20 (0.30) | 1.22 (0.35) | 1.29 (0.40) | 0.051 |
| Follow-up | 1.15 (0.33) | 1.16 (0.33) | 1.19 (0.34) | 0.735 |
| **Avoid** |  |  |  |  |
| Baseline | 1.30 (0.35) | 1.35 (0.31) | 1.51 (0.35) | <0.001*** |
| Follow-up | 1.29 (0.40) | 1.30 (0.35) | 1.45 (0.32) | 0.007** |
| **CONF 2** |  |  |  |  |
| Baseline | 1.25 (0.31) | 1.25 (0.34) | 1.26 (0.38) | 0.886 |
| Follow-up | 1.17 (0.33) | 1.18 (0.32) | 1.19 (0.38) | 0.930 |
| **CONF 4** |  |  |  |  |
| Baseline | 1.19 (0.31) | 1.22 (0.37) | 1.21 (0.36) | 0.831 |
| Follow-up | 1.14 (0.29) | 1.16 (0.33) | 1.16 (0.37) | 0.938 |
| **CONF 6** |  |  |  |  |
| Baseline | 1.24 (0.30) | 1.23 (0.33) | 1.22 (0.37) | 0.945 |
| Follow-up | 1.18 (0.32) | 1.17 (0.33) | 1.17 (0.36) | 0.968 |
| **Participants who Returned for Follow-up** | **HCs**<br>**(N=69)** | **DEP/ANX**<br>**(N=135)** | **SUDs**<br>**(N=83)** | $p^*$ |
| **All trials** |  |  |  |  |
| Baseline | 1.24 (0.27) | 1.25 (0.28) | 1.28 (0.35) | 0.611 |
| Follow-up | 1.19 (0.30) | 1.19 (0.29) | 1.23 (0.32) | 0.598 |
| **Approach** |  |  |  |  |
| Baseline | 1.20 (0.30) | 1.21 (0.33) | 1.29 (0.45) | 0.193 |
| Follow-up | 1.15 (0.32) | 1.16 (0.33) | 1.19 (0.34) | 0.735 |
| **Avoid** |  |  |  |  |
| Baseline | 1.27 (0.34) | 1.37 (0.31) | 1.51 (0.38) | <0.001*** |
| Follow-up | 1.29 (0.40) | 1.30 (0.35) | 1.45 (0.32) | 0.007** |
| **CONF 2** |  |  |  |  |
| Baseline | 1.26 (0.33) | 1.24 (0.34) | 1.23 (0.37) | 0.872 |
| Follow-up | 1.17 (0.33) | 1.18 (0.32) | 1.19 (0.38) | 0.930 |
| **CONF 4** |  |  |  |  |
| Baseline | 1.21 (0.34) | 1.22 (0.37) | 1.19 (0.37) | 0.798 |
| Follow-up | 1.14 (0.29) | 1.16 (0.33) | 1.16 (0.37) | 0.938 |
| **CONF 6** |  |  |  |  |
| Baseline | 1.24 (0.30) | 1.21 (0.29) | 1.19(0.39) | 0.679 |
| Follow-up | 1.18 (0.32) | 1.17 (0.33) | 1.17 (0.36) | 0.968 |

**Note:** \*\*\* $p < 0.001$ , \*\* $p < 0.01$ , \* $p < 0.05$ .

**Supplementary Table 11.**
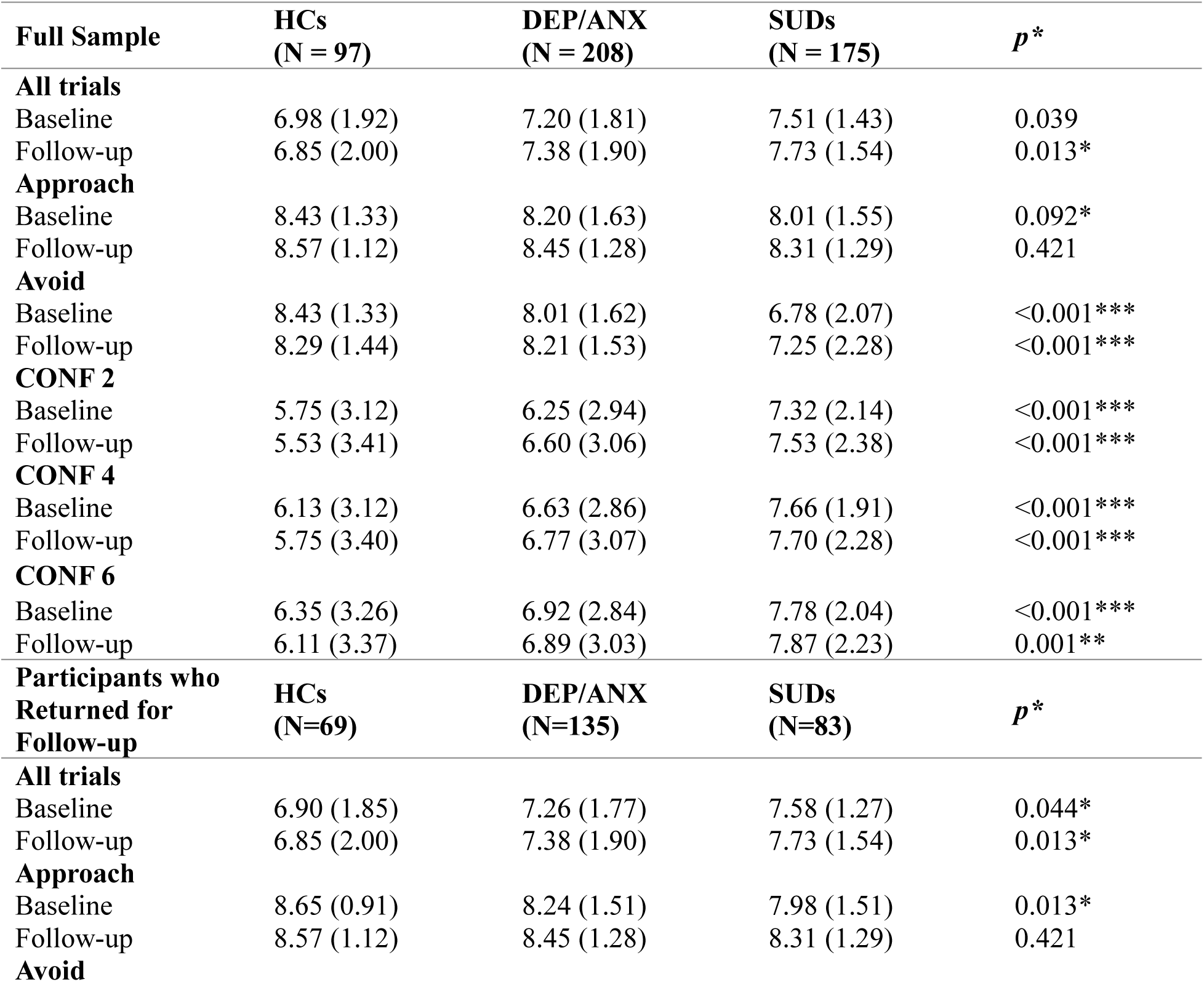
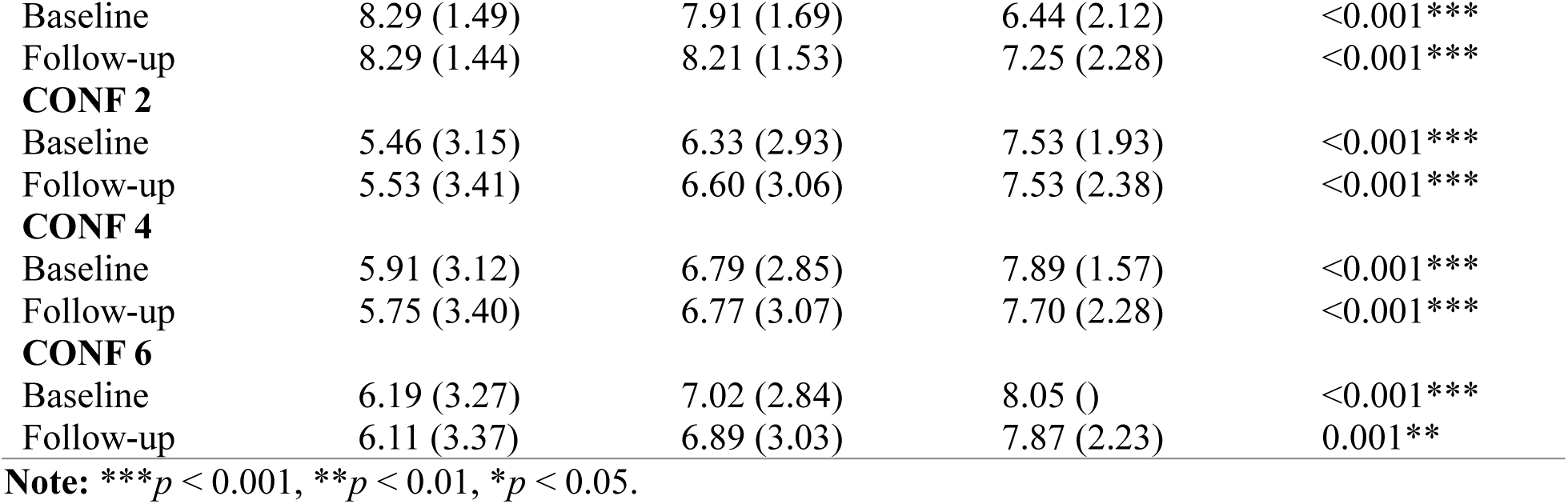
Summary statistics for average chosen runway positions (Mean (SD)) at baseline and follow-up.

| **Full Sample** | **HCs**<br>**(N = 97)** | **DEP/ANX**<br>**(N = 208)** | **SUDs**<br>**(N = 175)** | **$p^*$** |
| --- | --- | --- | --- | --- |
| **All trials** |  |  |  |  |
| Baseline | 6.98 (1.92) | 7.20 (1.81) | 7.51 (1.43) | 0.039 |
| Follow-up | 6.85 (2.00) | 7.38 (1.90) | 7.73 (1.54) | 0.013* |
| **Approach** |  |  |  |  |
| Baseline | 8.43 (1.33) | 8.20 (1.63) | 8.01 (1.55) | 0.092* |
| Follow-up | 8.57 (1.12) | 8.45 (1.28) | 8.31 (1.29) | 0.421 |
| **Avoid** |  |  |  |  |
| Baseline | 8.43 (1.33) | 8.01 (1.62) | 6.78 (2.07) | <0.001*** |
| Follow-up | 8.29 (1.44) | 8.21 (1.53) | 7.25 (2.28) | <0.001*** |
| **CONF 2** |  |  |  |  |
| Baseline | 5.75 (3.12) | 6.25 (2.94) | 7.32 (2.14) | <0.001*** |
| Follow-up | 5.53 (3.41) | 6.60 (3.06) | 7.53 (2.38) | <0.001*** |
| **CONF 4** |  |  |  |  |
| Baseline | 6.13 (3.12) | 6.63 (2.86) | 7.66 (1.91) | <0.001*** |
| Follow-up | 5.75 (3.40) | 6.77 (3.07) | 7.70 (2.28) | <0.001*** |
| **CONF 6** |  |  |  |  |
| Baseline | 6.35 (3.26) | 6.92 (2.84) | 7.78 (2.04) | <0.001*** |
| Follow-up | 6.11 (3.37) | 6.89 (3.03) | 7.87 (2.23) | 0.001** |
| **Participants who**<br>**Returned for**<br>**Follow-up** | **HCs**<br>**(N=69)** | **DEP/ANX**<br>**(N=135)** | **SUDs**<br>**(N=83)** | **$p^*$** |
| **All trials** |  |  |  |  |
| Baseline | 6.90 (1.85) | 7.26 (1.77) | 7.58 (1.27) | 0.044* |
| Follow-up | 6.85 (2.00) | 7.38 (1.90) | 7.73 (1.54) | 0.013* |
| **Approach** |  |  |  |  |
| Baseline | 8.65 (0.91) | 8.24 (1.51) | 7.98 (1.51) | 0.013* |
| Follow-up | 8.57 (1.12) | 8.45 (1.28) | 8.31 (1.29) | 0.421 |
| **Avoid** |  |  |  |  |
| Baseline | 8.29 (1.49) | 7.91 (1.69) | 6.44 (2.12) | <0.001*** |
| Follow-up | 8.29 (1.44) | 8.21 (1.53) | 7.25 (2.28) | <0.001*** |
| **CONF 2** |  |  |  |  |
| Baseline | 5.46 (3.15) | 6.33 (2.93) | 7.53 (1.93) | <0.001*** |
| Follow-up | 5.53 (3.41) | 6.60 (3.06) | 7.53 (2.38) | <0.001*** |
| **CONF 4** |  |  |  |  |
| Baseline | 5.91 (3.12) | 6.79 (2.85) | 7.89 (1.57) | <0.001*** |
| Follow-up | 5.75 (3.40) | 6.77 (3.07) | 7.70 (2.28) | <0.001*** |
| **CONF 6** |  |  |  |  |
| Baseline | 6.19 (3.27) | 7.02 (2.84) | 8.05 () | <0.001*** |
| Follow-up | 6.11 (3.37) | 6.89 (3.03) | 7.87 (2.23) | 0.001** |

**Note:** \*\*\* $p < 0.001$ , \*\* $p < 0.01$ , \* $p < 0.05$ .

**Supplementary Table 12.**
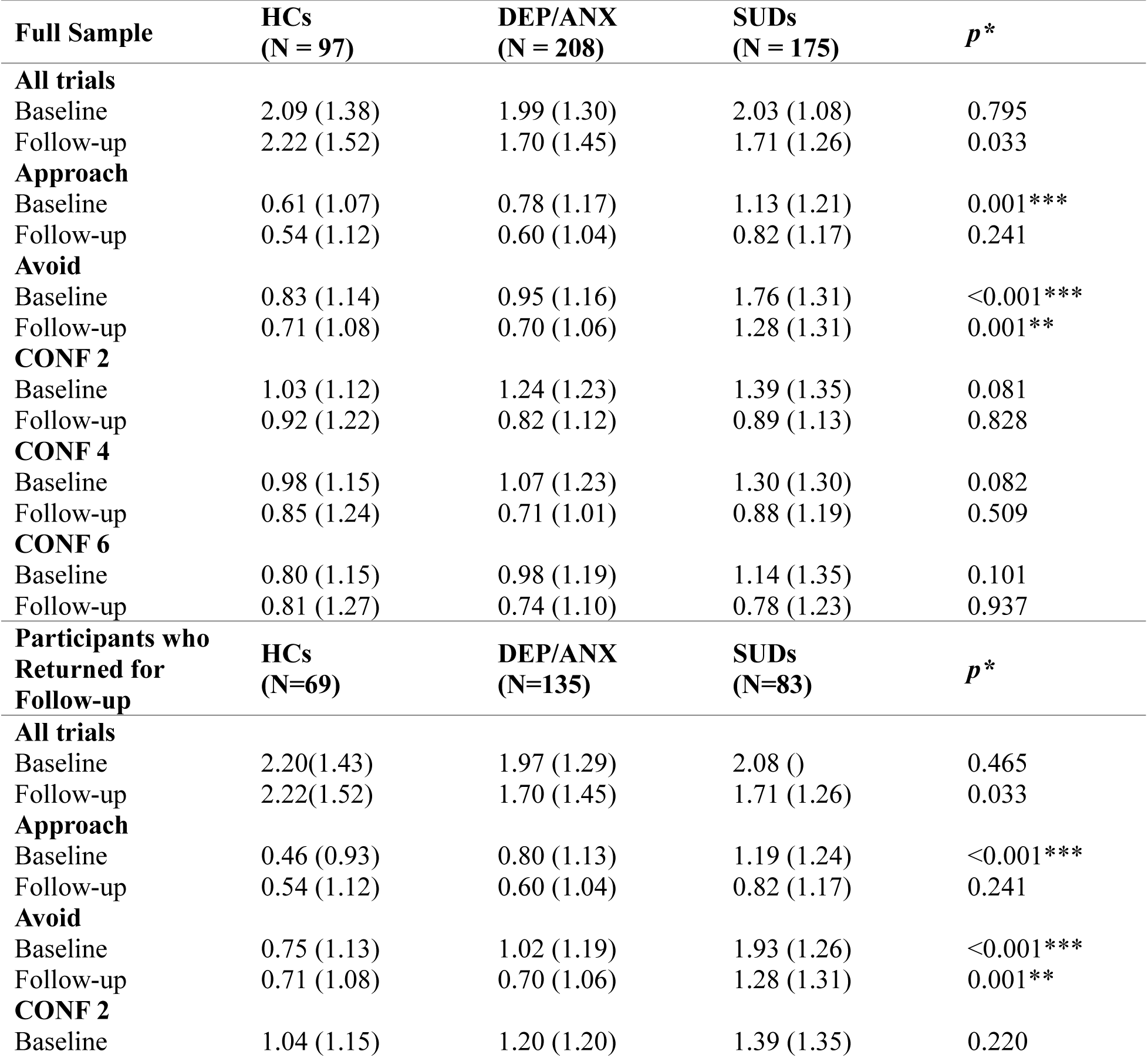
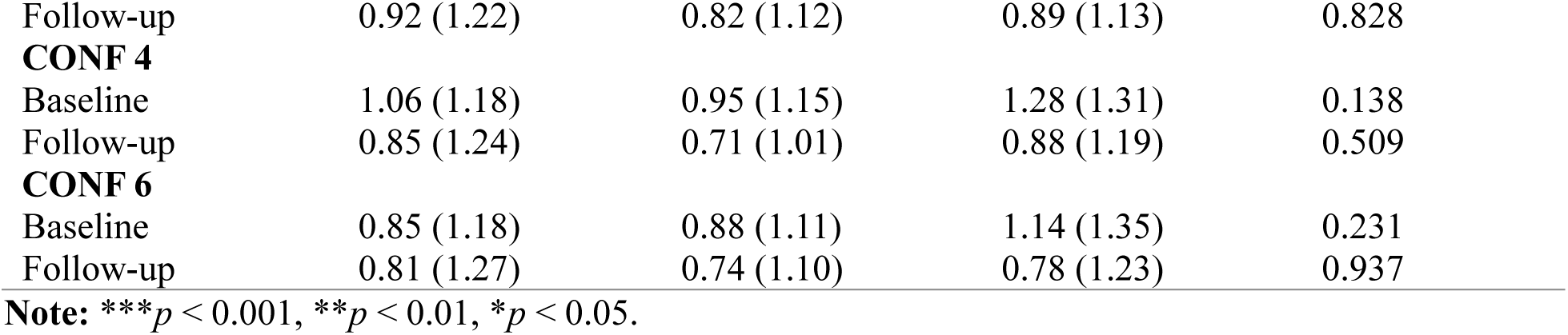
Summary statistics for variability (SD) in chosen runway positions (Mean (SD)) at baseline and follow-up.

**Supplementary Table 13.** Results of Linear mixed effects models predicting response time (RTs) in participants who returned for follow-up visit, when accounting for effects of group, time, and their interaction.

| Predictor* | Group | Time | Group x Time |
| --- | --- | --- | --- |
| **All Trials** | $F(2, 283)=0.64, p=0.526$ | $F(1, 283)=9.33, p=0.002$<br>T1=1.257; T2=1.203<br>T1 - T2: $t(283.21)=3.055, p=0.002, d=0.27$ | $F(2, 283)=0.02, p=0.981$ |
| **Approach** | $F(2, 283)=1.16, p=0.315$ | $F(1, 283)=11.22, p<0.001$<br>T1=1.235; T2=1.164<br>T1 - T2: $t(283.23)=3.35, p<0.001, d=0.29$ | $F(2, 283)=0.68, p=0.507$ |
| **Avoid** | $F(2, 283)=9.54, p<0.001$<br>D/A=1.336; HCs=1.282;<br>SUDs=1.48<br>(D/A) - HCs: $t(283.37)=1.218, p=0.224, d=0.21$<br>(D/A) - SUDs: $t(284.23)=-3.478, p<0.001, d=-0.58$<br>HCs - SUDs: $t(283.7)=-4.086, p<0.001, d=-0.79$ | $F(1, 282)=2.89, p=0.090$ | $F(2, 282)=1.32, p=0.269$ |
| **CONF2** | $F(2, 283)=0.01, p=0.992$ | $F(1, 283)=8.15, p=0.005$<br>T1=1.243; T2=1.183<br>T1 - T2: $t(283.24)=2.855, p=0.005, d=0.25$ | $F(2, 283)=0.39, p=0.678$ |
| **CONF4** | $F(2, 283)=0.09, p=0.912$ | $F(1, 283)=6.64, p=0.010$<br>T1=1.207; T2=1.152<br>T1 - T2: $t(283.23)=2.577, p=0.010, d=0.22$ | $F(2, 283)=0.33, p=0.723$ |
| **CONF6** | $F(2, 283)=0.20, p=0.822$ | $F(1, 283)=3.73, p=0.055$ | $F(2, 283)=0.22, p=0.804$ |

**Note:** HCs = Healthy Comparisons; D/A = Depression and Anxiety; SUDs = Substance Use Disorder; T1 = Baseline; T2 = Follow-up; NS = nonsignificant.
For interpretability, sum coding was used for time (baseline = -1; follow-up = 1), and group (with HCs coded as -1).
\*\*\* $p < 0.001$ , \*\* $p < 0.01$ , \* $p < 0.05$ .

**Supplementary Table 14.**
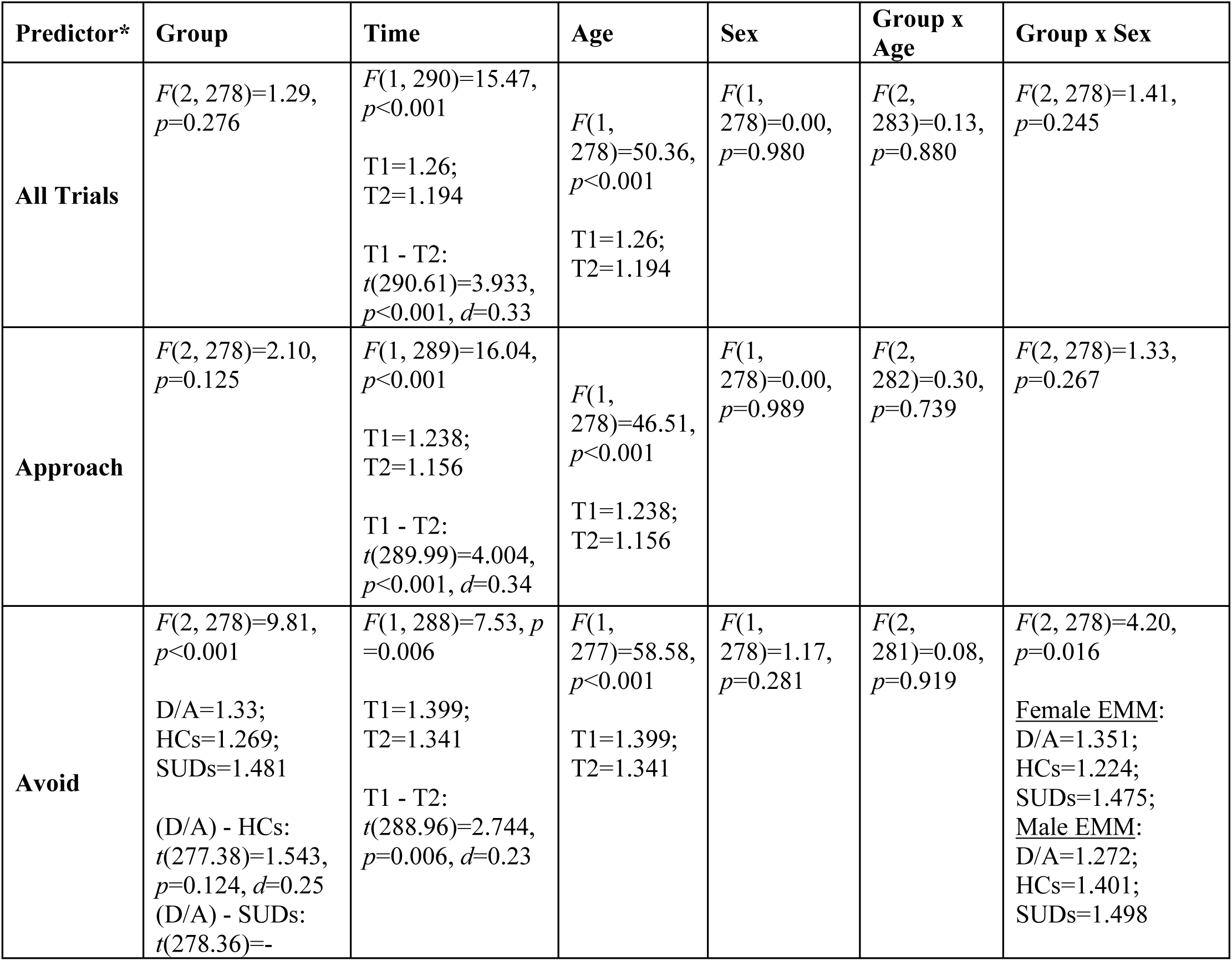
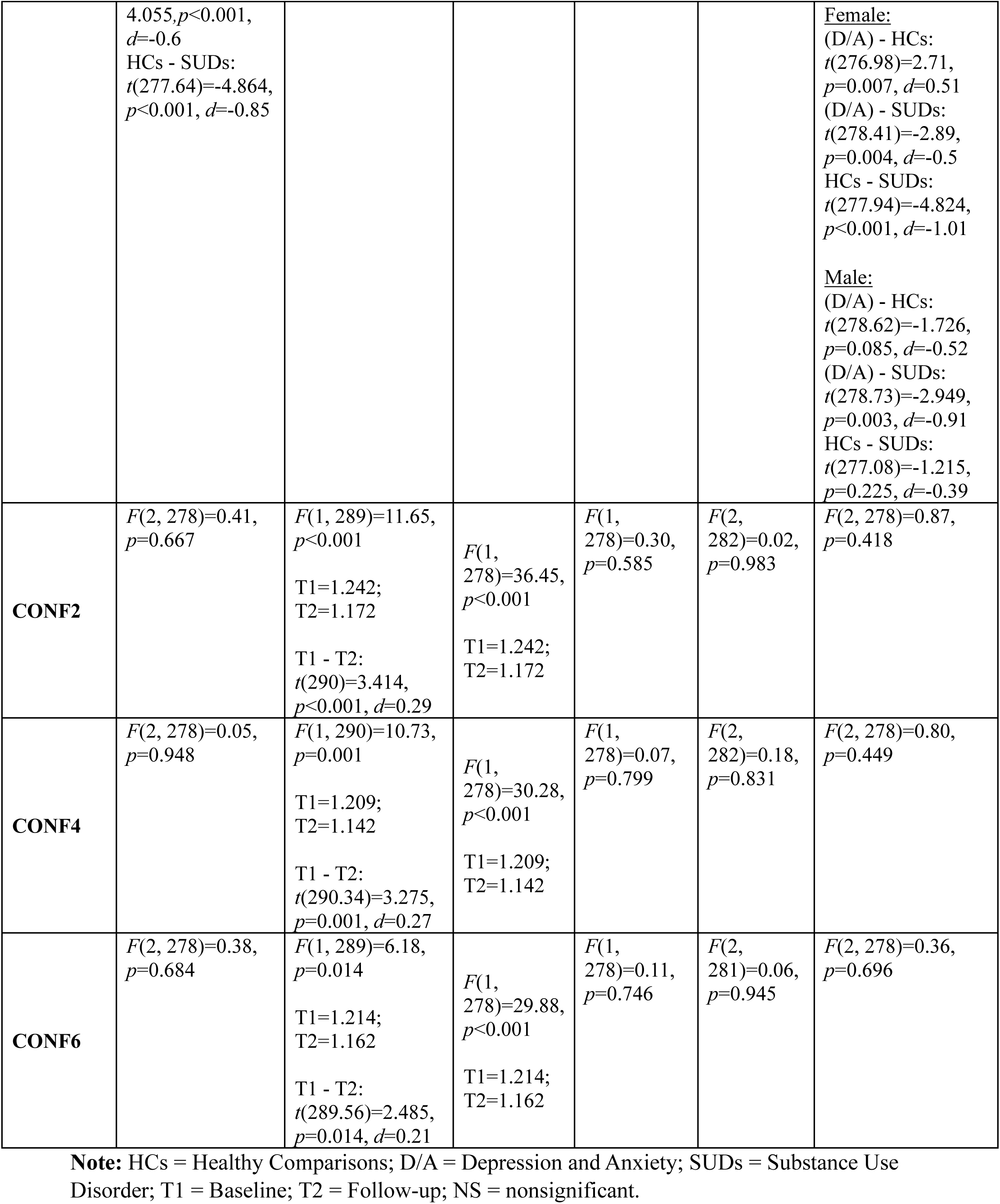
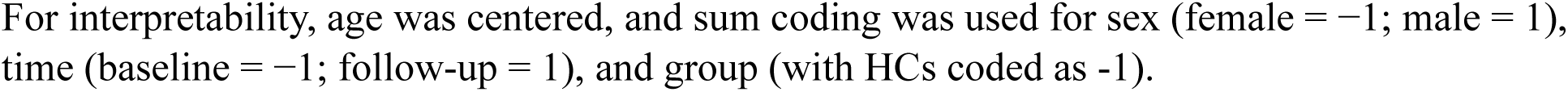
Results of Linear mixed effects models predicting response time (RTs) in participants who returned for follow-up visit, when accounting for effects of group, time, age and sex.

**Supplementary Table 15.**
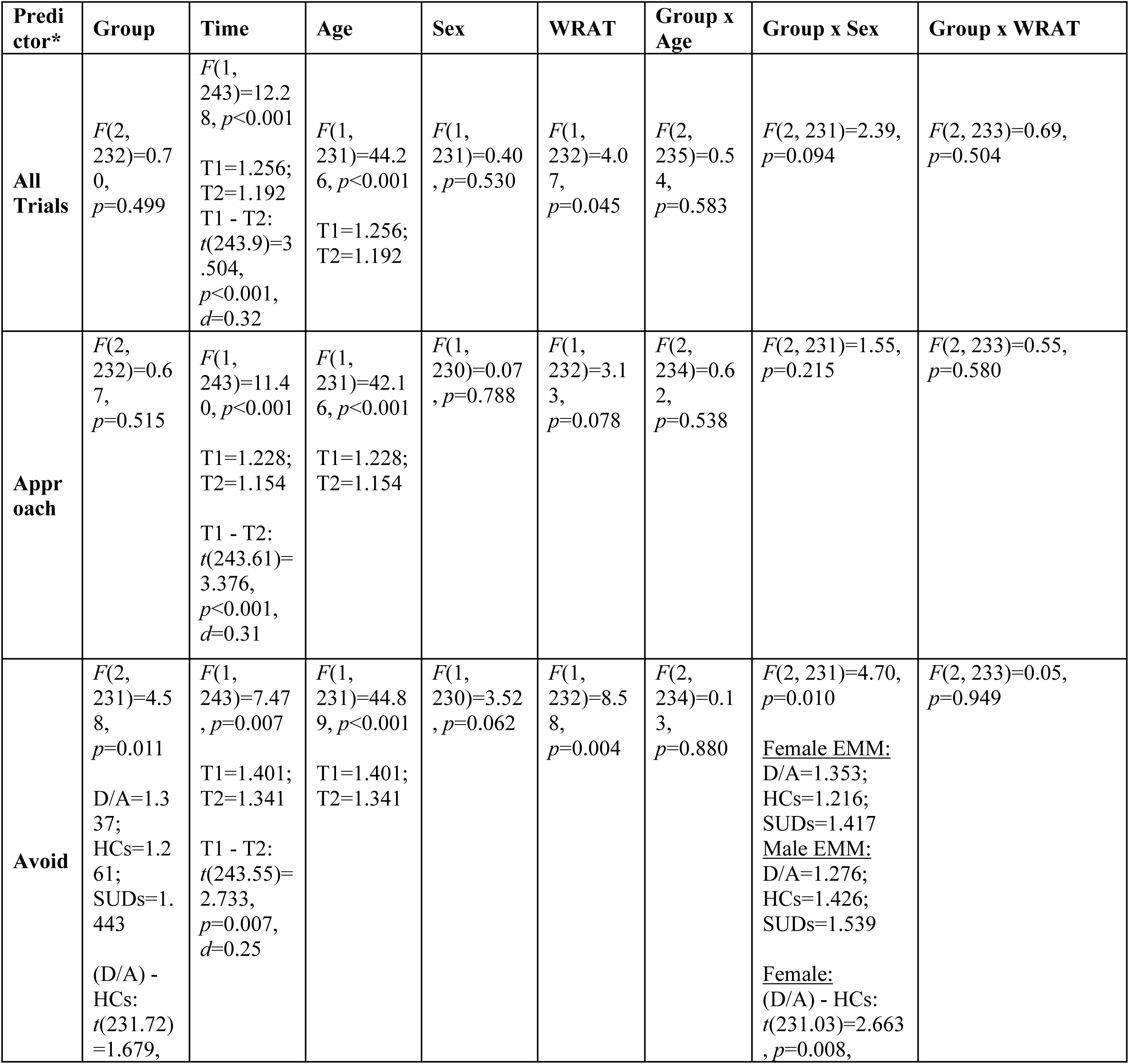
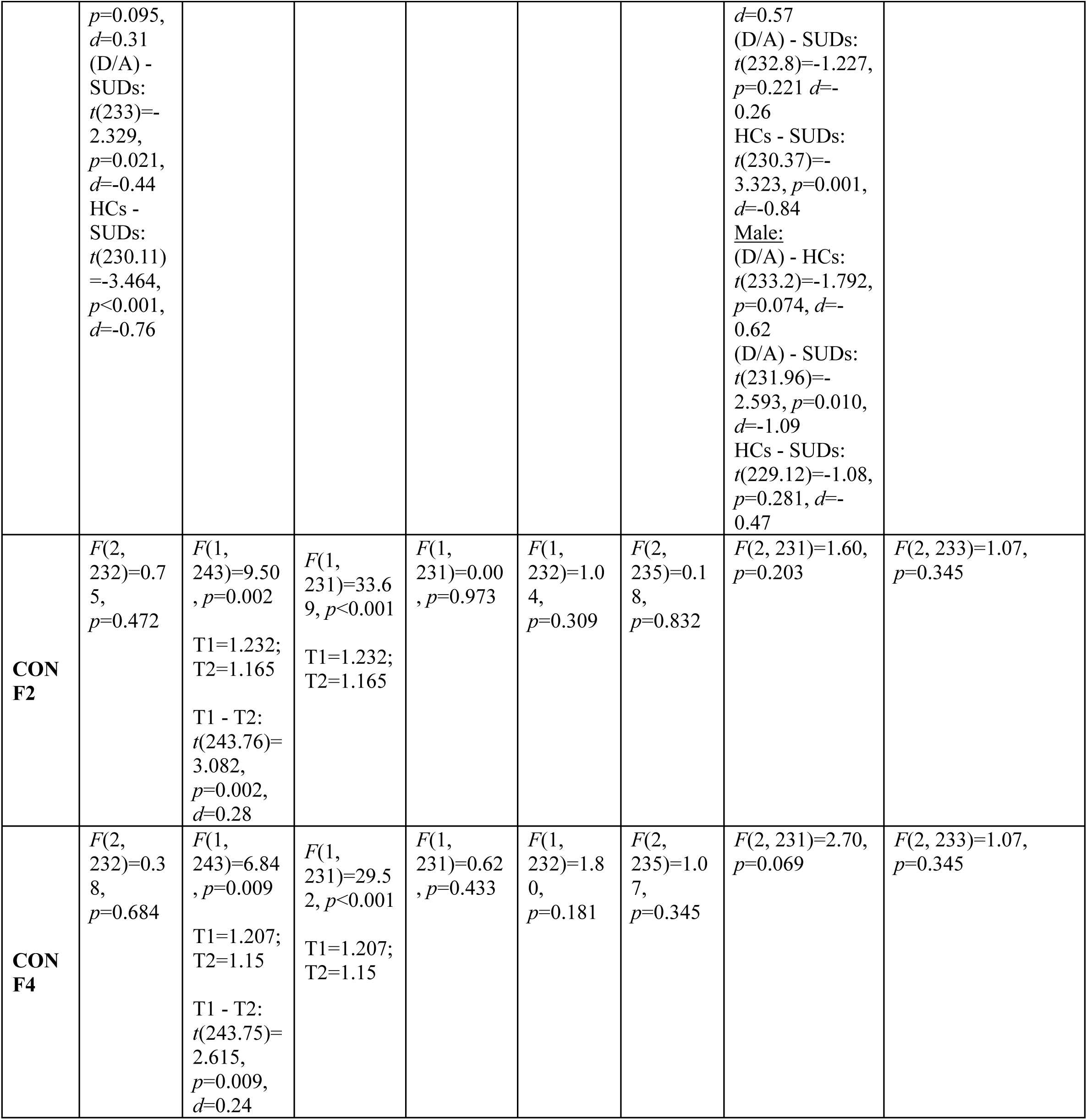
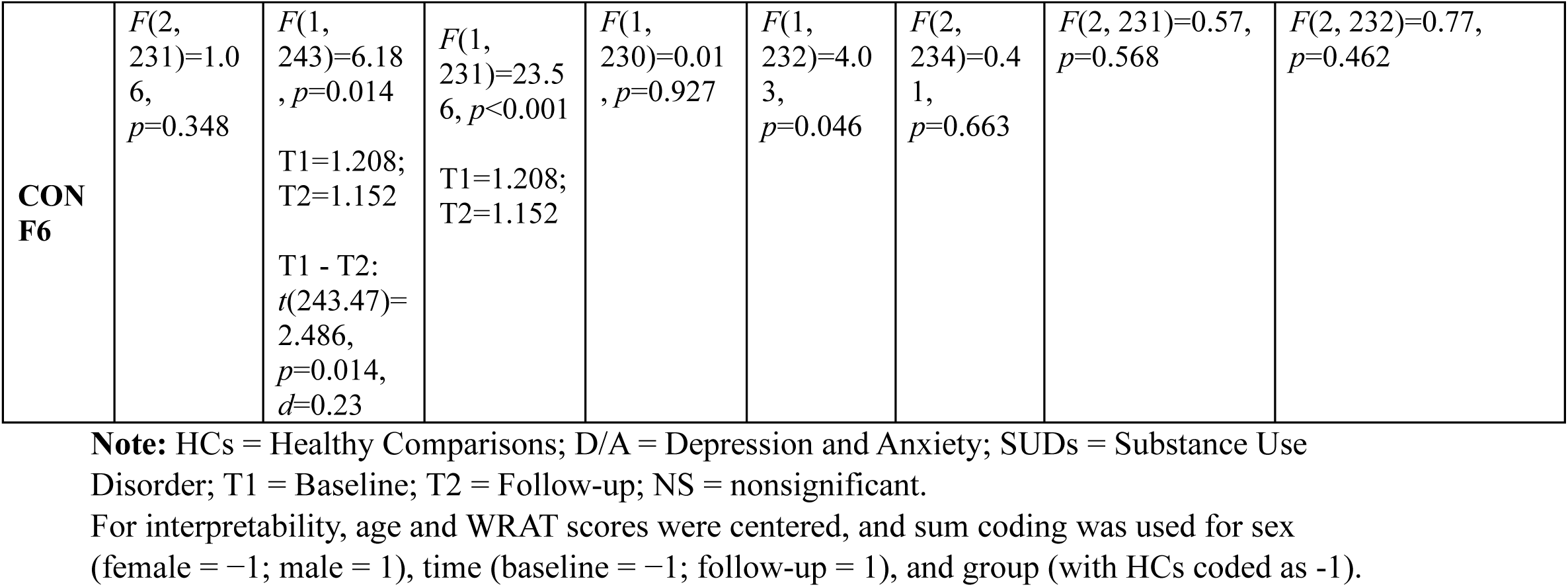
Results of Linear mixed effects models predicting response time (RTs) in participants who returned for follow-up visit, when accounting for effects of group, time, age and sex.

**Supplementary Table 16.** Results of Linear mixed effects models predicting average chosen runway positions (RPs) in participants who returned for follow-up visit, when accounting for effects of group, time, and their interaction.

| Predictor* | Group | Time | Group x Time |
| --- | --- | --- | --- |
| **All Trials** | $F(2, 284)=4.47, p=0.012$<br>D/A=7.321; HCs=6.874; SUDs=7.656<br>(D/A) - HCs: $t(284)=1.88, p=0.061, d=0.46$<br>(D/A) - SUDs: $t(284)=-1.498, p=0.135, d=-0.34$<br>HCs - SUDs: $t(284)=-2.99, p=0.003, d=-0.8$ | $F(1, 284)=0.83, p=0.362$ | $F(2, 283)=0.48, p=0.617$ |
| **Approach** | $F(2, 283)=3.08, p=0.048$<br>D/A=8.348; HCs=8.613; SUDs=8.146<br>(D/A) - HCs: $t(284)=-1.546, p=0.123, d=-0.3$<br>(D/A) - SUDs: $t(284)=1.255, p=0.210, d=0.23$<br>HCs - SUDs: $t(284)=2.479, p=0.014, d=0.52$ | $F(1, 284)=3.99, p=0.047$<br>T1=8.291;<br>T2=8.447<br>T1 - T2:<br>$t(284)=-1.999, p=0.047, d=-0.17$ | $F(2, 284)=2.03, p=0.133$ |
| **Avoid** | $F(2, 284)=20.14, p<0.001$<br>D/A=8.056; HCs=8.29; SUDs=6.847<br>(D/A) - HCs: $t(284)=-1.002, p=0.317, d=-0.21$<br>(D/A) - SUDs: $t(284)=5.49, p<0.001,$ | $F(1, 284)=14.22, p<0.001$<br>T1=7.545;<br>T2=7.917 | $F(2, 283)=5.04, p=0.007$<br><u>Baseline EMM:</u><br>D/A=7.906; HCs=8.287; SUDs=6.441;<br><u>Follow-up EMM:</u><br>D/A=8.206; HCs=8.292; SUDs=7.253 |
| | $d=1.06$<br>HCs - SUDs: $t(284)=5.61, p<0.001, d=1.27$ | T1 - T2:<br>$t(284)=-3.771, p<0.001, d=-0.33$ | <u>Baseline:</u><br>(D/A) - HCs: $t(422.13)=-1.456, p=0.146, d=-0.34$<br>(D/A) - SUDs: $t(422.13)=5.927, p<0.001, d=1.29$<br>HCs - SUDs: $t(422.13)=6.397, p<0.001, d=1.62$<br><u>Follow-up:</u><br>(D/A) - HCs: $t(422.13)=-0.328, p=0.743, d=-0.08$<br>(D/A) - SUDs: $t(422.13)=3.857, p<0.001, d=0.84$<br>HCs - SUDs: $t(422.13)=3.6, p<0.001, d=0.91$ |
| **CONF2** | $F(2, 283)=11.57, p<0.001$<br>D/A=6.462; HCs=5.492; SUDs=7.533<br>(D/A) - HCs: $t(284)=2.506, p=0.013, d=0.59$<br>(D/A) - SUDs: $t(284)=-2.933, p=0.004, d=-0.65$<br>HCs - SUDs: $t(284)=-4.788, p<0.001, d=-1.25$ | $F(1, 283)=0.64, p=0.426$ | $F(2, 284)=0.41, p=0.667$ |
| **CONF4** | $F(2, 284)=11.29, p<0.001$<br>D/A=6.783; HCs=5.825; SUDs=7.796<br>(D/A) - HCs: $t(284)=2.533, p=0.012, d=0.6$<br>(D/A) - SUDs: $t(284)=-2.844, p=0.005, d=-0.64$<br>HCs - SUDs: $t(284)=-4.736, p<0.001, d=-1.24$ | $F(1, 283)=0.78, p=0.379$ | $F(2, 283)=0.18, p=0.836$ |
| **CONF6** | $F(2, 284)=9.54, p<0.001$<br>D/A=6.954; HCs=6.148; SUDs=7.96<br>(D/A) - HCs: $t(284)=2.118, p=0.035, d=0.52$<br>(D/A) - SUDs: $t(284)=-2.805, p=0.005, d=-0.65$<br>HCs - SUDs: $t(284)=-4.325, p<0.001, d=-1.18$ | $F(1, 283)=1.00, p=0.318$ | $F(2, 284)=0.05, p=0.954$ |

**Note:** HCs = Healthy Comparisons; D/A = Depression and Anxiety; SUDs = Substance Use Disorder; T1 = Baseline; T2 = Follow-up; NS = nonsignificant.
For interpretability, sum coding was used for time (baseline = -1; follow-up = 1), and group (with HCs coded as -1).

**Supplementary Table 17.** Results of Linear mixed effects models predicting average chosen runway positions (RPs) in participants who returned for follow-up visit, when accounting for effects of group, time, age and sex.

| Predictor* | Group | Time | Age | Sex | Group x Age | Group x Sex |
| --- | --- | --- | --- | --- | --- | --- |
| **All Trials** | $F(2, 278)=2.99, p=0.052$ | $F(1, 295)=2.10, p=0.149$ | $F(1, 279)=4.93, p=0.027$ | $F(1, 278)=0.93, p=0.336$ | $F(2, 287)=0.68, p=0.507$ | $F(2, 278)=1.67, p=0.190$ |
| **Approach** | $F(2, 278)=3.87, p=0.022$<br>D/A=8.335; HCs=8.598; SUDs=8.142<br>(D/A) - HCs: $t(278)=-1.552, p=0.122, d=-0.29$<br>(D/A) - SUDs: $t(278)=1.224, p=0.222, d=0.22$<br>HCs - SUDs: $t(278)=2.457, p=0.015, d=0.51$ | $F(1, 291)=7.25, p=0.007$<br>T1=8.234; T2=8.437<br>T1 - T2: $t(291.59)=-2.693, p=0.007, d=-0.23$ | $F(1, 279)=10.07, p=0.002$<br>T1=8.234; T2=8.437 | $F(1, 278)=0.36, p=0.550$ | $F(2, 283)=0.54, p=0.585$ | $F(2, 278)=1.14, p=0.321$ |
| **Avoid** | $F(2, 278)=12.76, p<0.001$<br>D/A=8.054; HCs=8.3; SUDs=6.842<br>(D/A) - HCs: $t(278)=-1.048, p=0.296, d=-0.21$<br>(D/A) - SUDs: $t(278)=5.514, p<0.001, d=1.05$<br>HCs - SUDs: $t(278)=5.654, p<0.001, d=1.26$ | $F(1, 292)=17.14, p<0.001$<br>T1=7.545; T2=7.946<br>T1 - T2: $t(292.57)=-4.141, p<0.001, d=-0.35$ | $F(1, 279)=3.86, p=0.051$ | $F(1, 278)=0.06, p=0.799$ | $F(2, 284)=0.42, p=0.658$ | $F(2, 278)=0.79, p=0.457$ |
| **CONF2** | $F(2, 278)=7.46, p<0.001$<br>D/A=6.453; HCs=5.396; SUDs=7.526<br>(D/A) - HCs: $t(278)=2.712, p=0.007, d=0.65$<br>(D/A) - SUDs: $t(278)=-2.941, p=0.004, d=-0.66$<br>HCs - SUDs: $t(278)=-4.978, p<0.001, d=-1.31$ | $F(1, 295)=1.50, p=0.222$ | $F(1, 279)=1.40, p=0.237$ | $F(1, 278)=0.73, p=0.393$ | $F(2, 287)=0.87, p=0.419$ | $F(2, 278)=1.78, p=0.170$ |
| **CONF4** | $F(2, 278)=7.98, p<0.001$<br>D/A=6.774; HCs=5.738;<br>SUDs=7.789<br>(D/A) - HCs:<br>$t(278)=2.718, p=0.007, d=0.65$<br>(D/A) - SUDs: $t(278)=-2.844, p = 0.005 d=-0.64$<br>HCs - SUDs: $t(278)=-4.901, p<0.001, d=-1.29$ | $F(1, 295)=0.31, p=0.580$ | $F(1, 279)=2.08, p=0.151$ | $F(1, 278)=0.93, p=0.335$ | $F(2, 287)=0.31, p=0.736$ | $F(2, 278)=1.46, p=0.235$ |
| **CONF6** | $F(2, 278)=7.18, p<0.001$<br>D/A=6.952; HCs=6.063;<br>SUDs=7.95<br>(D/A) - HCs:<br>$t(278)=2.317, p=0.021, d=0.58$<br>(D/A) - SUDs: $t(278)=-2.782, p=0.006, d=-0.65$<br>HCs - SUDs: $t(278)=-4.483, p<0.001, d=-1.23$ | $F(1, 295)=0.66, p=0.416$ | $F(1, 279)=2.81, p=0.095$ | $F(1, 278)=1.65, p=0.200$ | $F(2, 288)=0.45, p=0.639$ | $F(2, 278)=1.10, p=0.336$ |

**Note:** HCs = Healthy Comparisons; D/A = Depression and Anxiety; SUDs = Substance Use Disorder; T1 = Baseline; T2 = Follow-up; NS = nonsignificant.
For interpretability, Age was centered, and sum coding was used for sex (female = -1; male = 1), time (baseline = -1; follow-up = 1), and group (with HCs coded as -1).

**Supplementary Table 18.** Results of Linear mixed effects models predicting average chosen runway positions (RPs) in participants who returned for follow-up visit, when accounting for effects of group, time, age, sex and WRAT scores.

| **Predictor \*** | **Group** | **Time** | **Age** | **Sex** | **WRAT** | **Group x Age** | **Group x Sex** | **Group x WRAT** |
| --- | --- | --- | --- | --- | --- | --- | --- | --- |
| **All Trials** | $F(2, 232)=4.52, p=0.012$<br>D/A=7.44;<br>HCs=6.585;<br>SUDs=7.826<br>(D/A) - HCs:<br>$t(232.47)=3.26, p=0.001, d=0.94$<br>(D/A) - SUDs:<br>$t(233.23)=-1.455,$ | $F(1, 246)=2.81, p=0.095$ | $F(1, 233)=8.90, p=0.003$ | $F(1, 232)=3.01, p=0.084$ | $F(1, 233)=0.17, p=0.680$ | $F(2, 241)=1.62, p=0.200$ | $F(2, 232)=1.56, p=0.212$ | $F(2, 233)=1.28, p=0.279$ |
| | $p=0.147$, $d=-0.43$<br>HCs - SUDs:<br>$t(231.53)=-4.057$,<br>$p<0.001$, $d=-1.37$ | | | | | | | |
| **Approach** | $F(2, 232)=1.89$,<br>$p=0.153$ | $F(1, 244)=6.94$,<br>$p=0.009$<br>T1=8.286;<br>T2=8.491<br>T1 - T2:<br>$t(244.04)=-2.634$,<br>$p=0.009$,<br>$d=-0.24$ | $F(1, 232)=9.95$,<br>$p=0.002$<br>T1=8.286;<br>T2=8.491 | $F(1, 231)=0.00$,<br>$p=0.970$ | $F(1, 232)=14.30$,<br>$p<0.001$ | $F(2, 236)=1.04$,<br>$p=0.355$ | $F(2, 231)=1.69$,<br>$p=0.187$ | $F(2, 233)=1.12$,<br>$p=0.329$ |
| **Avoid** | $F(2, 232)=5.68$,<br>$p=0.004$<br>D/A=7.863;<br>HCs=8.161;<br>SUDs=6.813<br>(D/A) - HCs:<br>$t(232.05)=-1.157$,<br>$p=0.249$, $d=-0.25$<br>(D/A) - SUDs:<br>$t(233.15)=4.035$,<br>$p<0.001$, $d=0.9$<br>HCs - SUDs:<br>$t(230.69)=4.495$,<br>$p<0.001$, $d=1.15$ | $F(1, 244)=14.50$,<br>$p<0.001$<br>T1 =7.443;<br>T2= 7.854<br>T1 - T2:<br>$t(244.2)=-3.807$,<br>$p<0.001$,<br>$d=-0.35$ | $F(1, 232)=3.80$,<br>$p=0.053$ | $F(1, 231)=0.21$,<br>$p=0.645$ | $F(1, 232)=18.71$,<br>$p<0.001$ | $F(2, 236)=0.85$,<br>$p=0.430$ | $F(2, 231)=0.38$,<br>$p=0.686$ | $F(2, 233)=5.58$,<br>$p=0.004$<br>D/A = 0.131; HCs = 0.098; SUDs = 0.004<br>(D/A) - HCs:<br>$t(234)=0.712$, $p = 0.757$<br>(D/A) - SUDs:<br>$t(235)=3.24$, $p = 0.004$, $d=0.11$<br>HCs - SUDs:<br>$t(230)=2.06$,<br>$p=0.101$ |
| **CONF2** | $F(2, 232)=7.41$,<br>$p<0.001$<br>D/A=6.787;<br>HCs=5.112;<br>SUDs=7.792<br>(D/A) - HCs:<br>$t(232.44)=3.905$,<br>$p<0.001$, $d=1.1$<br>(D/A) - SUDs:<br>$t(233.23)=-2.318$,<br>$p=0.021$, $d=-0.66$<br>HCs - SUDs: | $F(1, 246)=1.58$,<br>$p=0.210$ | $F(1, 233)=3.70$,<br>$p=0.056$ | $F(1, 231)=3.07$,<br>$p=0.081$ | $F(1, 233)=1.93$,<br>$p=0.167$ | $F(2, 240)=1.59$,<br>$p=0.205$ | $F(2, 232)=1.80$,<br>$p=0.167$ | $F(2, 233)=2.72$,<br>$p=0.068$ |
| | $t(231.47)=-5.358$,<br>$p<0.001$, $d=-1.77$ | | | | | | | |
| **CON F4** | $F(2, 232)=7.62$,<br>$p<0.001$<br>D/A=7.054;<br>HCs=5.444;<br>SUDs=8.003<br>(D/A) - HCs:<br>$t(232.44)=3.825$,<br>$p<0.001$, $d=1.08$<br>(D/A) - SUDs:<br>$t(233.23)=-2.231$,<br>$p=0.027$, $d=-0.64$<br>HCs - SUDs:<br>$t(231.48)=-5.214$,<br>$p<0.001$, $d=-1.72$ | $F(1, 246)=0.08$,<br>$p=0.773$ | $F(1, 233)=5.35$,<br>$p=0.022$ | $F(1, 231)=2.44$,<br>$p=0.120$ | $F(1, 233)=0.66$,<br>$p=0.419$ | $F(2, 240)=1.13$,<br>$p=0.324$ | $F(2, 232)=1.31$,<br>$p=0.272$ | $F(2, 233)=2.12$,<br>$p=0.123$ |
| **CON F6** | $F(2, 232)=6.55$,<br>$p=0.002$<br>D/A=7.202;<br>HCs=5.747;<br>SUDs=8.129<br>(D/A) - HCs:<br>$t(232.5)=3.413$,<br>$p<0.001$, $d=1.02$<br>(D/A) - SUDs:<br>$t(233.23)=-2.152$,<br>$p=0.032$, $d=-0.65$<br>HCs - SUDs:<br>$t(231.61)=-4.791$,<br>$p<0.001$, $d=-1.67$ | $F(1, 247)=0.15$,<br>$p=0.696$ | $F(1, 233)=6.13$,<br>$p=0.014$ | $F(1, 232)=3.03$,<br>$p=0.083$ | $F(1, 233)=0.61$,<br>$p=0.435$ | $F(2, 242)=0.93$,<br>$p=0.394$ | $F(2, 232)=1.14$,<br>$p=0.322$ | $F(2, 233)=1.99$,<br>$p=0.139$ |

**Note:** HCs = Healthy Comparisons; D/A = Depression and Anxiety; SUDs = Substance Use Disorder; T1 = Baseline; T2 = Follow-up; NS = nonsignificant.
For interpretability, age and WRAT scores were centered, and sum coding was used for sex (female = -1; male = 1), time (baseline = -1; follow-up = 1), and group (with HCs coded as -1).

**Supplementary Table 19.** Results of Linear mixed effects models predicting choice variability in participants who returned for follow-up visit, when accounting for effects of group, time, and their interaction.

| Predictor* | Group | Time | Group x Time |
| --- | --- | --- | --- |
| All Trials | $F(2, 283)=2.28, p=0.104$ | $F(1, 284)=8.28, p=0.004$<br>T1=2.081; T2=1.874<br>T1 - T2: $t(284)=2.877, p=0.004, d=0.25$ | $F(2, 284)=2.25, p=0.107$ |
| Approach | $F(2, 284)=5.89, p=0.003$<br>D/A=0.701; HCs=0.501; SUDs=1.006<br>(D/A) - HCs: $t(284)=1.464, p=0.144, d=0.23$<br>(D/A) - SUDs: $t(284)=-2.371, p=0.018, d=-0.35$<br>HCs - SUDs: $t(284)=-3.361, p<0.001, d=-0.57$ | $F(1, 284)=4.62, p=0.033$<br>T1=0.818; T2=0.654<br>T1 - T2: $t(284)=2.149, p=0.033, d=0.19$ | $F(2, 284)=2.49, p=0.084$ |
| Avoid | $F(2, 284)=18.52, p<0.001,$<br>D/A=0.859; HCs=0.731; SUDs=1.605<br>(D/A) - HCs: $t(284)=0.864, p=0.388, d=0.15$<br>(D/A) - SUDs: $t(284)=-5.313, p<0.001, d=-0.88$<br>HCs - SUDs: $t(284)=-5.334, p<0.001, d=-1.03$ | $F(1, 284)=21.24, p<0.001$<br>T1=1.235; T2=0.895<br>T1 - T2: $t(284)=4.609, p<0.001, d=0.4$ | $F(2, 284)=5.03, p=0.007$<br><u>Baseline EMM:</u><br>D/A=1.018; HCs=0.753; SUDs=1.935;<br><u>Follow-up EMM:</u><br>D/A=0.701; HCs=0.708; SUDs=1.276<br><u>Baseline:</u><br>(D/A) - HCs: $t(463.59)=1.524, p=0.128, d=0.31$<br>(D/A) - SUDs: $t(463.59)=-5.611, p<0.001, d=-1.08$<br>HCs - SUDs: $t(463.59)=-6.188, p<0.001, d=-1.39$<br><u>Follow-up:</u><br>(D/A) - HCs: $t(463.59)=-0.04, p=0.968, d=-0.01$<br>(D/A) - SUDs: $t(463.59)=-3.513, p<0.001, d=-0.68$<br>HCs - SUDs: $t(463.59)=-2.971, p=0.003, d=-0.67$ |
| CONF2 | $F(2, 284)=0.63, p=0.533$ | $F(1, 283)=16.49, p<0.001$ | $F(2, 283)=1.49, p=0.226$ |
| | | T1=1.211; T2=0.877<br>T1 - T2: $t(284)=4.061$,<br>$p<0.001$, $d=0.35$ | |
| **CONF4** | $F(2, 284)=1.76$, $p=0.173$ | $F(1, 283)=12.76$, $p<0.001$<br>T1=1.097; T2=0.811<br>T1 - T2: $t(284)=3.572$,<br>$p<0.001$, $d=0.31$ | $F(2, 284)=0.56$, $p=0.570$ |
| **CONF6** | $F(2, 284)=0.60$, $p=0.551$ | $F(1, 284)=5.04$, $p=0.026$<br>T1=0.959; T2=0.776<br>T1 - T2: $t(284)=2.245$,<br>$p=0.026$, $d=0.19$ | $F(2, 283)=1.19$, $p=0.307$ |

**Note:** HCs = Healthy Comparisons; D/A = Depression and Anxiety; SUDs = Substance Use Disorder; T1 = Baseline; T2 = Follow-up; NS = nonsignificant.
For interpretability, sum coding was used for time (baseline = -1; follow-up = 1), and group (with HCs coded as -1).

**Supplementary Table 20.**
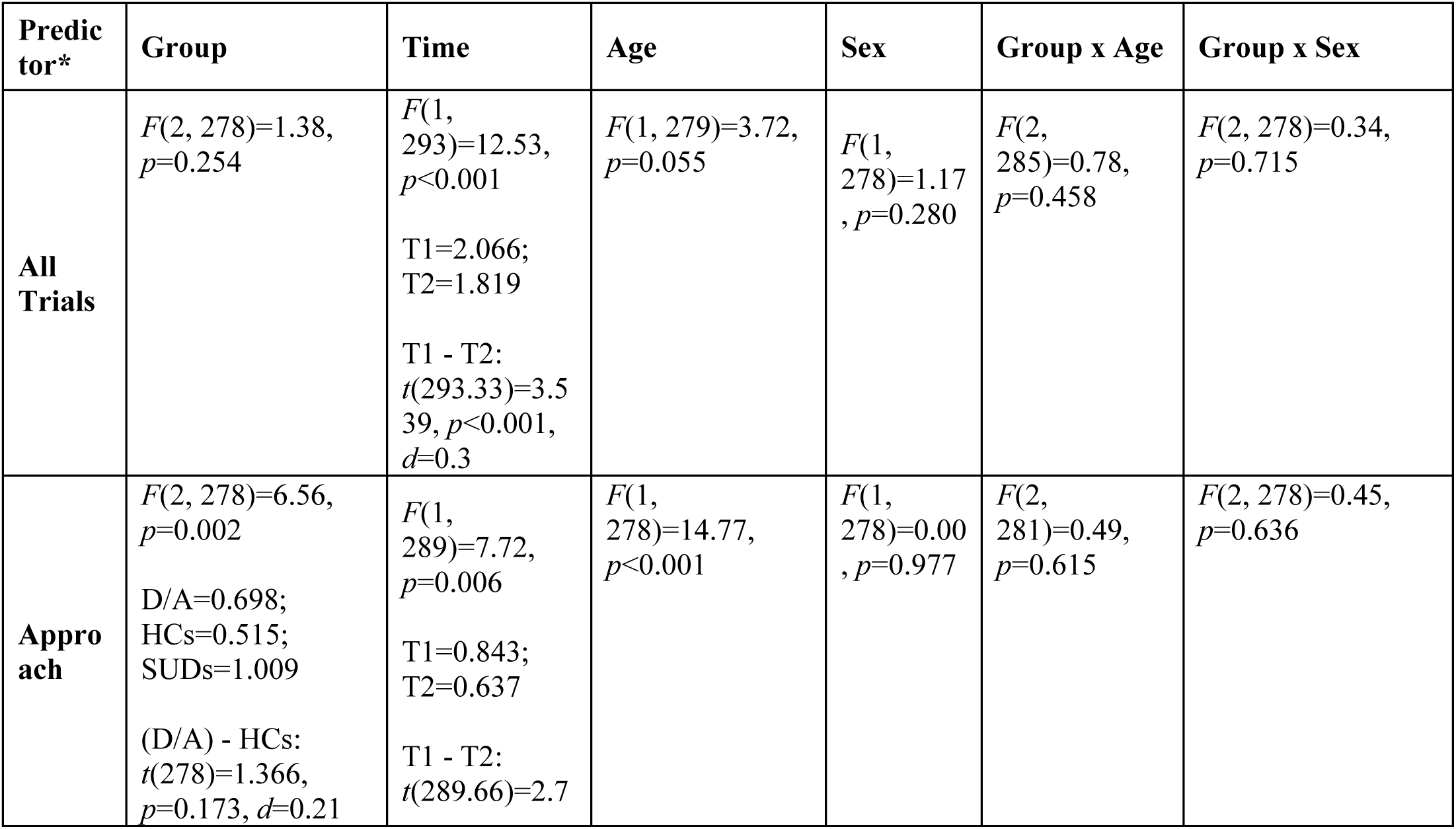
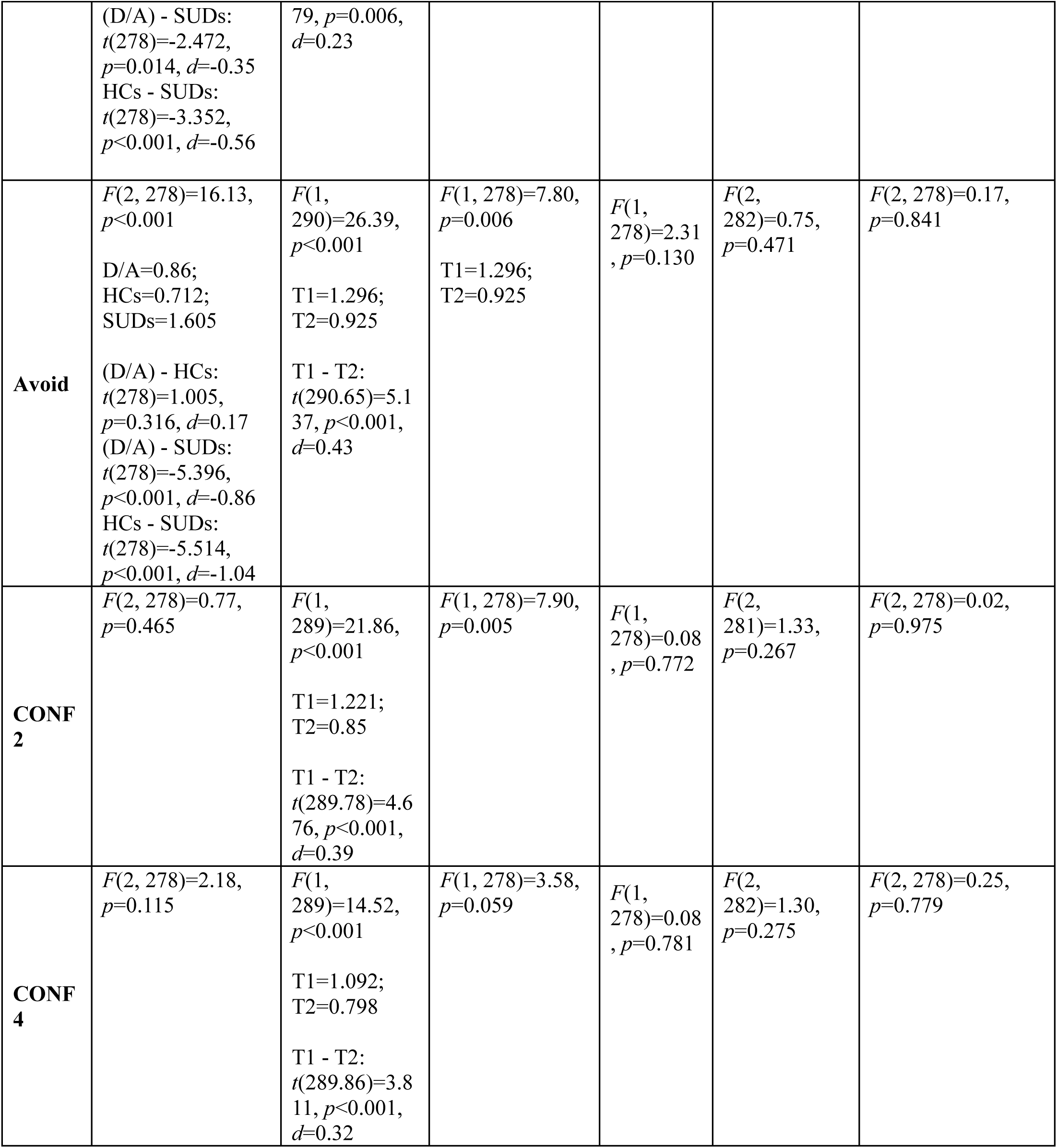
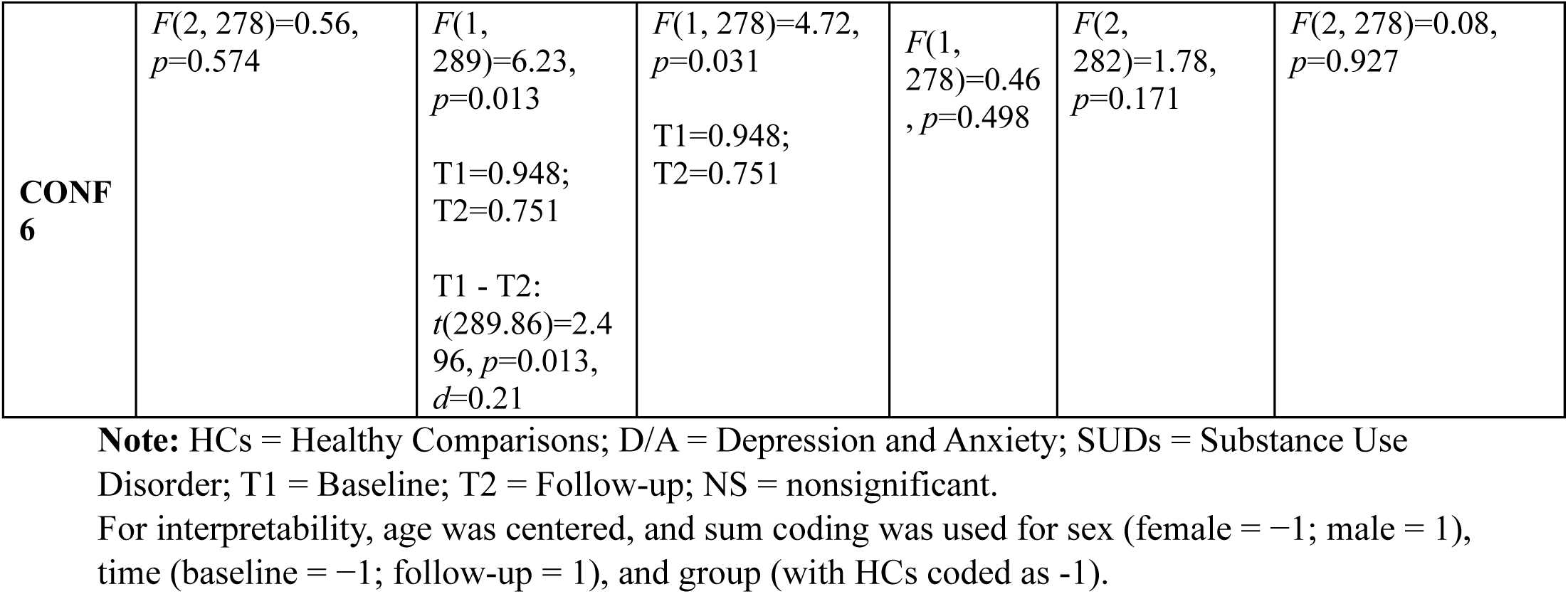
Results of Linear mixed effects models predicting choice variability in participants who returned for follow-up visit, when accounting for effects of group, time, age, and sex.

**Supplementary Table 21.** Results of Linear mixed effects models predicting choice variability in participants who returned for follow-up visit, when accounting for effects of group, time, age, sex and WRAT scores.

| Predictor* | Group | Time | Age | Sex | WRAT | Group x Age | Group x Sex | Group x WRAT |
| --- | --- | --- | --- | --- | --- | --- | --- | --- |
| **All Trials** | $F(2, 232)=3.19$,<br>$p=0.043$<br>D/A=1.773;<br>HCs=2.384;<br>SUDs=1.799<br>(D/A) - HCs:<br>$t(232.24)=-3.064$,<br>$p=0.002$, $d=-0.75$<br>(D/A) - SUDs:<br>$t(233.2)=-0.127$,<br>$p=0.899$, $d=-0.03$<br>HCs - SUDs:<br>$t(231.07)=2.518$,<br>$p=0.012$, $d=0.72$ | $F(1, 245)=12.73$,<br>$p<0.001$<br>T1=2.006;<br>T2=1.739<br>T1 - T2:<br>$t(245.03)=3.567$,<br>$p<0.001$,<br>$d=0.33$ | $F(1, 232)=6.59$,<br>$p=0.011$ | $F(1, 231)=3.78$,<br>$p=0.053$ | $F(1, 233)=0.58$,<br>$p=0.445$ | $F(2, 238)=2.01$,<br>$p=0.137$ | $F(2, 232)=0.11$,<br>$p=0.900$ | $F(2, 233)=0.63$,<br>$p=0.536$ |
| **Approach** | $F(2, 231)=0.82$,<br>$p=0.443$ | $F(1, 243)=4.37$,<br>$p=0.038$<br>T1=0.767;<br>T2=0.602<br>T1 - T2:<br>$t(243.37)=2.091$, | $F(1, 231)=13.11$,<br>$p<0.001$<br>T1=0.767;<br>T2=0.602 | $F(1, 230)=0.80$,<br>$p=0.373$ | $F(1, 232)=16.28$, $p<0.001$ | $F(2, 233)=0.56$,<br>$p=0.574$ | $F(2, 231)=0.22$,<br>$p=0.802$ | $F(2, 232)=1.65$,<br>$p=0.194$ |
| | | $p=0.038$,<br>$d=0.19$ | | | | | | |
| **Avoid** | $F(2, 231)=5.24$,<br>$p=0.006$<br>$D/A=0.962$;<br>$HCs=0.753$;<br>$SUDs=1.475$<br>(D/A) - HCs:<br>$t(231.64)=1.306$,<br>$p=0.193$, $d=0.24$<br>(D/A) - SUDs:<br>$t(232.96)=-3.155$,<br>$p=0.002$, $d=-0.58$<br>HCs - SUDs: $t(230)=-3.862$, $p<0.001$, $d=-0.82$ | $F(1, 243)=21.42$,<br>$p<0.001$<br>$T1=1.287$;<br>$T2=0.912$<br>$T1 - T2$:<br>$t(243.48)=4.629$,<br>$p<0.001$,<br>$d=0.42$ | $F(1, 231)=5.95$,<br>$p=.015$<br>$T1=1.287$;<br>$T2=0.912$ | $F(1, 230)=0.69$,<br>$p=0.406$ | $F(1, 232)=16.10$, $p<0.001$ | $F(2, 234)=1.47$,<br>$p=0.232$ | $F(2, 231)=0.43$,<br>$p=0.651$ | $F(2, 232)=1.51$,<br>$p=0.223$ |
| **CON F2** | $F(2, 231)=0.20$,<br>$p=0.817$ | $F(1, 243)=13.15$,<br>$p<0.001$<br>$T1=1.201$;<br>$T2=0.88$<br>$T1 - T2$:<br>$t(243.34)=3.626$,<br>$p<0.001$,<br>$d=0.33$ | $F(1, 230)=6.03$,<br>$p=0.015$ | $F(1, 230)=0.27$,<br>$p=0.607$ | $F(1, 232)=10.09$, $p=0.002$ | $F(2, 233)=0.52$,<br>$p=0.595$ | $F(2, 230)=0.03$,<br>$p=0.971$ | $F(2, 232)=1.78$,<br>$p=0.171$ |
| **CON F4** | $F(2, 231)=0.85$,<br>$p=0.428$ | $F(1, 243)=13.98$,<br>$p<0.001$<br>$T1=1.126$;<br>$T2=0.811$<br>$T1 - T2$:<br>$t(243.41)=3.738$,<br>$p<0.001$,<br>$d=0.34$ | $F(1, 231)=2.68$,<br>$p=0.103$ | $F(1, 230)=0.04$,<br>$p=0.833$ | $F(1, 232)=8.05$,<br>$p=0.005$ | $F(2, 234)=1.11$,<br>$p=0.331$ | $F(2, 231)=0.41$,<br>$p=0.662$ | $F(2, 232)=1.63$,<br>$p=0.199$ |
| **CON**<br>**F6** | $F(2, 231)=0.08$,<br>$p=0.921$ | $F(1, 243)=4.58$,<br>$p=0.033$<br>T1=0.967;<br>T2=0.781<br>T1 - T2:<br>$t(243.36)=2.139$,<br>$p=0.033$,<br>$d=0.2$ | $F(1, 230)=3.54$,<br>$p=0.061$ | $F(1, 230)=0.46$,<br>$p=0.497$ | $F(1, 232)=10.89$, $p=0.001$ | $F(2, 233)=0.96$,<br>$p=0.385$ | $F(2, 231)=0.04$,<br>$p=0.963$ | $F(2, 232)=1.37$,<br>$p=0.255$ |

**Note:** HCs = Healthy Comparisons; D/A = Depression and Anxiety; SUDs = Substance Use Disorder; T1 = Baseline; T2 = Follow-up; NS = nonsignificant.
For interpretability, age and WRAT scores were centered, and sum coding was used for sex (female = -1; male = 1), time (baseline = -1; follow-up = 1), and group (with HCs coded as -1).

**Supplementary Table 22.** Results of Linear mixed effects models predicting self-reported items in participants who returned for follow-up visit, when accounting for effects of group, time and their interaction.

| Predictor* | Group | Time | Group x Time |
| --- | --- | --- | --- |
| **Neg. Anxious** | $F(2, 284)=2.69$, $p=0.070$ | $F(1, 284)=0.00$, $p=0.980$ | $F(2, 284)=0.72$, $p=0.489$ |
| **Diff. Decide** | $F(2, 284)=0.66$, $p=0.519$ | $F(1, 283)=3.58$, $p=0.060$ | $F(2, 283)=0.17$, $p=0.846$ |
| **App. Reward** | $F(2, 283)=8.54$, $p<0.001$<br>D/A=4.815; HCs=4.239; SUDs=5.627<br>(D/A) - HCs: $t(284)=1.858$, $p=0.064$, $d=0.37$<br>(D/A) - SUDs: $t(284)=-2.78$, $p=0.006$, $d=-0.53$<br>HCs - SUDs: $t(284)=-4.068$, $p<0.001$, $d=-0.9$ | $F(1, 283)=1.19$, $p=0.276$ | $F(2, 284)=0.31$, $p=0.733$ |
| **Avoid Neg.** | $F(2, 284)=4.98$, $p=0.007$<br>D/A=3.044; HCs=3.333; SUDs=2.386<br>(D/A) - HCs: $t(284)=-1.005$, $p=0.316$, $d=-0.19$<br>(D/A) - SUDs: $t(284)=2.432$, $p=0.016$, $d=0.42$<br>HCs - SUDs: $t(284)=2.995$, $p=0.003$, $d=0.61$ | $F(1, 284)=0.04$, $p=0.845$ | $F(2, 283)=0.67$, $p=0.513$ |

**Note:** HCs = Healthy Comparisons; D/A = Depression and Anxiety; SUDs = Substance Use Disorder; T1 = Baseline; T2 = Follow-up; NS = nonsignificant.
For interpretability, sum coding was used time (baseline = -1; follow-up = 1), and group (with HCs coded as -1).

**Supplementary Table 23.**
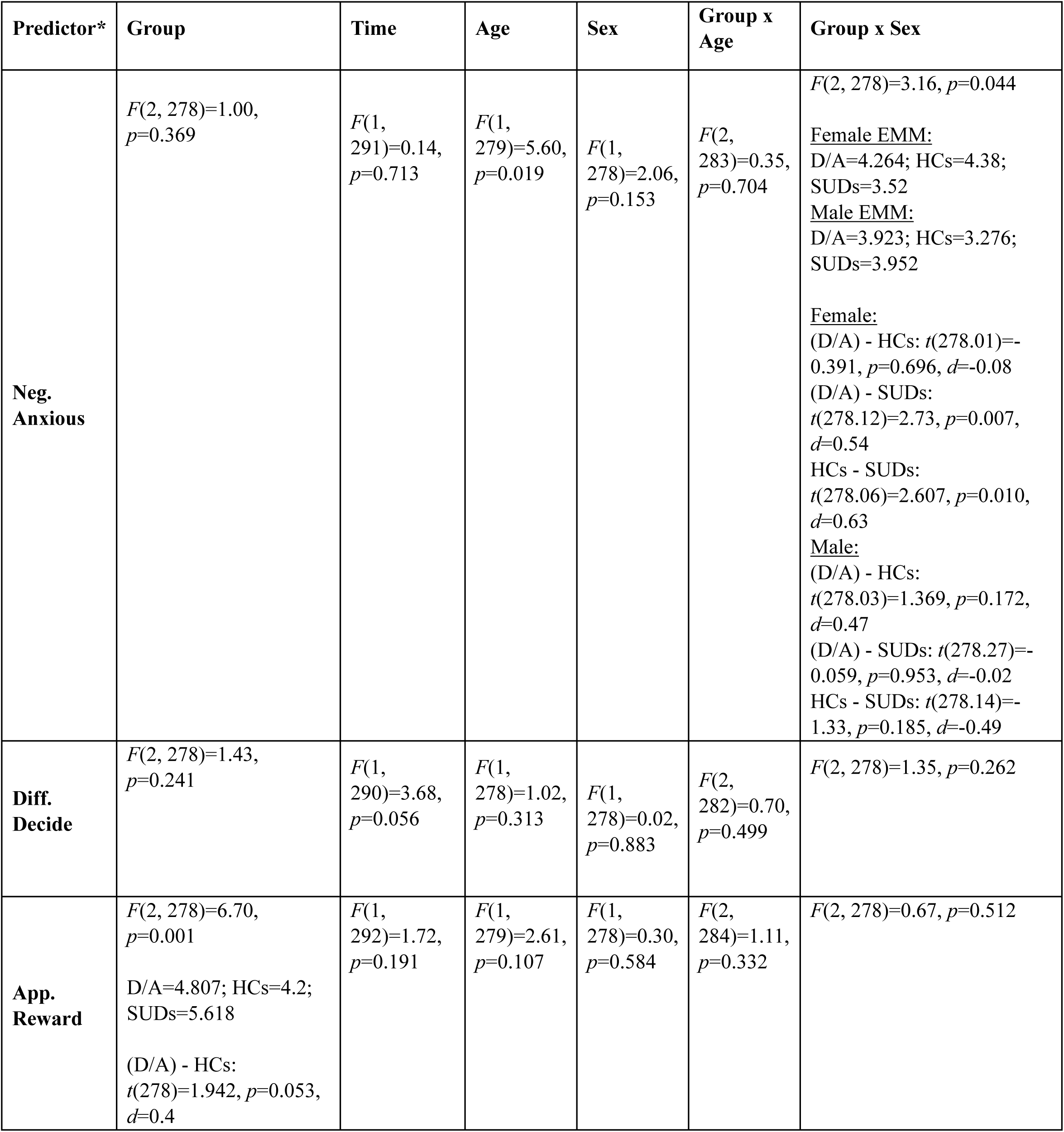
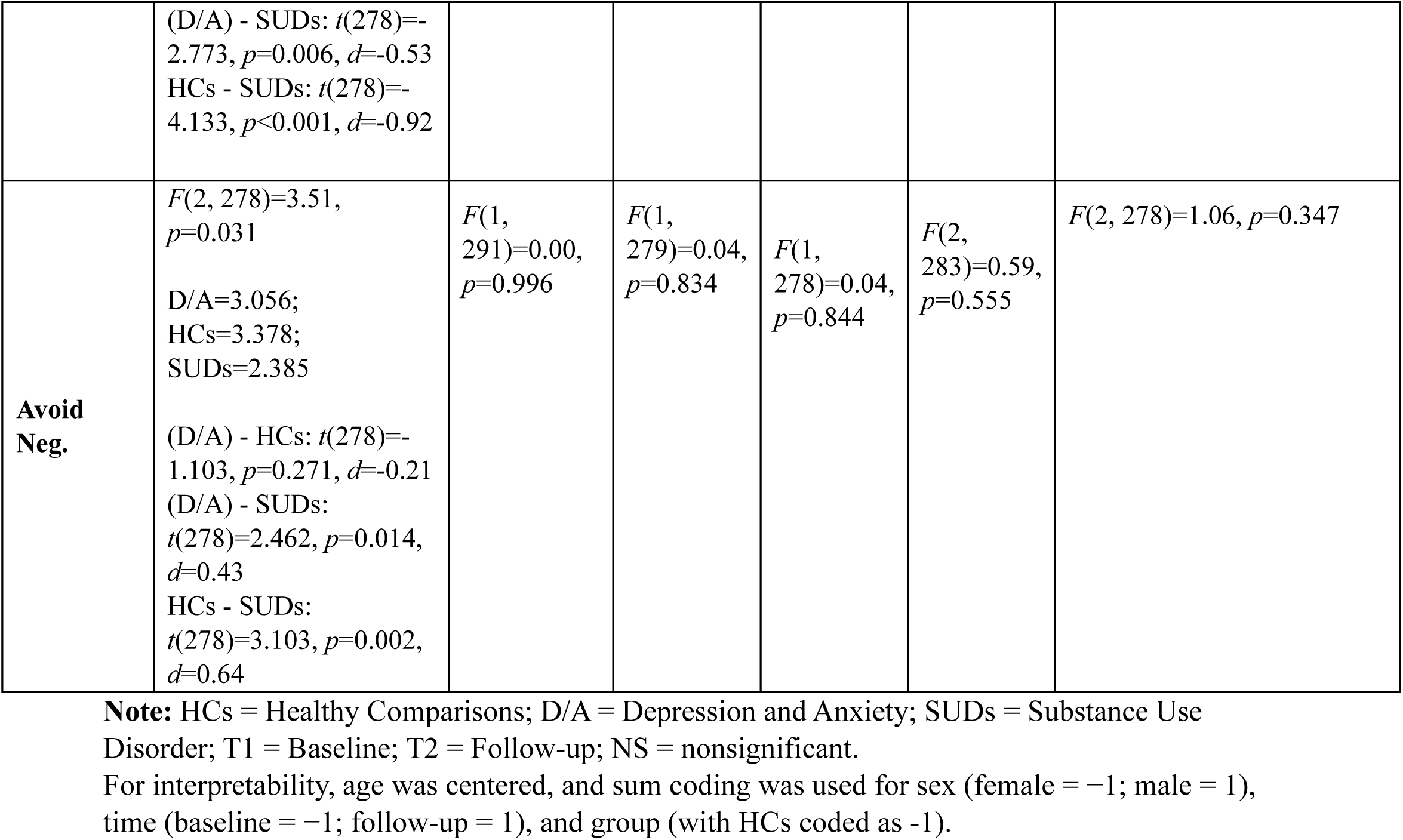
Results of Linear mixed effects models predicting self-reported items in participants who returned for follow-up visit, when accounting for effects of group, time age and sex.

**Supplementary Table 24.** Results of Linear mixed effects models predicting self-reported items in participants who returned for follow-up visit, when accounting for effects of group, time age, sex and WRAT scores.

| Predictor* | Group | Time | Age | Sex | WRAT | Group x Age | Group x Sex | Group x WRAT |
| --- | --- | --- | --- | --- | --- | --- | --- | --- |
| **Neg. Anxious** | $F(2, 232)=2.14, p=0.120$ | $F(1, 244)=0.10, p=0.757$ | $F(1, 232)=2.71, p=0.101$ | $F(1, 231)=2.09, p=0.150$ | $F(1, 232)=0.07, p=0.797$ | $F(2, 236)=0.86, p=0.424$ | $F(2, 231)=2.08, p=0.127$ | $F(2, 233)=3.37, p=0.036$<br>D/A=4.17;<br>HCs=4.108;<br>SUDs=3.193<br>(D/A) - HCs: $t(232)=0.217, p=0.828, d=0.05$<br>(D/A) - SUDs: $t(233.13)=3.345, p<0.001, d=0.72$ |
| | | | | | | | | HCs - SUDs:<br>$t(230.59)=2.718$,<br>$p=0.007$, $d=0.68$ |
| **Diff. Decide** | $F(2, 232)=0.48$,<br>$p=0.620$ | $F(1, 243)=1.33$,<br>$p=0.251$ | $F(1, 231)=0.16$,<br>$p=0.692$ | $F(1, 231)=0.07$,<br>$p=0.791$ | $F(1, 232)=1.19$,<br>$p=0.276$ | $F(2, 235)=0.45$,<br>$p=0.640$ | $F(2, 231)=0.81$,<br>$p=0.447$ | $F(2, 233)=0.14$,<br>$p=0.872$ |
| **App. Reward** | $F(2, 232)=5.64$,<br>$p=0.004$<br>D/A=4.926;<br>HCs=3.966;<br>SUDs=5.746<br>(D/A) - HCs:<br>$t(232.23)=2.681$,<br>$p=0.008$, $d=0.65$<br>(D/A) - SUDs:<br>$t(233.2)=-2.261$,<br>$p=0.025$, $d=-0.56$<br>HCs - SUDs:<br>$t(231.04)=-4.261$,<br>$p<0.001$, $d=-1.21$ | $F(1, 244)=1.13$,<br>$p=0.289$ | $F(1, 232)=5.09$,<br>$p=0.025$ | $F(1, 231)=1.08$,<br>$p=0.300$ | $F(1, 232)=0.04$,<br>$p=0.851$ | $F(2, 238)=1.63$,<br>$p=0.198$ | $F(2, 231)=0.50$,<br>$p=0.605$ | $F(2, 233)=0.47$,<br>$p=0.626$ |
| **Avoid Neg.** | $F(2, 232)=4.43$,<br>$p=0.013$<br>D/A=2.951;<br>HCs=3.565;<br>SUDs=2.159<br>(D/A) - HCs:<br>$t(231.92)=-1.898$,<br>$p=0.059$, $d=-0.39$<br>(D/A) - SUDs:<br>$t(233.1)=2.422$,<br>$p=0.016$, $d=0.5$ | $F(1, 243)=0.05$,<br>$p=0.826$ | $F(1, 231)=1.11$,<br>$p=0.294$ | $F(1, 231)=1.26$,<br>$p=0.262$ | $F(1, 232)=0.06$,<br>$p=0.814$ | $F(2, 235)=3.46$,<br>$p=0.033$<br>D/A=2.951;<br>HCs=3.565;<br>SUDs=2.159<br>(D/A) - HCs:<br>$t(231.92)=-1.898$,<br>$p=0.059$, $d=-0.39$<br>(D/A) - SUDs:<br>$t(233.1)=2.422$,<br>$p=0.016$, $d=0.5$ | $F(2, 231)=0.82$,<br>$p=0.440$ | $F(2, 233)=0.28$,<br>$p=0.756$ |
| | HCs - SUDs:<br>$t(230.47)=3.732$,<br>$p<0.001$, $d=0.9$ | | | | | HCs - SUDs:<br>$t(230.47)=3.732$,<br>$p<0.001$, $d=0.9$ | | |

**Note:** HCs = Healthy Comparisons; D/A = Depression and Anxiety; SUDs = Substance Use Disorder; T1 = Baseline; T2 = Follow-up; NS = nonsignificant.
For interpretability, age and WRAT scores were centered, and sum coding was used for sex (female = -1; male = 1), time (baseline = -1; follow-up = 1), and group (with HCs coded as -1).

**Supplementary Table 25.** Summary statistics for model parameters (Mean (SD)) for various sub-diagnosis at baseline and follow-up in the exploratory and the confirmatory samples.

|  | Baseline |  | Follow-up |  | *p* |
| --- | --- | --- | --- | --- | --- |
|  | *Exploratory* | *Confirmatory* | *Exploratory* | *Confirmatory* |  |
| **Alcohol** | 55 | 63 | 27 | 26 |  |
| **DU** | 1.51 (0.90) | 1.29 (1.03) | 0.66 (1.22) | 0.81 (1.01) | 0.001 |
| **EC** | 0.35 (0.59) | 0.44 (0.68) | 0.34 (0.58) | 0.37 (0.78) | 0.88 |
| **Cannabis** | 84 | 74 | 40 | 29 |  |
| **DU** | 1.44 (0.96) | 0.92 (0.97) | 1.10 (1.04) | 0.47 (0.94) | <0.001 |
| **EC** | 0.56 (0.81) | 0.34 (0.71) | 0.61 (0.71) | 0.01 (0.65) | 0.002 |
| **Stimulant** | 121 | 157 | 63 | 72 |  |
| **DU** | 1.42 (0.89) | 1.17 (1.01) | 0.92 (1.06) | 0.73 (1.07) | <0.001 |
| **EC** | 0.48 (0.76) | 0.42 (0.68) | 0.48 (0.71) | 0.24 (0.81) | 0.132 |
| **Opioid** | 64 | 58 | 32 | 30 |  |
| **DU** | 1.51 (0.84) | 1.14 (0.98) | 0.83 (1.09) | 0.68 (1.00) | <0.001 |
| **EC** | 0.44 (0.67) | 0.44 (0.64) | 0.56 (0.73) | 0.29 (0.79) | 0.507 |
| **Hallucinogen** | 11 | 3 | 4 | 1 |  |
| **DU** | 1.94 (0.31) | 1.73 (0.57) | 2.18 (0.57) | 0.14 (NA) | NA |
| **EC** | 0.61 (0.40) | 0.34 (0.65) | 0.81 (0.27) | 0.37 (NA) | NA |
| **MDD** | 315 | 304 | 216 | 176 |  |
| **DU** | 1.27 (0.92) | 1.01 (1.02) | 0.69 (1.14) | 0.68 (1.11) | <0.001 |
| **EC** | 0.73 (0.81) | 0.68 (0.76) | 0.82 (0.84) | 0.60 (0.84) | 0.061 |
| **GAD** | 136 | 141 | 97 | 88 |  |
| **DU** | 1.24 (0.91) | 0.97 (0.99) | 0.67 (1.10) | 0.48 (1.08) | <0.001 |
| **EC** | 0.81 (0.81) | 0.69 (0.76) | 0.87 (0.88) | 0.60 (0.84) | 0.096 |
| **Social** | 68 | 96 | 44 | 58 |  |
| **DU** | 1.03 (0.92) | 0.90 (0.95) | 0.53 (1.07) | 0.61 (1.05) | 0.021 |
| **EC** | 0.77 (0.81) | 0.69 (0.78) | 0.67 (0.88) | 0.51 (0.83) | 0.345 |
| **Panic** | 73 | 40 | 48 | 28 |  |
| **DU** | 1.37 (0.87) | 0.85 (1.07) | 0.80 (1.20) | 0.44 (1.17) | <0.001 |
| **EC** | 0.78 (0.91) | 0.63 (0.65) | 0.81 (0.90) | 0.48 (0.62) | 0.262 |
| **PTSD** | 77 | 46 | 52 | 28 |  |
| **DU** | 1.29 (0.90) | 1.15 (1.01) | 0.76 (1.18) | 0.64 (1.11) | 0.005 |
| **EC** | 0.81 (0.87) | 0.69 (0.72) | 0.89 (0.84) | 0.59 (0.82) | 0.384 |

**Supplementary Table 26.** Results of Linear mixed effects models predicting *DU* when accounting for effects of group, time and their interaction.

|  | **Group** | **Time** | **Group x Time** |
| --- | --- | --- | --- |
| **Alcohol** | $F(1, 298)=9.02, p=0.003$<br>HCs=0.802; Alcohol=1.171<br>HCs - Alcohol: $t(278.45)=-3.365, p<0.001, d=-0.57$ | $F(1, 208)=40.32, p<0.001$<br>T1=1.107; T2=0.682<br>T1 - T2: $t(202.21)=6.272, p<0.001, d=0.66$ | $F(1, 208)=3.32, p=0.070$ |
| **Cannabis** | $F(1, 333)=7.39, p=0.007$<br>HCs=0.805; Cannabis=1.082<br>HCs - Cannabis: $t(311.22)=-2.776, p=0.006, d=-0.41$ | $F(1, 234)=21.05, p<0.001$<br>T1=1.049; T2=0.731<br>T1 - T2: $t(231.06)=4.675, p<0.001, d=0.47$ | $F(1, 234)=0.02, p=0.890$ |
| **Stimulant** | $F(1, 429)=12.91, p<0.001$<br>HCs=0.805; Stimulant=1.134<br>HCs - Stimulant: $t(409.96)=-3.77, p<0.001, d=-0.46$ | $F(1, 311)=36.01, p<0.001$<br>T1=1.138; T2=0.77<br>T1 - T2: $t(321.21)=6.122, p<0.001, d=0.52$ | $F(1, 311)=0.30, p=0.587$ |
| **Opioid** | $F(1, 293)=9.31, p=0.002$<br>HCs=0.799; Opioid=1.141<br>HCs - Opioid: $t(278.14)=-3.293, p=0.001, d=-0.53$ | $F(1, 216)=36.16, p<0.001$<br>T1=1.091; T2=0.697<br>T1 - T2: $t(211.86)=5.989, p<0.001, d=0.61$ | $F(1, 216)=1.52, p=0.218$ |
| **Sedative** | $F(1, 282)=4.42, p=0.036$<br>HCs=0.802; Sedative=1.083<br>HCs - Sedative: $t(257.17)=-2.341, p=0.020, d=-0.44$ | $F(1, 191)=22.92, p<0.001$<br>T1=1.028; T2=0.656<br>T1 - T2: $t(178.76)=5.17, p<0.001, d=0.58$ | $F(1, 191)=0.87, p=0.353$ |
| **Hallucinogen** | $F(1, 228)=16.18, p<0.001$<br>HCs=0.79; Hallucinogen=1.83<br>HCs - Hallucinogen: $t(210.11)=-4.143, p<0.001, d=-1.65$ | $F(1, 163)=1.65, p=0.201$ | $F(1, 163)=0.20, p=0.655$ |
| **MDD** | $F(1, 753)=4.45, p=0.035$<br>HCs=0.796; MDD=0.977<br>HCs - MDD: $t(736.85)=-2.239, p=0.025, d=-0.26$ | $F(1, 588)=53.19, p<0.001$<br>T1=1.096; T2=0.701<br>T1 - T2: $t(607.18)=9.344, p<0.001, d=0.56$ | $F(1, 588)=0.73, p=0.393$ |
| **GAD** | $F(1, 431)=1.49, p=0.222$ | $F(1, 343)=55.12, p<0.001$<br>T1=1.036; T2=0.63<br>T1 - T2: $t(346.3)=7.979, p<0.001, d=0.62$ | $F(1, 343)=1.53, p=0.217$ |
| **Social** | $F(1, 320)=0.14, p=0.712$ | $F(1, 258)=24.37, p<0.001$<br>T1=0.938; T2=0.621<br>T1 - T2: $t(258.69)=4.942, p<0.001, d=0.45$ | $F(1, 258)=0.04, p=0.851$ |
| **Panic** | $F(1, 271)=3.47, p=0.064$ | $F(1, 219)=31.94, p<0.001$<br>T1=1.034; T2=0.661<br>T1 - T2: $t(218.59)=5.614, p<0.001, d=0.55$ | $F(1, 219)=0.71, p=0.399$ |
| **PTSD** | $F(1, 282)=4.08, p=0.044$<br>HCs=0.791; PTSD=1.022<br>HCs - PTSD: $t(274.13)=-2.201, p=0.029, d=-0.34$ | $F(1, 227)=39.79, p<0.001$<br>T1=1.059; T2=0.65<br>T1 - T2: $t(226.04)=6.19, p<0.001, d=0.6$ | $F(1, 227)=2.06, p=0.152$ |

**Note:** T1 = Baseline; T2 = Follow-up; NS = nonsignificant.
For interpretability, sum coding was used for time (baseline = -1; follow-up = 1), and group (with HCs coded as -1).

**Supplementary Table 27.** Results of Linear mixed effects models predicting *DU* when accounting for effects of group, time, age and sex.

|  | **Group** | **Time** | **Age** | **Sex** | **Group x Age** | **Group x Sex** |
| --- | --- | --- | --- | --- | --- | --- |
| **Alcohol** | $F(1, 271)=11.86, p<0.001$<br>HCs=0.792; Alcohol=1.166<br>HCs - Alcohol: $t(273.14)=-3.34, p<0.001, d=-0.58$ | $F(1, 202)=40.11, p<0.001$<br>T1=1.11; T2=0.679<br>T1 - T2: $t(202.06)=6.333, p<0.001, d=0.66$ | $F(1, 247)=0.36, p=0.547$ | $F(1, 250)=2.28, p=0.133$ | $F(1, 276)=2.99, p=0.085$ | $F(1, 270)=0.64, p=0.425$ |
| **Cannabis** | $F(1, 304)=8.83, p=0.003$<br>HCs=0.801; Cannabis=1.075<br>HCs - Cannabis:<br>$t(306.27)=-2.739, p=0.007, d=-0.4$ | $F(1, 228)=23.64, p<0.001$<br>T1=1.052; T2=0.723<br>T1 - T2:<br>$t(228.41)=4.862, p<0.001, d=0.48$ | $F(1, 277)=0.22, p=0.636$ | $F(1, 280)=2.23, p=0.136$ | $F(1, 307)=0.74, p=0.390$ | $F(1, 305)=1.67, p=0.198$ |
| **Stimulant** | $F(1, 409)=15.92, p<0.001$<br>HCs=0.815; Stimulant=1.127<br>HCs - Stimulant:<br>$t(405.97)=-3.606, p<0.001, d=-0.44$ | $F(1, 320)=40.45, p<0.001$<br>T1=1.142; T2=0.759<br>T1 - T2:<br>$t(320.14)=6.36, p<0.001, d=0.54$ | $F(1, 378)=0.28, p=0.594$ | $F(1, 383)=2.32, p=0.128$ | $F(1, 409)=3.44, p=0.064$ | $F(1, 410)=1.92, p=0.166$ |
| **Opioid** | $F(1, 269)=13.35, p<0.001$<br>HCs=0.795; Opioid=1.144<br>HCs - Opioid:<br>$t(271.81)=-3.366, p<0.001, d=-0.54$ | $F(1, 212)=36.72, p<0.001$<br>T1=1.098; T2=0.698<br>T1 - T2:<br>$t(212.93)=6.06, p<0.001, d=0.62$ | $F(1, 252)=0.33, p=0.565$ | $F(1, 255)=2.40, p=0.122$ | $F(1, 274)=1.45, p=0.230$ | $F(1, 274)=1.91, p=0.169$ |
| **Sedative** | $F(1, 240)=9.47, p=0.002$<br>HCs=0.804; Sedative=1.13 | $F(1, 176)=27.77, p<0.001$<br>T1=1.045; T2=0.667 | $F(1, 215)=0.30, p=0.583$ | $F(1, 218)=2.37, p=0.583$ | $F(1, 248)=1.77, p=0.184$ | $F(1, 246)=1.53, p=0.218$ |
| | HCs -<br>Sedative:<br>$t(244.11)=-2.737$,<br>$p=0.007$, $d=-0.51$ | T1 - T2:<br>$t(176.59)=5.27$,<br>$p<0.001$,<br>$d=0.59$ | | | | |
| **Hallucino-**<br>**nogen** | $F(1, 190)=17.22$,<br>$p<0.001$<br>HCs=0.791;<br>Hallucinogen=<br>1.786<br>HCs -<br>Hallucinogen:<br>$t(178.98)=-4.036$,<br>$p<0.001$, $d=-1.58$ | $F(1, 138)=16.97$,<br>$p<0.001$<br>T1=0.992;<br>T2=0.668<br>T1 - T2:<br>$t(138.55)=4.12$,<br>$p<0.001$,<br>$d=0.51$ | $F(1, 158)=0.24$,<br>$p=0.623$ | $F(1, 160)=2.46$,<br>$p=0.119$ | $F(1, 211)=0.00$,<br>$p=0.946$ | $F(1, 182)=0.34$,<br>$p=0.563$ |
| **MDD** | $F(1, 734)=4.87$,<br>$p=0.028$<br>HCs=0.822;<br>MDD=0.967<br>HCs - MDD:<br>$t(724.21)=-1.825$,<br>$p=0.068$, $d=-0.21$ | $F(1, 618)=97.98$,<br>$p<0.001$<br>T1=1.1;<br>T2=0.681<br>T1 - T2:<br>$t(618.71)=9.898$,<br>$p<0.001$,<br>$d=0.59$ | $F(1, 719)=0.32$,<br>$p=0.572$ | $F(1, 725)=2.23$,<br>$p=0.136$ | $F(1, 730)=9.10$,<br>$p=0.003$<br>HCs=0.004;<br>MDD=0.025<br>HCs - MDD:<br>$t(730)=-3.016$,<br>$p=0.003$, $d=-0.03$ | $F(1, 739)=1.12$,<br>$p=0.289$ |
| **GAD** | $F(1, 423)=1.47$,<br>$p=0.227$<br>HCs=0.822;<br>GAD=0.894;<br>HCs - GAD:<br>$t(416.47)=-0.809$,<br>$p=0.419$, $d=-0.11$ | $F(1, 352)=69.52$,<br>$p<0.001$<br>T1=1.036;<br>T2=0.612<br>T1 - T2:<br>$t(352.89)=8.338$,<br>$p<0.001$,<br>$d=0.66$ | $F(1, 412)=0.35$,<br>$p=0.552$ | $F(1, 415)=2.27$,<br>$p=0.133$ | $F(1, 421)=6.52$,<br>$p=0.011$<br>HCs=0.004;<br>GAD=0.025;<br>HCs - GAD:<br>$t(422)=-2.55$,<br>$p=0.011$, $d=-0.03$ | $F(1, 425)=0.53$,<br>$p=0.467$ |
| **Social** | $F(1, 309)=1.86$,<br>$p=0.174$ | $F(1, 263)=28.39$,<br>$p<0.001$<br>T1=0.959;<br>T2=0.618<br>T1 - T2:<br>$t(263.75)=5.328$,<br>$p<0.001$,<br>$d=0.49$ | $F(1, 294)=0.25$,<br>$p=0.614$ | $F(1, 298)=2.48$,<br>$p=0.116$ | $F(1, 304)=11.66$,<br>$p<0.001$<br>HCs=0.003;<br>Social=0.034;<br>HCs - GAD:<br>$t(305)=-3.41$,<br>$p<0.001$, $d=-0.045$ | $F(1, 309)=0.19$,<br>$p=0.659$ |
| **Panic** | $F(1, 265)=4.33$,<br>$p=0.038$<br>HCs=0.805;<br>Panic=0.968<br>HCs - Panic:<br>$t(261.82)=-1.531$,<br>$p=0.127$, $d=-0.24$ | $F(1, 224)=35.23$,<br>$p<0.001$<br>T1=1.027;<br>T2=0.631<br>T1 - T2:<br>$t(224.13)=5.935$,<br>$p<0.001$,<br>$d=0.59$ | $F(1, 252)=0.31$,<br>$p=0.576$ | $F(1, 255)=2.37$,<br>$p=0.125$ | $F(1, 256)=12.45$,<br>$p<0.001$<br>HCs=0.003;<br>Panic=0.037<br>HCs - Panic:<br>$t(257)=-3.53$,<br>$p<0.001$, $d=-0.050$ | $F(1, 266)=1.80$,<br>$p=0.181$ |
| **PTSD** | $F(1, 276)=3.38$,<br>$p=0.067$ | $F(1, 230)=40.31$,<br>$p<0.001$<br>T1=1.058;<br>T2=0.634<br>T1 - T2:<br>$t(230.35)=6.349$,<br>$p<0.001$,<br>$d=0.62$ | $F(1, 261)=0.34$,<br>$p=0.559$ | $F(1, 264)=2.33$,<br>$p=0.128$ | $F(1, 274)=3.58$,<br>$p=0.059$ | $F(1, 277)=0.07$,<br>$p=0.794$ |

**Note:** T1 = Baseline; T2 = Follow-up; NS = nonsignificant.
For interpretability, age was centered, and sum coding was used for sex (female = -1; male = 1), time (baseline = -1; follow-up = 1), and group (with HCs coded as -1).

**Supplementary Table 28.** Results of Linear mixed effects models predicting *EC* when accounting for effects of group, time and their interaction.

|  | **Group** | **Time** | **Group x Time** |
| --- | --- | --- | --- |
| **Alcohol** | $F(1, 297)=43.00$, $p<0.001$<br>HCs=1.022; Alcohol=0.421 | $F(1, 200)=2.65$,<br>$p=0.105$ | $F(1, 200)=0.12$,<br>$p=0.729$ |
| | HCS - Alcohol: $t(279.2)=6.662$,<br>$p<0.001$, $d=1.28$ | | |
| **Cannabis** | $F(1, 335)=40.56$, $p<0.001$<br>HCS=1.021; Cannabis=0.467<br>HCS - Cannabis: $t(314.53)=6.393$,<br>$p<0.001$, $d=1.14$ | $F(1, 218)=1.56$,<br>$p=0.213$ | $F(1, 218)=0.69$,<br>$p=0.409$ |
| **Stimulant** | $F(1, 436)=60.93$, $p<0.001$<br>HCS=1.021; Stimulant=0.461<br>HCS - Stimulant: $t(416.83)=7.827$,<br>$p<0.001$, $d=1.12$ | $F(1, 296)=2.92$,<br>$p=0.089$ | $F(1, 296)=0.55$,<br>$p=0.461$ |
| **Opioid** | $F(1, 294)=37.67$, $p<0.001$<br>HCS=1.023; Opioid=0.469<br>HCS - Opioid: $t(279.17)=6.23$,<br>$p<0.001$, $d=1.1$ | $F(1, 210)=3.07$,<br>$p=0.081$ | $F(1, 210)=0.04$,<br>$p=0.839$ |
| **Sedative** | $F(1, 279)=37.90$, $p<0.001$<br>HCS=1.022; Sedative=0.358<br>HCS - Sedative: $t(256.41)=6.427$,<br>$p<0.001$, $d=1.38$ | $F(1, 181)=4.21$,<br>$p=0.042$<br>T1=0.778;<br>T2=0.899<br>T1 - T2:<br>$t(171.43)=-2.209$,<br>$p=0.029$, $d=-0.25$ | $F(1, 181)=0.19$,<br>$p=0.663$ |
| **Hallucinogen** | $F(1, 219)=2.25$, $p=0.135$ | $F(1, 147)=1.34$,<br>$p=0.249$ | $F(1, 147)=0.20$,<br>$p=0.653$ |
| **MDD** | $F(1, 759)=20.53$, $p<0.001$<br>HCS=1.024; MDD=0.727<br>HCS - MDD: $t(742.95)=4.492$,<br>$p<0.001$, $d=0.56$ | $F(1, 577)=4.40$,<br>$p=0.036$<br>T1=0.764;<br>T2=0.829<br>T1 - T2:<br>$t(592.98)=-2.059$,<br>$p=0.04$, $d=-0.12$ | $F(1, 577)=0.43$,<br>$p=0.510$ |
| **GAD** | $F(1, 434)=12.51$, $p<0.001$<br>HCS=1.025; GAD=0.759 | $F(1, 333)=2.33$,<br>$p=0.128$ | $F(1, 333)=1.24$,<br>$p=0.267$ |
| | HCs - GAD: $t(425.25)=3.463$,<br>$p<0.001$, $d=0.55$ | | |
| **Social** | $F(1, 324)=15.56$, $p<0.001$<br>HCs=1.035; Social=0.692<br>HCs - Social: $t(324.47)=3.945$,<br>$p<0.001$, $d=0.66$ | $F(1, 246)=0.17$,<br>$p=0.679$ | $F(1, 246)=3.10$,<br>$p=0.079$ |
| **Panic** | $F(1, 272)=9.33$, $p=0.002$<br>HCs=1.026; Panic=0.733<br>HCs - Panic: $t(266.47)=3.003$,<br>$p=0.003$, $d=0.58$ | $F(1, 212)=1.33$,<br>$p=0.249$ | $F(1, 212)=0.78$,<br>$p=0.378$ |
| **PTSD** | $F(1, 283)=8.03$, $p=0.005$<br>HCs=1.026; PTSD=0.767<br>HCs - PTSD: $t(275.73)=2.776$,<br>$p=0.006$, $d=0.48$ | $F(1, 221)=1.20$,<br>$p=0.275$ | $F(1, 221)=0.77$,<br>$p=0.380$ |

**Note:** T1 = Baseline; T2 = Follow-up; NS = nonsignificant.
For interpretability, sum coding was used for time (baseline = -1; follow-up = 1), and group (with HCs coded as -1).

**Supplementary Table 29.**
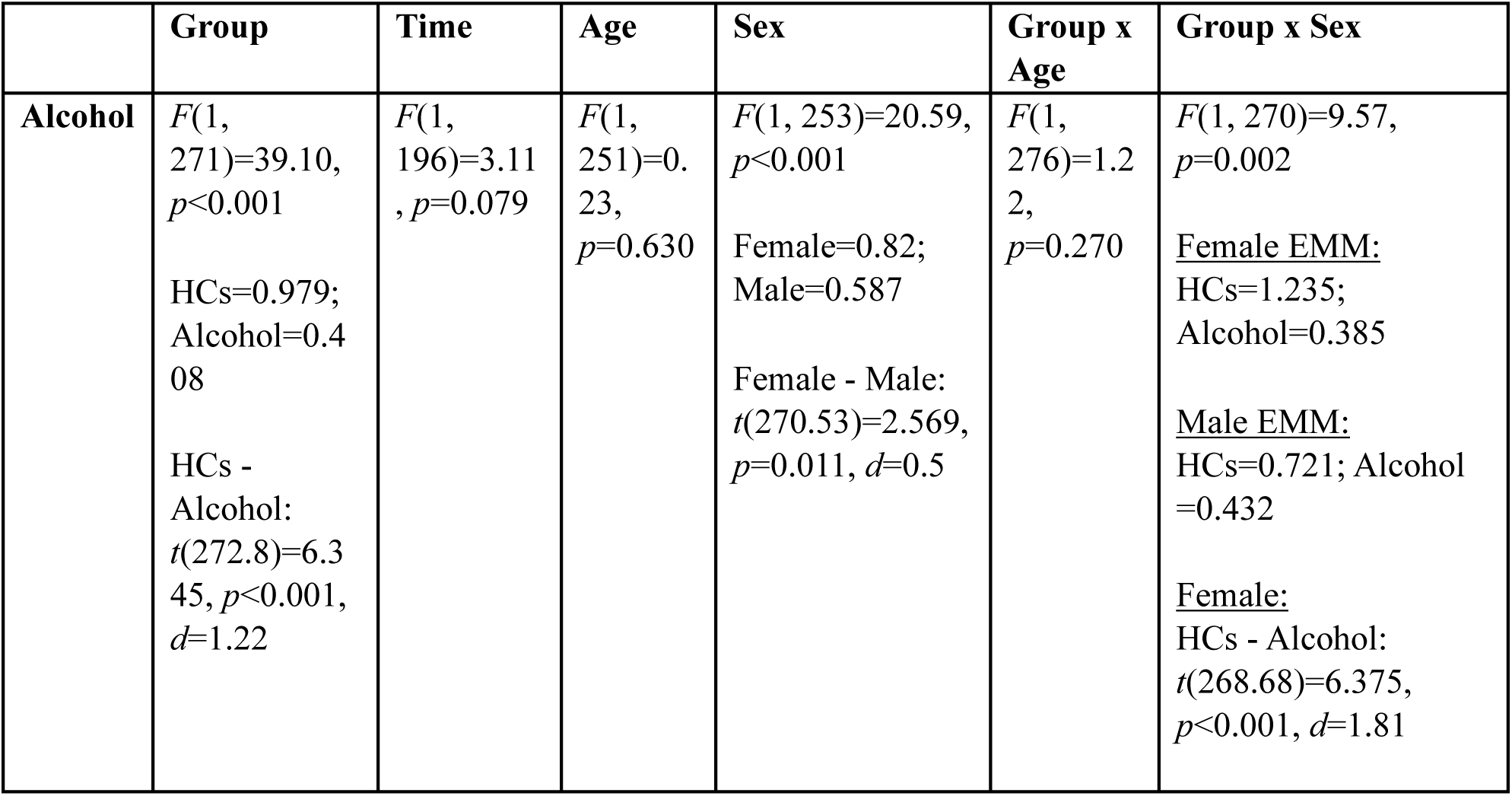
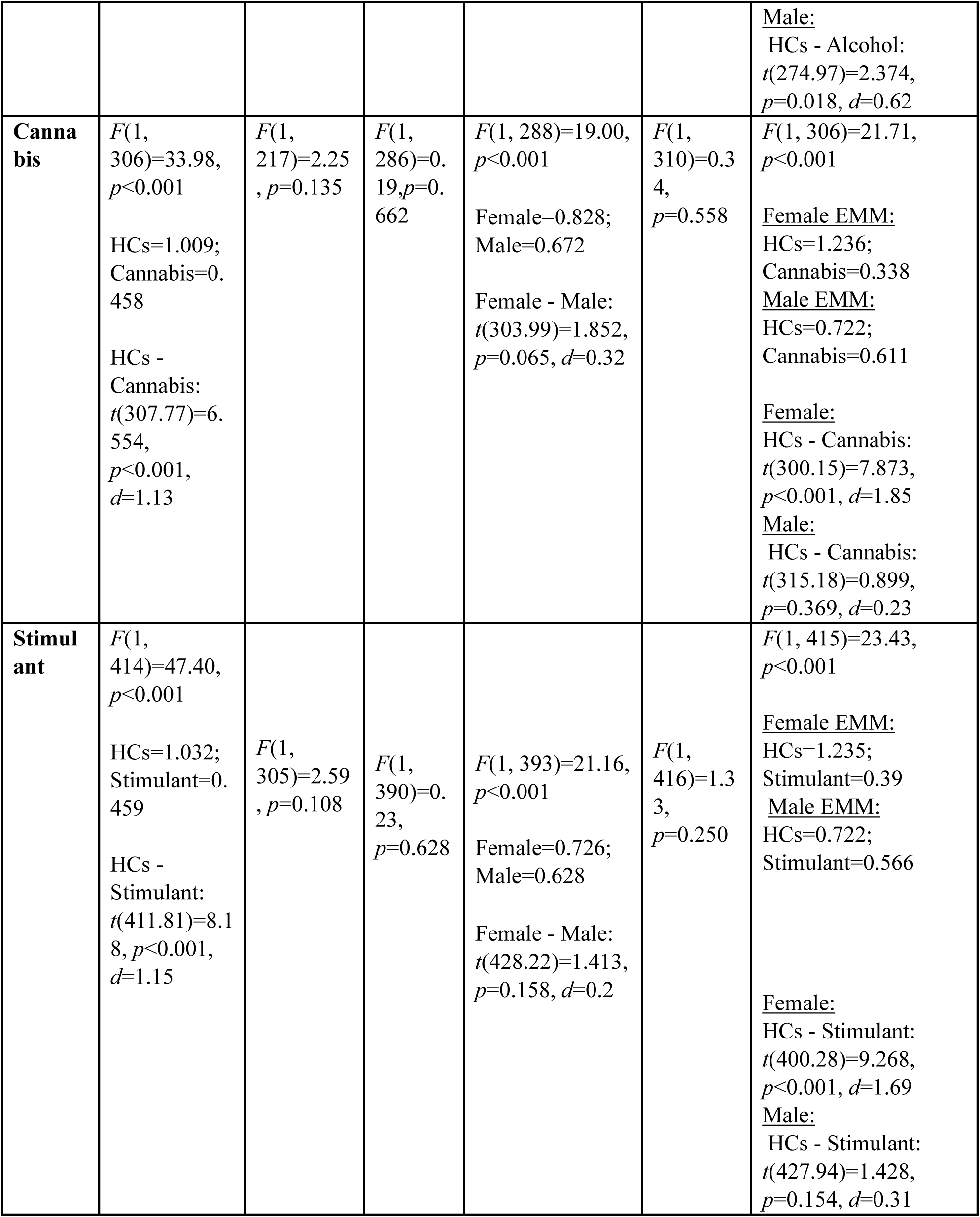
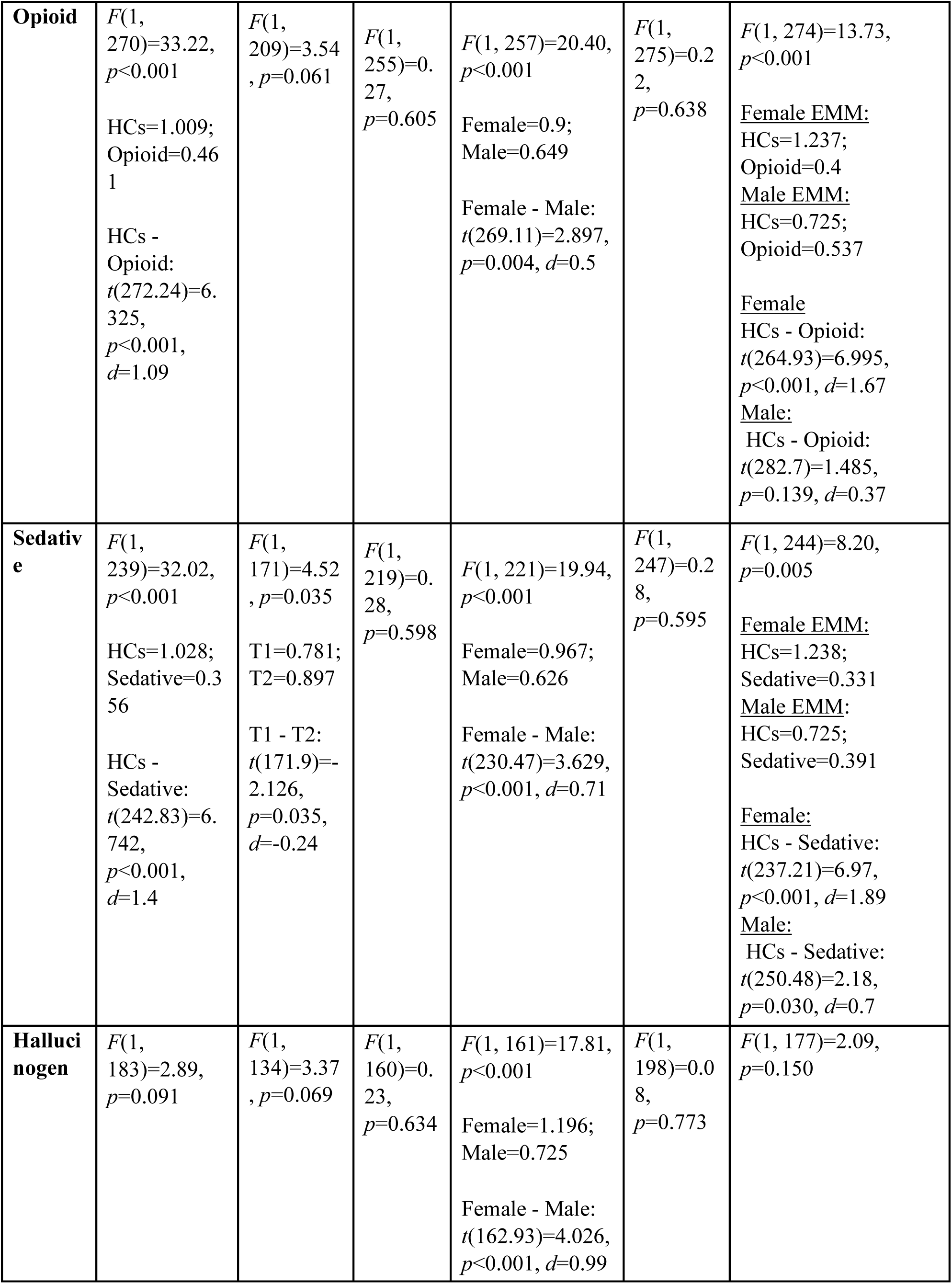
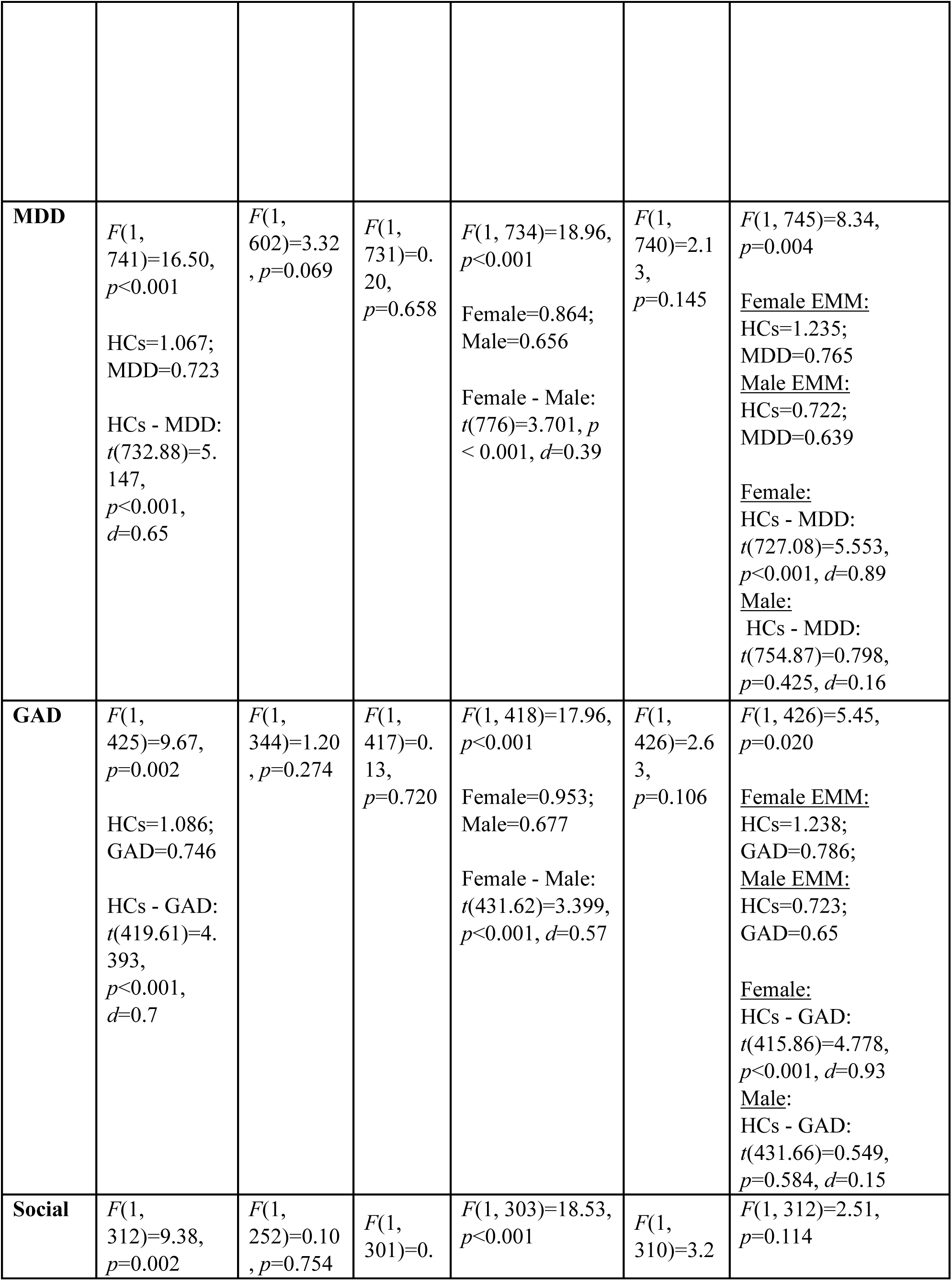
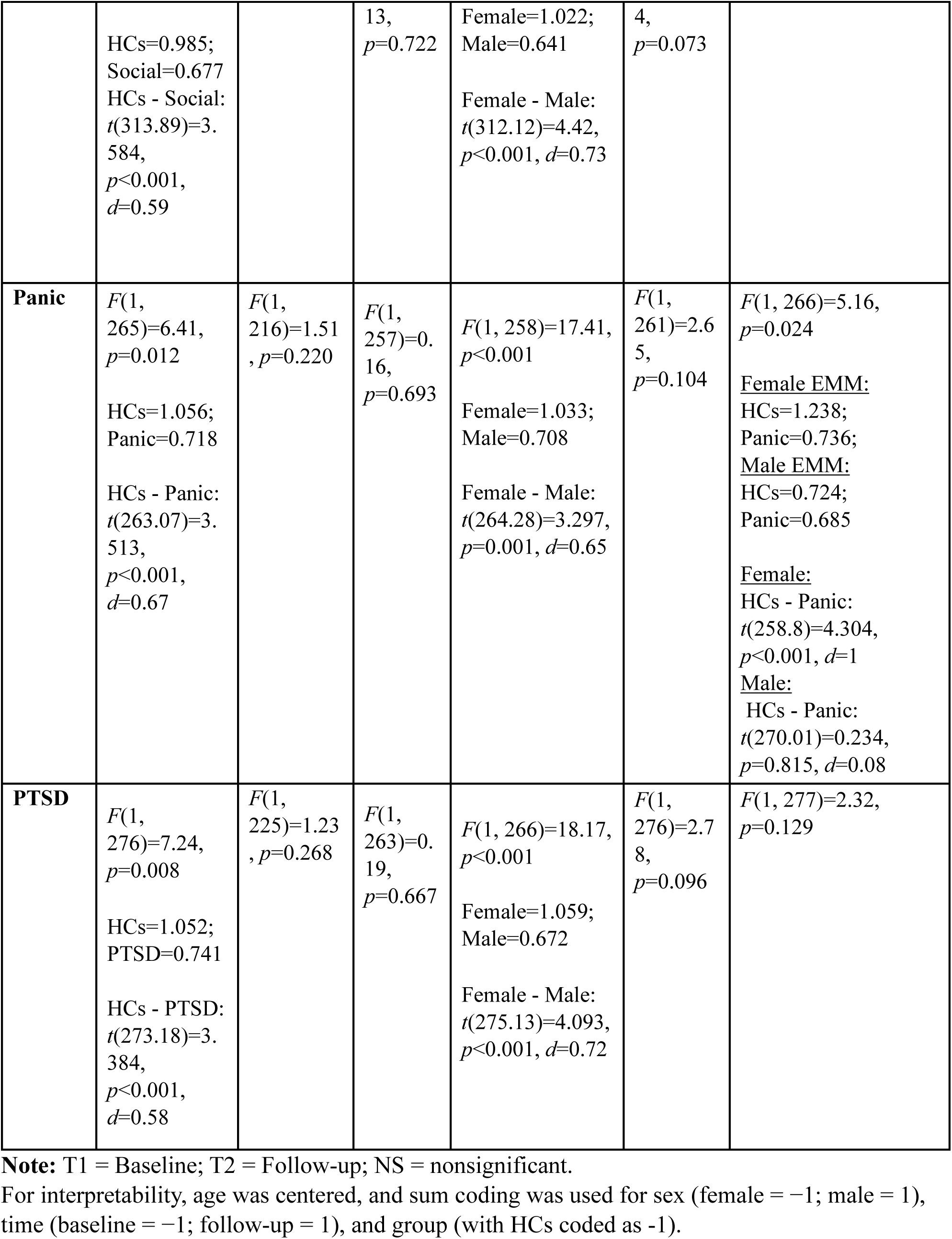
Results of Linear mixed effects models predicting *EC* when accounting for effects of group, time, age and sex.

**Supplementary Table 30.** Performance metrics for predictive categorization of individuals with affective disorders (no comorbid SUDs) and those with SUDs (but no comorbid affective disorders)

| Predictor | Method | AUC | Balanced Accuracy | Sensitivity | Specificity |
| --- | --- | --- | --- | --- | --- |
| Age, Sex | ENET | 0.495 | 0.477 | 0.300 | 0.653 |
| Age, Sex | KNN | 0.600 | 0.569 | 0.600 | 0.538 |
| Age, Sex | ADABAG | 0.565 | 0.551 | 0.740 | 0.363 |
| Age, Sex | STACK | 0.617 | 0.551 | 0.720 | 0.381 |
| Model parameters | ENET | 0.752 | 0.648 | 0.609 | 0.688 |
| Model parameters | KNN | 0.620 | 0.596 | 0.607 | 0.584 |
| Model parameters | ADABAG | 0.675 | 0.584 | 0.505 | 0.663 |
| Model parameters | STACK | 0.699 | 0.647 | 0.587 | 0.706 |
| Model parameters, Age, Sex | ENET | 0.763 | 0.694 | 0.731 | 0.656 |
| Model parameters, Age, Sex | KNN | 0.670 | 0.632 | 0.711 | 0.553 |
| Model parameters, Age, Sex | ADABAG | 0.692 | 0.658 | 0.569 | 0.747 |
| Model parameters, Age, Sex | STACK | 0.740 | 0.704 | 0.707 | 0.700 |
| All parameters | ENET | 0.720 | 0.673 | 0.680 | 0.666 |
| All parameters | KNN | 0.549 | 0.530 | 0.560 | 0.500 |
| All parameters | ADABAG | 0.662 | 0.577 | 0.360 | 0.794 |
| All parameters | STACK | 0.668 | 0.571 | 0.420 | 0.722 |
| All parameters, Age, Sex | **ENET** | **0.773** | **0.714** | **0.762** | **0.666** |
| All parameters, Age, Sex | KNN | 0.573 | 0.553 | 0.604 | 0.503 |
| All parameters, Age, Sex | ADABAG | 0.668 | 0.629 | 0.545 | 0.713 |
| All parameters, Age, Sex | STACK | 0.700 | 0.626 | 0.565 | 0.688 |

**Note:** Model parameters = *EC* and *DU* at baseline and follow-up; All parameters = *EC* and *DU* (at baseline and follow-up) and *choice uncertainty*-RT correlation coefficients. The model with the highest balanced accuracy is bolded.

**Supplementary Table 31.** Performance metrics for predictive categorization of individuals with and without SUDs.

| Predictor | Method | AUC | Balanced Accuracy | Sensitivity | Specificity |
| --- | --- | --- | --- | --- | --- |
| Age, Sex | ENET | 0.537 | 0.495 | 0.406 | 0.583 |
| Age, Sex | KNN | 0.606 | 0.572 | 0.642 | 0.502 |
| Age, Sex | ADABAG | 0.603 | 0.552 | 0.704 | 0.400 |
| Age, Sex | Stack | 0.605 | 0.583 | 0.728 | 0.437 |
| Model parameters | ENET | 0.680 | 0.620 | 0.612 | 0.628 |
| Model parameters | KNN | 0.582 | 0.585 | 0.587 | 0.582 |
| Model parameters | ADABAG | 0.664 | 0.621 | 0.592 | 0.649 |
| Model parameters | Stack | 0.658 | 0.622 | 0.605 | 0.640 |
| Model parameters, Age, Sex | ENET | 0.719 | 0.677 | 0.713 | 0.640 |
| Model parameters, Age, Sex | KNN | 0.692 | 0.655 | 0.736 | 0.573 |
| Model parameters, Age, Sex | ADABAG | 0.682 | 0.639 | 0.683 | 0.595 |
| Model parameters, Age, Sex | Stack | 0.713 | 0.657 | 0.712 | 0.601 |
| All parameters | **ENET** | **0.761** | **0.694** | **0.653** | **0.734** |
| All parameters | KNN | 0.669 | 0.623 | 0.605 | 0.642 |
| All parameters | ADABAG | 0.722 | 0.660 | 0.773 | 0.548 |
| All parameters | Stack | 0.735 | 0.673 | 0.713 | 0.633 |
| All parameters, Age, Sex | ENET | 0.753 | 0.684 | 0.683 | 0.684 |
| All parameters, Age, Sex | KNN | 0.656 | 0.628 | 0.689 | 0.567 |
| All parameters, Age, Sex | ADABAG | 0.694 | 0.632 | 0.641 | 0.622 |
| All parameters, Age, Sex | Stack | 0.722 | 0.691 | 0.719 | 0.662 |

**Note:** Model parameters = *EC* and *DU* at baseline and follow-up; All parameters = *EC* and *DU* (at baseline and follow-up) and *choice uncertainty*-RT correlation coefficients. The model with the highest balanced accuracy is bolded.

**Supplementary Table 32.** Performance metrics for predictive categorization of individuals with and without affective disorders.

| Predictor | Method | AUC | Balanced Accuracy | Sensitivity | Specificity |
| --- | --- | --- | --- | --- | --- |
| Age, Sex | ENET | 0.447 | 0.473 | 0.352 | 0.594 |
| Age, Sex | KNN | 0.443 | 0.492 | 0.511 | 0.473 |
| Age, Sex | ADABAG | 0.519 | 0.507 | 0.844 | 0.170 |
| Age, Sex | Stack | 0.456 | 0.467 | 0.721 | 0.213 |
| Model parameters | **ENET** | **0.748** | **0.688** | **0.756** | **0.619** |
| Model parameters | KNN | 0.627 | 0.623 | 0.674 | 0.571 |
| Model parameters | ADABAG | 0.661 | 0.595 | 0.468 | 0.722 |
| Model parameters | Stack | 0.714 | 0.622 | 0.520 | 0.725 |
| **Model parameters, Age, Sex** | ENET | 0.625 | 0.610 | 0.624 | 0.596 |
| **Model parameters, Age, Sex** | KNN | 0.506 | 0.511 | 0.476 | 0.546 |
| **Model parameters, Age, Sex** | ADABAG | 0.617 | 0.539 | 0.406 | 0.672 |
| **Model parameters, Age, Sex** | STACK | 0.592 | 0.524 | 0.389 | 0.658 |
| **All parameters** | ENET | 0.630 | 0.596 | 0.561 | 0.632 |
| **All parameters** | KNN | 0.477 | 0.448 | 0.441 | 0.455 |
| **All parameters** | ADABAG | 0.526 | 0.508 | 0.274 | 0.743 |
| **All parameters** | STACK | 0.543 | 0.509 | 0.309 | 0.708 |
| **All parameters, Age, Sex** | ENET | 0.629 | 0.575 | 0.533 | 0.616 |
| **All parameters, Age, Sex** | KNN | 0.531 | 0.533 | 0.517 | 0.549 |
| **All parameters, Age, Sex** | ADABAG | 0.579 | 0.533 | 0.633 | 0.432 |
| **All parameters, Age, Sex** | STACK | 0.570 | 0.563 | 0.583 | 0.543 |

**Note:** Model parameters = *EC* and *DU* at baseline and follow-up; All parameters = *EC* and *DU* (at baseline and follow-up) and *choice uncertainty*-RT correlation coefficients. The model with the highest balanced accuracy is bolded.

**Supplementary Table 33.**
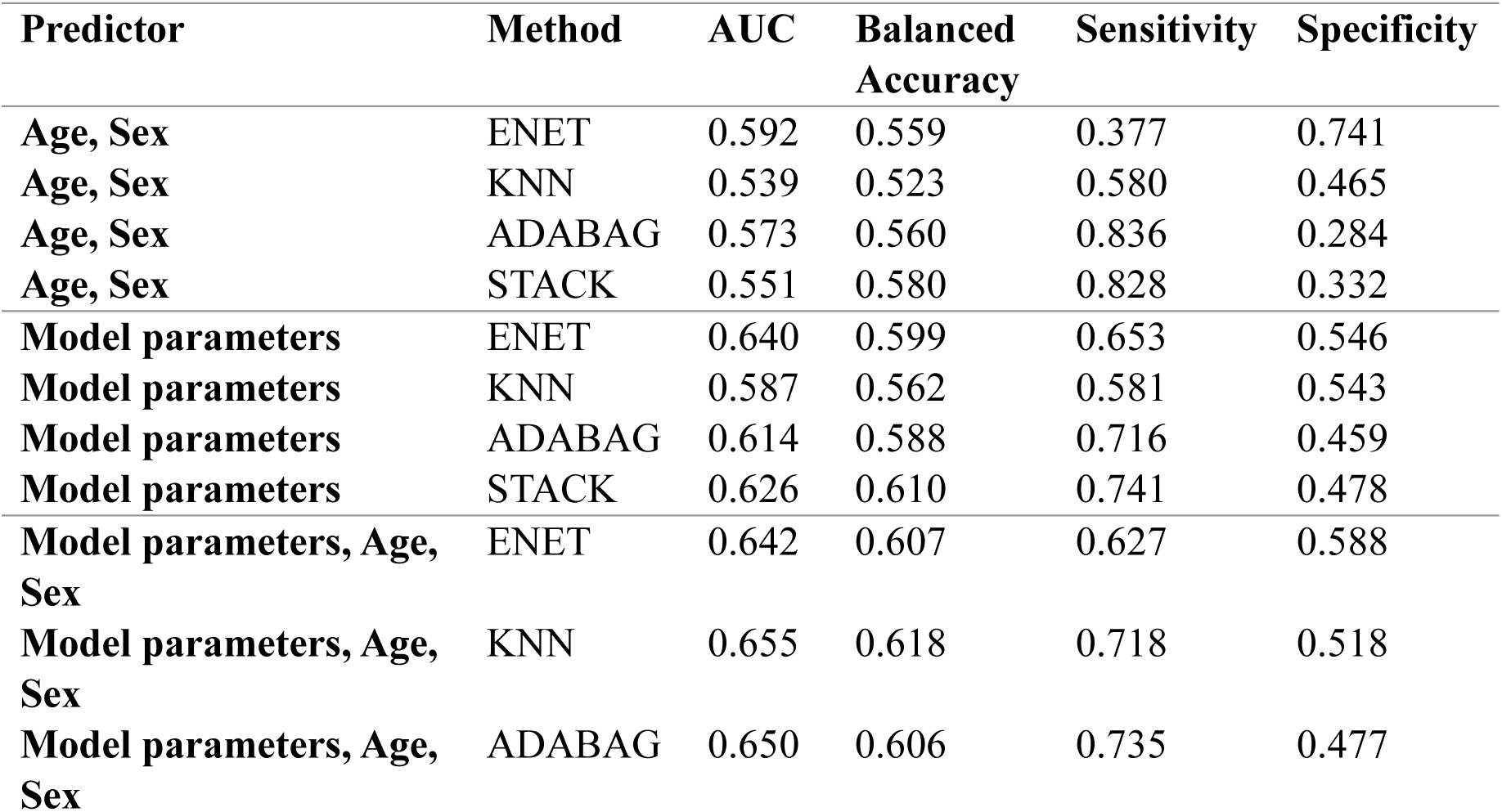
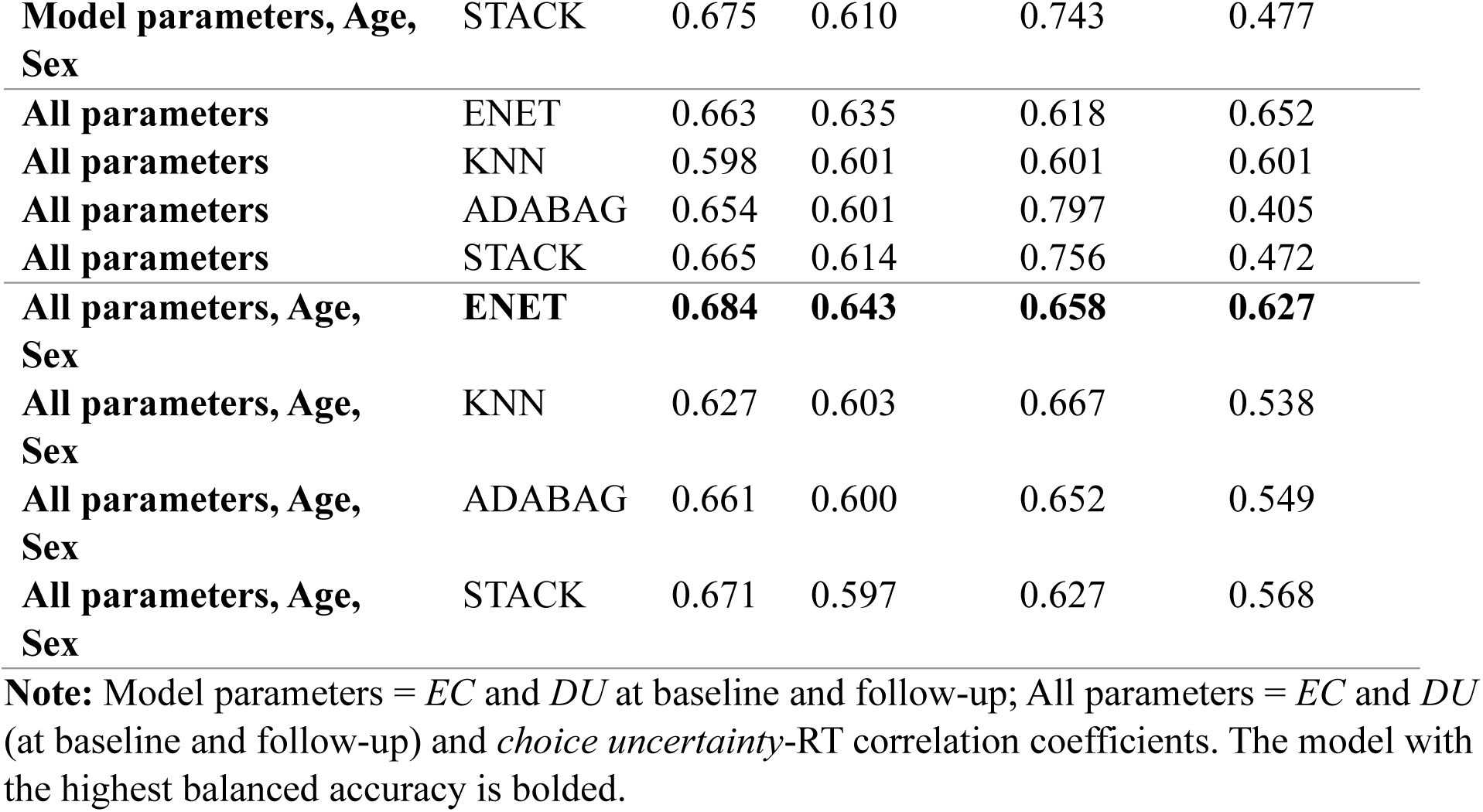
Performance metrics for predictive categorization of individuals with and without comorbid affective and substance use disorders.

| Predictor | Method | AUC | Balanced Accuracy | Sensitivity | Specificity |
| --- | --- | --- | --- | --- | --- |
| **Age, Sex** | ENET | 0.592 | 0.559 | 0.377 | 0.741 |
| **Age, Sex** | KNN | 0.539 | 0.523 | 0.580 | 0.465 |
| **Age, Sex** | ADABAG | 0.573 | 0.560 | 0.836 | 0.284 |
| **Age, Sex** | STACK | 0.551 | 0.580 | 0.828 | 0.332 |
| **Model parameters** | ENET | 0.640 | 0.599 | 0.653 | 0.546 |
| **Model parameters** | KNN | 0.587 | 0.562 | 0.581 | 0.543 |
| **Model parameters** | ADABAG | 0.614 | 0.588 | 0.716 | 0.459 |
| **Model parameters** | STACK | 0.626 | 0.610 | 0.741 | 0.478 |
| **Model parameters, Age, Sex** | ENET | 0.642 | 0.607 | 0.627 | 0.588 |
| **Model parameters, Age, Sex** | KNN | 0.655 | 0.618 | 0.718 | 0.518 |
| **Model parameters, Age, Sex** | ADABAG | 0.650 | 0.606 | 0.735 | 0.477 |
| **Model parameters, Age, Sex** | STACK | 0.675 | 0.610 | 0.743 | 0.477 |
| **All parameters** | ENET | 0.663 | 0.635 | 0.618 | 0.652 |
| **All parameters** | KNN | 0.598 | 0.601 | 0.601 | 0.601 |
| **All parameters** | ADABAG | 0.654 | 0.601 | 0.797 | 0.405 |
| **All parameters** | STACK | 0.665 | 0.614 | 0.756 | 0.472 |
| **All parameters, Age, Sex** | **ENET** | **0.684** | **0.643** | **0.658** | **0.627** |
| **All parameters, Age, Sex** | KNN | 0.627 | 0.603 | 0.667 | 0.538 |
| **All parameters, Age, Sex** | ADABAG | 0.661 | 0.600 | 0.652 | 0.549 |
| **All parameters, Age, Sex** | STACK | 0.671 | 0.597 | 0.627 | 0.568 |

**Note:** Model parameters = *EC* and *DU* at baseline and follow-up; All parameters = *EC* and *DU* (at baseline and follow-up) and *choice uncertainty*-RT correlation coefficients. The model with the highest balanced accuracy is bolded.

## References

Aupperle, R. L., Melrose, A. J., Francisco, A., Paulus, M. P., & Stein, M. B. (2015). Neural substrates of approach-avoidance conflict decision-making. Human brain mapping, 36(2), 449–462.

Aupperle, R. L., & Paulus, M. P. (2010). Neural systems underlying approach and avoidance in anxiety disorders. Dialogues Clin Neurosci, 12(4), 517–531. 10.31887/DCNS.2010.12.4/raupperle

Aupperle, R. L., Sullivan, S., Melrose, A. J., Paulus, M. P., & Stein, M. B. (2011). A reverse translational approach to quantify approach-avoidance conflict in humans. Behavioural brain research, 225(2), 455–463.

Barlow, D. H., Allen, L. B., & Choate, M. L. (2016). Toward a Unified Treatment for Emotional Disorders – Republished Article. Behavior Therapy, 47(6), 838–853. 10.1016/j.beth.2016.11.005

Bohn, M., Babor, T., & Kranzler, H. (1991). Validity of the Drug Abuse Screening Test (DAST-10) in inpatient substance abusers. Problems of drug dependence, 119, 233–235.

Carver, C. S., & White, T. L. (1994). Behavioral inhibition, behavioral activation, and affective responses to impending reward and punishment: the BIS/BAS scales. Journal of personality and social psychology, 67(2), 319.

Cella, D., Riley, W., Stone, A., Rothrock, N., Reeve, B., Yount, S., Amtmann, D., Bode, R., Buysse, D., & Choi, S. (2010). The Patient-Reported Outcomes Measurement Information System (PROMIS) developed and tested its first wave of adult self-reported health outcome item banks: 2005–2008. Journal of clinical epidemiology, 63(11), 1179–1194.

Chrysikou, E. G., Gorey, C., & Aupperle, R. L. (2017). Anodal transcranial direct current stimulation over right dorsolateral prefrontal cortex alters decision making during approach-avoidance conflict. Social cognitive and affective neuroscience, 12(3), 468–475.

Cicchetti, D. V. (1994). Guidelines, criteria, and rules of thumb for evaluating normed and standardized assessment instruments in psychology. Psychological Assessment, 6(4), 284–290. 10.1037/1040-3590.6.4.284

Dalgleish, T., Black, M., Johnston, D., & Bevan, A. (2020). Transdiagnostic approaches to mental health problems: Current status and future directions. Journal of Consulting and Clinical Psychology, 88(3), 179–195. 10.1037/ccp0000482

Ekhtiari, H., Kuplicki, R., Yeh, H.-w., & Paulus, M. P. (2019). Physical characteristics not psychological state or trait characteristics predict motion during resting state fMRI. Scientific Reports, 9(1), 419. 10.1038/s41598-018-36699-0

Friston, K., Mattout, J., Trujillo-Barreto, N., Ashburner, J., & Penny, W. (2007). Variational free energy and the Laplace approximation. NeuroImage, 34(1), 220–234.

Gard, D. E., Gard, M. G., Kring, A. M., & John, O. P. (2006). Anticipatory and consummatory components of the experience of pleasure: a scale development study. J Res Pers, 40(6), 1086–1102.

Gobinath, A. R., Choleris, E., & Galea, L. A. M. (2017). Sex, hormones, and genotype interact to influence psychiatric disease, treatment, and behavioral research. Journal of Neuroscience Research, 95(1-2), 50–64. 10.1002/jnr.23872

Green, T., Flash, S., & Reiss, A. L. (2019). Sex differences in psychiatric disorders: what we can learn from sex chromosome aneuploidies. Neuropsychopharmacology, 44(1), 9–21. 10.1038/s41386-018-0153-2

Hester, R., Bell, R. P., Foxe, J. J., & Garavan, H. (2013). The influence of monetary punishment on cognitive control in abstinent cocaine-users. Drug and Alcohol Dependence, 133(1), 86–93. 10.1016/j.drugalcdep.2013.05.027

Huys, Q. J. M., Browning, M., Paulus, M. P., & Frank, M. J. (2021). Advances in the computational understanding of mental illness. Neuropsychopharmacology, 46(1), 3–19. 10.1038/s41386-020-0746-4

Johnstone, B., Callahan, C. D., Kapila, C. J., & Bouman, D. E. (1996). The comparability of the WRAT-R reading test and NAART as estimates of premorbid intelligence in neurologically impaired patients. Archives of Clinical Neuropsychology, 11(6), 513–519.

Kelly, M. M., Tyrka, A. R., Price, L. H., & Carpenter, L. L. (2008). Sex differences in the use of coping strategies: predictors of anxiety and depressive symptoms. Depression and Anxiety, 25(10), 839–846. 10.1002/da.20341

Kessler, R. C., Sampson, N. A., Berglund, P., Gruber, M. J., Al-Hamzawi, A., Andrade, L., Bunting, B., Demyttenaere, K., Florescu, S., de Girolamo, G., Gureje, O., He, Y., Hu, C., Huang, Y., Karam, E., Kovess-Masfety, V., Lee, S., Levinson, D., Medina Mora, M. E., … Wilcox, M. A. (2015). Anxious and non-anxious major depressive disorder in the World Health Organization World Mental Health Surveys. Epidemiology and Psychiatric Sciences, 24(3), 210–226. 10.1017/S2045796015000189

Kirlic, N., Young, J., & Aupperle, R. L. (2017). Animal to human translational paradigms relevant for approach avoidance conflict decision making. Behaviour research and therapy, 96, 14–29. 10.1016/j.brat.2017.04.010

Klein Hofmeijer-Sevink, M., Batelaan, N. M., van Megen, H. J. G. M., Penninx, B. W., Cath, D. C., van den Hout, M. A., & van Balkom, A. J. L. M. (2012). Clinical relevance of comorbidity in anxiety disorders: A report from the Netherlands Study of Depression and Anxiety (NESDA). Journal of Affective Disorders, 137(1), 106–112. 10.1016/j.jad.2011.12.008

Kroenke, K., Spitzer, R. L., & Williams, J. B. W. (2001). The PHQ-9: Validity of a brief depression severity measure [doi:10.1046/j.1525-1497.2001.016009606.x].

Kuhn, M. (2008). Building Predictive Models in R Using the caret Package. Journal of Statistical Software, 28(5), 1–26. 10.18637/jss.v028.i05

Lang, P. J., & Bradley, M. M. (1999). International affective digitized sounds (IADS): Stimuli, instruction manual and affective ratings. In Technical Report no. b-2 (Vol. Vol. 803).

Lang, P. J., Bradley, M. M., & Cuthbert, B. N. (2008). International affective picture system (IAPS): Affective ratings of pictures and instruction manual. In. Technical Report A-8: University of Florida, Gainesville, FL.

Lavalley, C. A., Hakimi, N., Taylor, S., Kuplicki, R., Forthman, K. L., Stewart, J. L., Paulus, M. P., Khalsa, S. S., & Smith, R. (2024). Transdiagnostic failure to adapt interoceptive precision estimates across affective, substance use, and eating disorders: A replication and extension of previous results. Biol Psychol, 191, 108825. 10.1016/j.biopsycho.2024.108825

Letkiewicz, A. M., Kottler, H. C., Shankman, S. A., & Cochran, A. L. (2023). Quantifying aberrant approach-avoidance conflict in psychopathology: A review of computational approaches. Neuroscience and biobehavioral reviews, 147, 105103. 10.1016/j.neubiorev.2023.105103

Matud, M. P. (2004). Gender differences in stress and coping styles. Personality and Individual Differences, 37(7), 1401–1415. 10.1016/j.paid.2004.01.010

Norman, S. B., Hami Cissell, S., Means-Christensen, A. J., & Stein, M. B. (2006). Development and validation of an Overall Anxiety Severity And Impairment Scale (OASIS). Depression and Anxiety, 23(4), 245–249. 10.1002/da.20182

Panayiotou, G., Karekla, M., & Leonidou, C. (2017). Coping through avoidance may explain gender disparities in anxiety. Journal of Contextual Behavioral Science, 6(2), 215–220. 10.1016/j.jcbs.2017.04.005

Paulus, M. P. (2017). Evidence-Based Pragmatic Psychiatry—A Call to Action. JAMA Psychiatry, 74(12), 1185–1186. 10.1001/jamapsychiatry.2017.2439

Plana-Ripoll, O., Musliner, K. L., Dalsgaard, S., Momen, N. C., Weye, N., Christensen, M. K., Agerbo, E., Iburg, K. M., Laursen, T. M., Mortensen, P. B., Pedersen, C. B., Petersen, L. V., Santomauro, D. F., Vilhjálmsson, B. J., Whiteford, H. A., & McGrath, J. J. (2020). Nature and prevalence of combinations of mental disorders and their association with excess mortality in a population-based cohort study. World Psychiatry, 19(3), 339–349. 10.1002/wps.20802

Plana-Ripoll, O., Pedersen, C. B., Holtz, Y., Benros, M. E., Dalsgaard, S., de Jonge, P., Fan, C. C., Degenhardt, L., Ganna, A., Greve, A. N., Gunn, J., Iburg, K. M., Kessing, L. V., Lee, B. K., Lim, C. C. W., Mors, O., Nordentoft, M., Prior, A., Roest, A. M., … McGrath, J. J. (2019). Exploring Comorbidity Within Mental Disorders Among a Danish National Population. JAMA Psychiatry, 76(3), 259–270. 10.1001/jamapsychiatry.2018.3658

Riecher-Rössler, A. (2017). Sex and gender differences in mental disorders. The Lancet Psychiatry, 4(1), 8–9. 10.1016/S2215-0366(16)30348-0

Rutter, M., Caspi, A., & Moffitt, T. E. (2003). Using sex differences in psychopathology to study causal mechanisms: unifying issues and research strategies. Journal of Child Psychology and Psychiatry, 44(8), 1092–1115. 10.1111/1469-7610.00194

Sandin, B., Chorot, P., & McNally, R. J. (2001). Anxiety sensitivity index: normative data and its differentiation from trait anxiety. Behav Res Ther, 39(2), 213–219. 10.1016/s0005-7967(00)00009-7

Sheehan, D. V., Lecrubier, Y., Sheehan, K. H., Amorim, P., Janavs, J., Weiller, E., Hergueta, T., Baker, R., & Dunbar, G. C. (1998). The Mini-International Neuropsychiatric Interview (M.I.N.I.): the development and validation of a structured diagnostic psychiatric interview for DSM-IV and ICD-10. J Clin Psychiatry, 59 *Suppl 20*, 22–33;quiz 34-57.

Simons, J. S., & Arens, A. M. (2007). Moderating effects of sensitivity to punishment and sensitivity to reward on associations between marijuana effect expectancies and use. Psychology of Addictive Behaviors, 21(3), 409–414. 10.1037/0893-164X.21.3.409

Simons, J. S., Dvorak, R. D., & Batien, B. D. (2008). Methamphetamine use in a rural college population: Associations with marijuana use, sensitivity to punishment, and sensitivity to reward. Psychology of Addictive Behaviors, 22(3), 444–449. 10.1037/0893-164X.22.3.444

Sinha, S., & Latha, G. (2018). Coping response to same stressors varies with gender. National J Physiol Pharm Pharmacol, 7, 1053–1057.

Smith, R., Feinstein, J. S., Kuplicki, R., Forthman, K. L., Stewart, J. L., Paulus, M. P., Tulsa, I., & Khalsa, S. S. (2021a). Perceptual insensitivity to the modulation of interoceptive signals in depression, anxiety, and substance use disorders. Sci Rep, 11(1), 2108. 10.1038/s41598-021-81307-3

Smith, R., Kirlic, N., Stewart, J. L., Touthang, J., Kuplicki, R., Khalsa, S. S., Feinstein, J. S., Paulus, M. P., & Aupperle, R. L. (2021b). Greater decision uncertainty characterizes a transdiagnostic patient sample during approach-avoidance conflict: a computational modelling approach. Journal of Psychiatry and Neuroscience, 46(1), E74–E87.

Smith, R., Kirlic, N., Stewart, J. L., Touthang, J., Kuplicki, R., McDermott, T. J., Taylor, S., Khalsa, S. S., Paulus, M. P., & Aupperle, R. L. (2021c). Long-term stability of computational parameters during approach-avoidance conflict in a transdiagnostic psychiatric patient sample. Scientific Reports, 11(1), 11783.

Smith, R., Kuplicki, R., Feinstein, J., Forthman, K. L., Stewart, J. L., Paulus, M. P., Tulsa, i., & Khalsa, S. S. (2020). A Bayesian computational model reveals a failure to adapt interoceptive precision estimates across depression, anxiety, eating, and substance use disorders. PLoS Computational Biology, 16(12), e1008484. 10.1371/journal.pcbi.1008484

Smith, R., Lavalley, C. A., Taylor, S., Stewart, J. L., Khalsa, S. S., Berg, H., Ironside, M., Paulus, M. P., & Aupperle, R. L. (2023). Elevated decision uncertainty and reduced avoidance drives in depression, anxiety and substance use disorders during approach–avoidance conflict: a replication study. Journal of Psychiatry and Neuroscience, 48(3), E217–E231.

Spielberger, C. D., Gorsuch, R. L., & Lushene, R. E. (1970). Manual for the State-Trait Anxiety Inventory. Consulting Psychologists Press.

Stewart, J. L., May, A. C., Poppa, T., Davenport, P. W., Tapert, S. F., & Paulus, M. P. (2014). You are the danger: Attenuated insula response in methamphetamine users during aversive interoceptive decision-making. Drug and Alcohol Dependence, 142, 110–119. 10.1016/j.drugalcdep.2014.06.003

Victor, T. A., Khalsa, S. S., Simmons, W. K., Feinstein, J. S., Savitz, J., Aupperle, R. L., Yeh, H.- W., Bodurka, J., & Paulus, M. P. (2018). Tulsa 1000: a naturalistic study protocol for multilevel assessment and outcome prediction in a large psychiatric sample. BMJ open, 8(1).

Watson, D., Clark, L. A., & Tellegen, A. (1988). Development and validation of brief measures of positive and negative affect: the PANAS scales. Journal of personality and social psychology, 54(6), 1063.

Weinberg, A., Perlman, G., Kotov, R., & Hajcak, G. (2016). Depression and reduced neural response to emotional images: Distinction from anxiety, and importance of symptom dimensions and age of onset [doi:10.1037/abn0000118].

## References

1. Smith, R., et al., Elevated decision uncertainty and reduced avoidance drives in depression, anxiety and substance use disorders during approach–avoidance conflict: a replication study. Journal of Psychiatry and Neuroscience, 2023. 48(3): p. E217–E231.

